# What Australian Health Practitioner Tribunals Decide, and What Automated Analysis Misses: 3,586 Decisions, 1999 to February 2026

**DOI:** 10.64898/2026.02.13.26346299

**Authors:** Hayden Farquhar

**Affiliations:** Independent Researcher, Finley, New South Wales, Australia

**Keywords:** health practitioner regulation, professional misconduct, disciplinary tribunals, National Law, natural language processing, validation, opioid prescribing, Ahpra, Australia

## Abstract

Published tribunal decisions are the public record of the most serious health practitioner discipline in Australia, but they have not been analysed at scale. This study assembled 3,586 tribunal decisions (December 1999 to February 2026) from all eight jurisdictions. The author coded a random sample of 160 decisions, stratified by period and weighted to the corpus, blind to any machine output; these codes give the primary estimates. An independent second coder recoded 60 of them: agreement was substantial or better for disciplinary status (kappa 0.79) and most other fields, but moderate for dishonesty or fraud (0.59) and professional conduct (0.60). Among disciplinary decisions, breaches of professional conduct were found in 68.7% (95% CI 58.9–77.5), dishonesty or fraud in 41.6% (29.7–53.1), clinical competence failures in 38.6% (27.5–50.3), and boundary violations in 36.3% (24.8–48.0). Professional misconduct was found in 83.0%, and registration was cancelled in 30.6%. Across the ten professions regulated since 2010, disciplinary decisions rose from 1.25 to 2.36 per 10,000 registrants between 2011–2013 and 2023–2025. A text classifier, trained on a separate gold standard and applied to the whole corpus, identified disciplinary decisions accurately (accuracy 0.98) but missed almost half of dishonesty matters, and it detected them more often in later decisions, so its rising fraud trend partly reflects changing classifier error. Automated corpus-wide trends need checking against representative human coding across time.

## I. Introduction

The regulation of health practitioners in Australia was substantially reformed by the enactment of the Health Practitioner Regulation National Law (‘National Law’) in Queensland in 2009, subsequently adopted by all states and territories through an applied laws scheme.^1^ This National Registration and Accreditation Scheme replaced a patchwork of state-based regulatory bodies with a unified framework administered by the Australian Health Practitioner Regulation Agency (Ahpra) and 15 National Boards, responsible for the registration and regulation of practitioners across 16 health professions.^2^ At 30 June 2024, the scheme regulated 920,535 registered practitioners under a single national law, whereas in the United Kingdom health and care professions are regulated by ten separate statutory regulators.^3^ The National Law establishes a tiered system for addressing conduct concerns: health complaints entities in each state and territory receive and triage notifications, with less serious matters managed through conditions, undertakings, or counselling, while serious matters (those involving professional misconduct, a pattern of unsatisfactory professional conduct, or conduct warranting suspension or cancellation of registration) are referred to state and territory tribunals for formal adjudication.^4^ Plain-language definitions of the accuracy measures used in this article are given in a note.^5^

These tribunals, including the NSW Civil and Administrative Tribunal (NCAT), the Victorian Civil and Administrative Tribunal (VCAT), the Queensland Civil and Administrative Tribunal (QCAT), the South Australian Civil and Administrative Tribunal (SACAT), and the State Administrative Tribunal of Western Australia (SAT), issue published decisions that together form a large, publicly accessible record of regulatory adjudication. Decisions are typically published on the Australasian Legal Information Institute (Aust-LII) and give detailed accounts of the allegations, evidence, findings, and orders in each matter.

The framework that governs these decisions has changed repeatedly. Amendments in 2022 made protection of the public and public confidence in the safety of health services the paramount principle of the scheme, and gave National Boards a narrow discretion not to refer a professional-misconduct matter to a tribunal where there is no public interest in a hearing.^6^ Amendments passed in 2025 require tribunal findings of professional misconduct based on sexual misconduct to be recorded permanently on the public register, retrospectively to decisions made from July 2010, and require a cancelled practitioner to obtain a reinstatement order from a tribunal before reapplying for registration; both commenced in April 2026.^7^ In 2025 an independent review of the scheme was unable to say whether tribunals in different jurisdictions treat similar cases differently, and recommended longitudinal research on tribunal outcomes; in May 2026 Health Ministers accepted a recommendation that Ahpra research tribunal outcomes for 2020 to 2025.^8^ What tribunals have actually decided, and how that has changed, is therefore a live regulatory question.

The existing literature on health practitioner disciplinary proceedings in Australia has relied on manual coding. Millbank analysed 794 tribunal cases of serious misconduct or impairment brought against the five most populous health professions between July 2010 and June 2017, finding marked variation in outcomes between professions and between jurisdictions, and that sexual misconduct attracted the most severe outcomes.^9^ Kremer recently analysed 107 published tribunal decisions involving psychologists between 2016 and 2025, and found boundary violations or multiple relationships in 82 of them.^10^ Bismark, Spittal, Studdert and colleagues have analysed national complaint and notification data, identifying male sex and older age as risk factors for complaints, profession-specific differences in outcomes, and a higher risk of complaints against international medical graduates.^11^ Elkin and colleagues examined 485 doctors disciplined for professional misconduct in four Australian states and New Zealand between 2000 and 2009, developing a typology of misconduct that later work has adapted.^12^ In New Zealand, Surgenor and colleagues found that differences in disciplinary penalties between professions were largely accounted for by whether the practitioner attended the hearing and was legally represented, and that tribunals deciding whether to impose conditions commonly cited remorse or insight.^13^ Manual coding has limited these studies to a single profession, a subset of professions, or a restricted period.

Natural language processing (NLP), the use of computational methods to analyse text, has been applied in other legal domains to predict judicial decisions and classify legal documents.^14,15,16^ These methods make analysis of whole legal corpora practical,^17,18^ and machine-learning and language-model methods have recently been applied to health-related court decisions in South Korea and to complaint investigation reports in New Zealand.^19^ Topic modelling has been used to explore United Kingdom fitness-to-practise de-cisions,^20^ and the use of these methods in legal epidemiology is now being examined critically.^21^ To the author’s knowledge, however, no study has validated automated classification of health practitioner disciplinary decisions against representative human coding, or used such classification to estimate what tribunals decide. Automated labels carry measurement error, however, and an error rate estimated on a small or un-representative test set may not hold across a whole corpus or across time. A classifier whose accuracy changes over the study period can produce a trend that is not in the decisions.

This study combined the two approaches. A random sample of decisions, stratified by period and coded by hand, gives the primary estimates of what tribunals decide and how that has changed. A classifier applied to the full corpus gives exploratory, larger-scale analyses, and the coded sample measures how far each of them can be trusted. The study asked which kinds of misconduct, findings, and outcomes make up tribunal discipline across all regulated professions and jurisdictions; how these, and the number of disciplinary decisions relative to the registered workforce, have changed over 27 years; how accurately an automated classifier and a logged language-model coder reproduce human coding across the corpus and across time; and what the full corpus suggests, with that accuracy in view, about variation by profession and jurisdiction, prescribing misconduct, and boundary violations.

## II. Methods

This was a retrospective, cross-sectional, descriptive analysis of publicly available tribunal decisions involving health practitioners. No single reporting guideline fits this design; the methods draw on elements of STROBE (for the descriptive epidemiology) and TRIPOD+AI (for the classifier development and val-idation).^22^ Choices for the classifier, the second test sample, and the language-model coder were fixed in written protocols before the relevant data were scored; departures from them are reported as departures.

### A. Data Acquisition

Tribunal decisions were sourced from three repositories to maximise corpus coverage. The primary source was the Australasian Legal Information Institute (AustLII), Australia’s principal open-access legal database, from which 2,043 decisions were retained after keyword filtering across seven jurisdictions (Victoria, Queensland, South Australia, Western Australia, Australian Capital Territory, Northern Territory, and Tasmania). A registry of 24 tribunal identifiers was constructed to capture both current tribunals (NCAT, VCAT, QCAT, SACAT, SAT, ACAT, NTCAT) and legacy state-specific health practitioner tribunals that pre-dated the National Law. Scraping (Python, with requests and BeautifulSoup) observed the crawl delay that AustLII specifies for automated access (120 seconds between requests, verified February 2026). Sixty health-related keywords, matched in two tiers, excluded non-health decisions from tribunals with mixed jurisdiction (for example, NCAT also hears matters concerning architects) (Supplementary Appendix B).

A second source was the Open Australian Legal Corpus, hosted on HuggingFace, which provided 1,497 New South Wales decisions originally published on NSW Caselaw.^23^ This dataset was filtered to retain only decisions from the NCAT Occupational Division, the former NSW Medical Tribunal, and NCAT appeals, using the same keyword filter.

A third source was the Internet Archive’s Wayback Machine, from which 46 decisions were recovered from three legacy tribunals (Northern Territory Health Professional Review Tribunal, Tasmanian Health Practitioners Tribunal, and TasCAT) whose AustLII pages had been permanently removed (HTTP 410 Gone). Six further legacy tribunals had no archived content on either source, a known gap in coverage.

After deduplication on citation identifiers, the final corpus comprised 3,586 decisions (Figure 1) dated from 15 December 1999 to 9 February 2026 (the corpus was assembled in February 2026), drawn from 24 tribunals across eight jurisdictions (Table 1).

**Figure 1.**
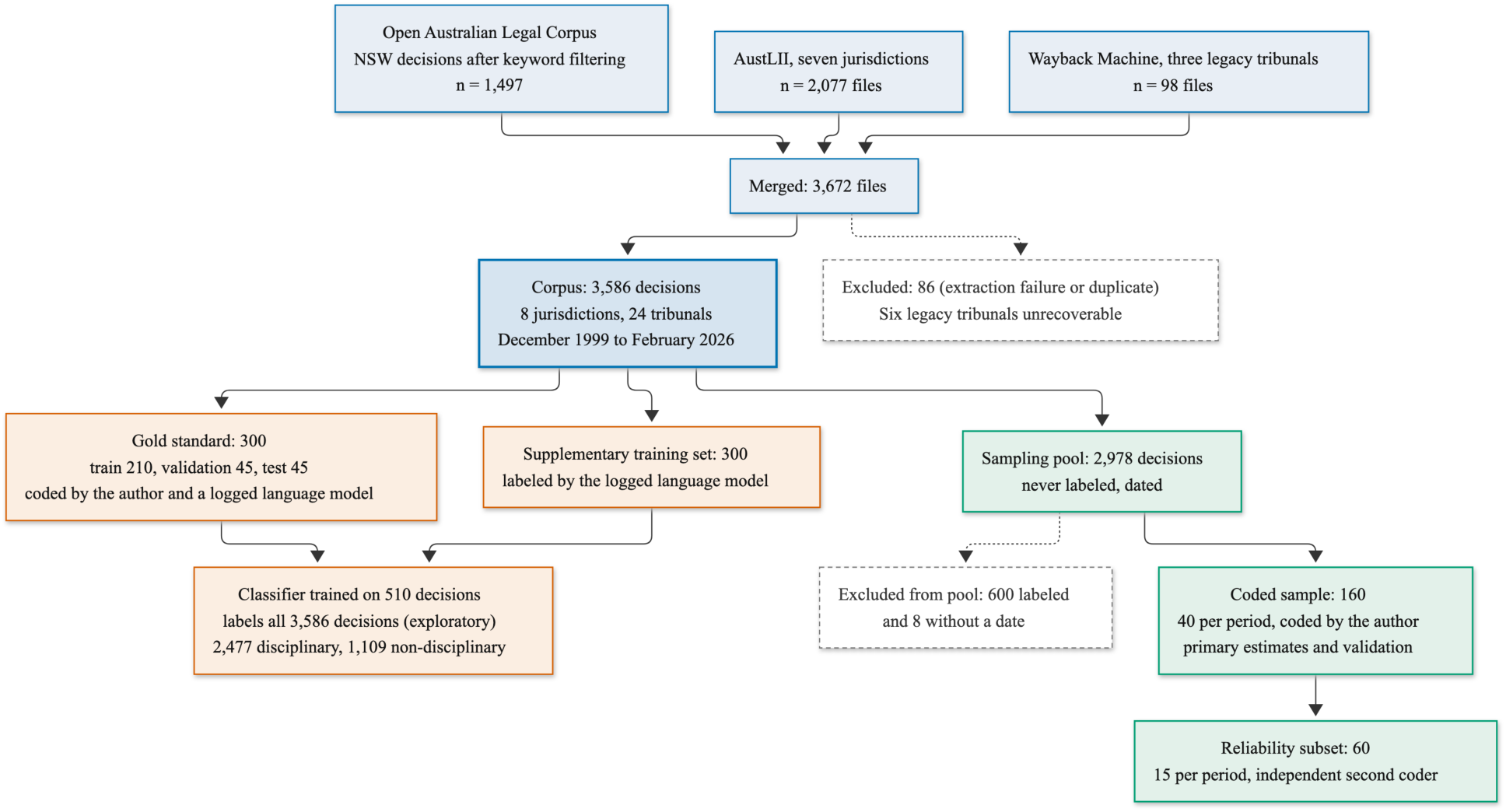
Corpus assembly and the samples used in the study

**Table 1.**
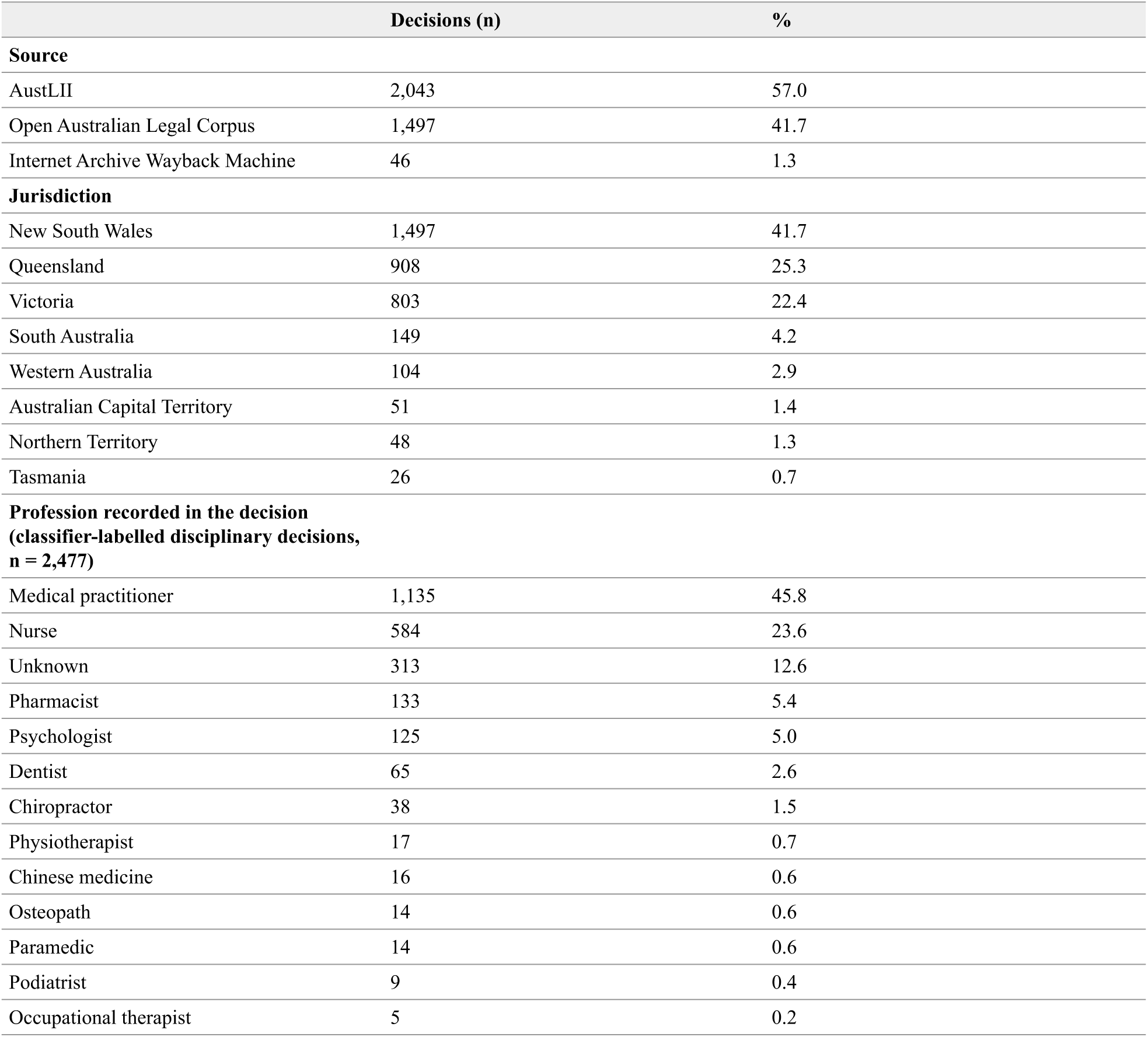
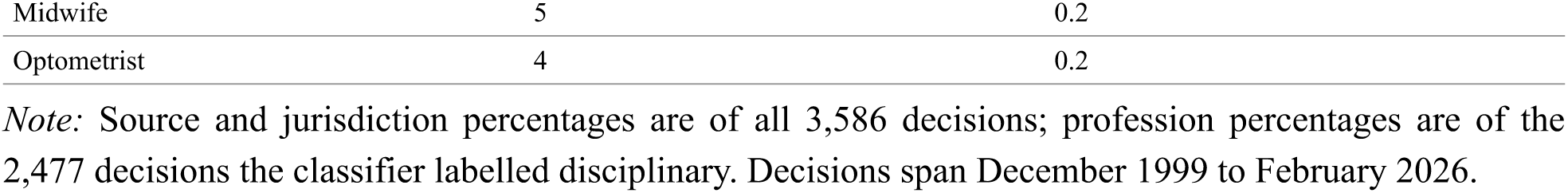
Corpus overview: sources, jurisdictions, and profession distribution (N = 3,586)

|  | Decisions (n) | % |
|---|---|---|
| **Source** |  |  |
| AustLII | 2,043 | 57.0 |
| Open Australian Legal Corpus | 1,497 | 41.7 |
| Internet Archive Wayback Machine | 46 | 1.3 |
| **Jurisdiction** |  |  |
| New South Wales | 1,497 | 41.7 |
| Queensland | 908 | 25.3 |
| Victoria | 803 | 22.4 |
| South Australia | 149 | 4.2 |
| Western Australia | 104 | 2.9 |
| Australian Capital Territory | 51 | 1.4 |
| Northern Territory | 48 | 1.3 |
| Tasmania | 26 | 0.7 |
| **Profession recorded in the decision (classifier-labelled disciplinary decisions, n = 2,477)** |  |  |
| Medical practitioner | 1,135 | 45.8 |
| Nurse | 584 | 23.6 |
| Unknown | 313 | 12.6 |
| Pharmacist | 133 | 5.4 |
| Psychologist | 125 | 5.0 |
| Dentist | 65 | 2.6 |
| Chiropractor | 38 | 1.5 |
| Physiotherapist | 17 | 0.7 |
| Chinese medicine | 16 | 0.6 |
| Osteopath | 14 | 0.6 |
| Paramedic | 14 | 0.6 |
| Podiatrist | 9 | 0.4 |
| Occupational therapist | 5 | 0.2 |
| Midwife | 5 | 0.2 |
| Optometrist | 4 | 0.2 |

*Note:* Source and jurisdiction percentages are of all 3,586 decisions; profession percentages are of the 2,477 decisions the classifier labelled disciplinary. Decisions span December 1999 to February 2026.

Text was extracted from HTML with inscriptis,^24^ the converter used to build the Open Australian Legal Corpus, from RTF files with striprtf, and from PDF files with PyMuPDF. Citation, date, practitioner name, profession, and jurisdiction were extracted from header fields and the decision text with regular expressions, profession combining structured metadata with keyword matching (for example, ‘Medical Board’), and decisions were divided into background, allegations, evidence, findings, and orders by heading patterns.

### B. Coding Scheme and Gold Standard

A seven-category misconduct taxonomy was developed from the categories of concern reported by Ahpra and the typology of Elkin and colleagues, as adapted by Millbank for tribunal decisions under the National Law, with dishonesty or fraud and communication failures added as separate categories:^25^

1. **Clinical competence** (CLIN): deficiencies in clinical knowledge, skill, or care delivery
2. **Prescribing misconduct** (PRESC): inappropriate prescribing, dispensing, or supply of medications
3. **Boundary violations** (BOUND): sexual, emotional, or financial boundary violations in the practitioner–patient relationship
4. **Professional conduct** (PROF): breaches of professional standards, codes of conduct, or registration conditions
5. **Dishonesty/fraud** (FRAUD): dishonest conduct including fraudulent billing, false records, or misrepresentation
6. **Impairment** (IMPR): health impairment affecting fitness to practise, including substance use disorders
7. **Communication failures** (COMM): failures in communication with patients, colleagues, or regulators

Each category was further divided into subcategories (34 in total), with detailed definitions and coding rules documented in annotation guidelines. Misconduct types were multi-label: a single decision could be assigned several, reflecting the overlapping nature of many disciplinary proceedings. Under the coding rules, professional conduct includes breaches of registration conditions and criminal conduct outside practice, so it is the broadest category. Boundary violations were further coded as sexual, emotional, or financial where the decision described that kind of breach.

A nine-category outcome taxonomy was applied: cancellation, suspension, conditions, reprimand, caution, fine, undertaking, no adverse finding, and not applicable (for non-disciplinary decisions). Non-standard orders (for example disqualification of an unregistered person, prohibition orders, and costs-only orders) were coded as ‘other’. A four-category finding taxonomy distinguished professional misconduct, unsatisfactory professional conduct (satisfactory), unprofessional conduct (general), and no finding. A decision was disciplinary if it decided whether the practitioner’s conduct, performance or health warranted findings or sanctions, including appeals and health or impairment proceedings; registration appeals, reviews of immediate action, and purely procedural rulings were non-disciplinary.

A gold standard of 300 decisions, a random sample stratified by jurisdiction, profession, and period (210 training, 45 validation, 45 test), was used to train and first test the classifier. Each decision was coded twice under the same written guidelines. The first coder was the author, who coded every decision from a packet containing only the decision text and neutral metadata (citation, tribunal, date, and URL), in randomised order and blind to all earlier labels and to any classifier output. The second coder was a language model, Claude Sonnet 5.5 (Anthropic), run once through the Anthropic Message Batches API with a fixed system prompt containing the guidelines and a structured output schema, following emerging practice in model-assisted annotation.^26,27^ Every request and response was logged with the model identifier, prompt hash, and token usage, and no model output was edited. Agreement (Cohen’s kappa, 2,000 bootstrap resamples) was high for disciplinary status (0.97), finding (0.94), and profession (0.95), and ranged from 0.60 to 0.90 for misconduct types (Supplementary Table S25). The author adjudicated all 268 disagreeing fields (in 158 decisions) against the decision text and the guidelines, keeping the author’s code for 127 and adopting the model’s for 141. This step was not blinded, since each disagreement displayed both codes; the agreement statistics come from the frozen blind codes. One further field was amended after adjudication.^28^

### C. Representative Coded Sample

Because the gold standard was stratified by profession and jurisdiction, it does not represent the corpus. A second sample was therefore drawn under a protocol written before sampling. The sampling pool was every decision that had never been labelled by anyone, with at least 500 characters of text and a known date (2,978 decisions; the 600 decisions excluded comprise the gold standard and the decisions used for classifier training). The pool was divided into four periods by decision year (1999–2009, 2010–2014, 2015–2019, and 2020–2026; 2010 marks the start of the National Law), containing 77, 416, 829, and 1,656 decisions. Forty decisions were drawn at random from each period (160 in total), so that early years were represented well enough to compare periods, and the order was shuffled. Estimates for the corpus were weighted back to the pool (weight = pool size of the period / 40).

The author coded the 160 decisions alone, from packets containing only the decision text and neutral metadata, under the same fields and rules as the gold standard, blind to every label and prediction, with no model pass and no adjudication. A first coding was set aside, and its results not used, after checks showed that for many rows the codes and notes described a decision other than the one in that row’s text. The author recoded all 160 decisions with a new workbook that required, for each row, a name taken from that row’s text (found for 160 of 160 rows). This recode, a departure from the protocol, was recorded in dated amendments before the recoded sample was scored; when recoding, the author knew the aggregate results of the first scoring but had seen no prediction for any individual decision (Supplementary Appendix G).

To measure reliability, an independent second coder recoded a random subset of 60 of the 160 decisions (15 per period) under a protocol fixed before recruitment. The coder was recruited through the Prolific platform^29^ from participants with a postgraduate degree in law or a health discipline, fluent English, and an approval rate of at least 99%. Five candidates were paid to code four practice decisions from the gold-standard training set (all five were paid whether or not they were selected). The earliest candidate to meet every objective criterion was selected: a complete export, the correct disciplinary status for all four decisions, the explicitly stated orders recorded, a decision-specific note, and a median of at least four minutes per decision. Agreement with the author on judgement-call fields was not used, so that selection would not favour a coder who resembled the author. The coder used the same guidelines, field rules, and texts as the author, in an in-browser workbook, with no training beyond the guidelines and the practice decisions and no feedback on agreement with the author, and coded two sessions of 30 decisions about 14 hours apart. The coder was blind to the author’s codes, to all machine output, and to the manuscript, and attested at the end of each session to working alone, from the text shown, without AI tools. Payment was £12 for the screener and £48 per session (about £22 an hour at the time taken). Each returned file was frozen with a cryptographic hash before any comparison. Agreement was measured with Cohen’s kappa and described with conventional benchmarks (0.41–0.60 moderate, 0.61–0.80 substantial, above 0.80 almost perfect).^30^ The second coder’s codes are used only to report agreement and a sensitivity comparison of prevalence: they do not replace or adjust the author’s codes, and no disagreement was adjudicated. Before the second session the coder was reminded to read each decision in full, an instruction not in the protocol (Supplementary Appendix H).

### D. Classifier and Corpus-Wide Labels

#### Misconduct classification

A multi-label text classifier assigned misconduct types to each decision from its full text, using term frequency–inverse document frequency (TF-IDF) features^31^ and one-vs-rest logistic regression, with a ‘not applicable’ category for non-disciplinary decisions. Classifiers of the same design predicted the statutory finding and whether a decision was disciplinary. Training followed a protocol written before any model was trained. Two training sets were fixed in advance: the 210 gold-standard training decisions alone, and those 210 plus 300 further corpus decisions (a random sample stratified by jurisdiction) labelled by the same logged language-model pass used as second coder. Regularisation strength was chosen from a fixed grid on the 45 validation decisions, and the 510-decision training set was selected on validation performance for all three classifiers. The 45-decision test set was scored once, after these choices were fixed. A linear model was preferred to a fine-tuned transformer such as Legal-BERT^32^ for interpretability and because the training set is small. Configuration details are in Supplementary Appendix E.

#### Outcome extraction

Outcomes were extracted with regular expressions targeting the orders and decision sections of each decision, using the statutory language of the National Law and common tribunal phrasing (for example, ‘registration is cancelled’ or ‘suspended for a period of [X] months’).

#### Named entity recognition

A rule-based system identified mentions of medications (92 drug names including brand and generic forms), medical conditions (42 terms), and regulatory boards (26 entities). Each medication was mapped to its schedule or schedules, and its Appendix D status, in the current Poisons Standard (October 2026),^33^ which may differ from scheduling at the time of the conduct.

#### Boundary subcategories and topics

Decisions classified as boundary violations were subcategorised as sexual, emotional, or financial by keywords in the full text (for example ‘sexual’, ‘dual relationship’, and ‘loan’), and an unsupervised topic model (BERTopic)^34^ identified 9 topics that mainly reflected jurisdictional and structural variation (Supplementary Appendix F).

The trained models were applied to all 3,586 decisions. Every corpus-wide result in this article that depends on misconduct types, findings, or outcomes rests on these automated labels and is reported as exploratory.

### E. Validation and the Language-Model Coder

The classifier was evaluated first on the 45 gold-standard test decisions and then, with nothing retrained or re-selected, on the 160-decision coded sample, using the same metrics (macro-F1 and micro-F1 for misconduct types, accuracy for disciplinary status and finding, and micro-F1 for outcomes), weighted to the pool on the coded sample with 95% confidence intervals from 10,000 bootstrap resamples within periods. Two checks addressed change over time: logistic regressions on decision year estimated whether, for each misconduct type, the chance of detecting a true case or of a false positive changed per decade, and the classifier’s predicted prevalence was compared with the coded prevalence by period. A corpus-wide trend was to be called robust only if the two prevalences moved in the same direction and neither error rate showed clear dependence on time; with fewer than 10 coded cases the check could not be completed.

After the classifier had been scored, a second protocol tested whether the logged language-model coder (the same model, prompt, and schema as the gold-standard second coder, run once on the 160 decisions) should replace the classifier for corpus-wide labels. The coder would replace it only if its weighted misconduct macro-F1 was at least 0.70, its disciplinary accuracy at least 0.90, and the 95% confidence interval for its advantage over the classifier excluded zero. If it failed, the protocol made the coded sample the primary evidence on prevalence and the automated corpus-wide results exploratory.

### F. Statistical Analysis

Primary estimates of the share of decisions that were disciplinary, and of misconduct types, boundary subcategories, findings, outcomes, and profession among disciplinary decisions, were weighted ratio estimates from the coded sample with within-period bootstrap confidence intervals (10,000 resamples). Change over time in the coded sample was described by period and tested, for each misconduct type, by logistic regression on decision year among disciplinary decisions (odds ratio per decade); these regressions were unweighted because sampling depended only on year. With about 27 disciplinary decisions per period, a null result is reported as no evidence of change, not as stability; p-values for these seven regressions are reported unadjusted, with Benjamini-Hochberg q-values across the seven types. Because the most recent period holds most of the pool (1,656 of 2,978 decisions), the weighted estimates mainly describe decisions since 2020. Two checks were added after the coded sample had been scored. One compared the classifier’s prevalence in the 600 decisions excluded from the pool with its prevalence in the pool. The other addressed the unit of analysis: one matter can produce several decisions, so each coded decision was re-weighted by the inverse of the number of corpus decisions about the same practitioner (Supplementary Table S32).

For the corpus as a whole, annual counts of disciplinary decisions were tested for monotonic trend with the Mann-Kendall test and Sen’s slope over complete calendar years (1999–2025). Rates per 10,000 registered practitioners were computed for 2011–2025 from Ahpra counts of registrants at 30 June of each year, including non-practising registrants and excluding the temporary pandemic sub-register of 2020–2022,^35^ for the ten professions regulated since July 2010, and trends were estimated with quasi-Poisson regression of annual counts on year with a log-registrant offset; decisions of unknown profession (311) were excluded, and a sensitivity analysis allocated them in proportion to known professions in the same year. In exploratory analyses of the automated labels, Mann-Kendall tests assessed trends in annual misconduct-type proportions (with a sensitivity analysis excluding years before 2005, when annual counts were small); chi-square tests with Cramér’s V, and Fisher’s exact test for 2x2 comparisons, assessed associations between profession, jurisdiction, misconduct type, and outcome; and Benjamini-Hochberg false discovery rate (FDR) correction was applied across all 63 such tests. Rogan-Gladen estimates,^36^ which adjust a clas-sifier’s prevalence for its sensitivity and specificity on the coded sample, are a sensitivity analysis. Analyses used Python 3.12 with scipy 1.17, statsmodels 0.14.6, scikit-learn 1.8, and pymannkendall 1.4.3. Classifier calibration was not assessed, because the classifier was used only to assign labels.^37^

## III. Results

### A. Corpus Characteristics

The corpus comprised 3,586 decisions dated between December 1999 and February 2026 across eight jurisdictions (Table 1). New South Wales (41.7%), Queensland (25.3%), and Victoria (22.4%) contributed 89.5% of decisions. The classifier labelled 2,477 decisions (69.1%) as disciplinary, close to the share in the coded sample (70.3%, 95% CI 61.6–78.6). Medical practitioners (45.8%) and nurses (23.6%) were the most common professions; profession could not be determined from the decision for 313 (12.6%), all dated 2011 or later.

### B. What Tribunals Decide: The Coded Sample

Of the 160 coded decisions, 109 were disciplinary. Weighted to the corpus, breaches of professional conduct were the most common misconduct type, found in 68.7% of disciplinary decisions (95% CI 58.9–77.5), followed by dishonesty or fraud (41.6%; 29.7–53.1), clinical competence failures (38.6%; 27.5–50.3), and boundary violations (36.3%; 24.8–48.0) (Table 2; Figure 2; Supplementary Table S27). Prescribing misconduct was found in 24.7% (14.9–35.3), impairment in 7.5% (2.3–14.2), and communication failures in 5.5% (0.6–12.0). Among the 39 coded boundary-violation decisions, the weighted shares involving sexual, emotional, and financial breaches were 83.6%, 76.4%, and 5.5% (on the emotional share, see the reliability results below).

**Figure 2.**
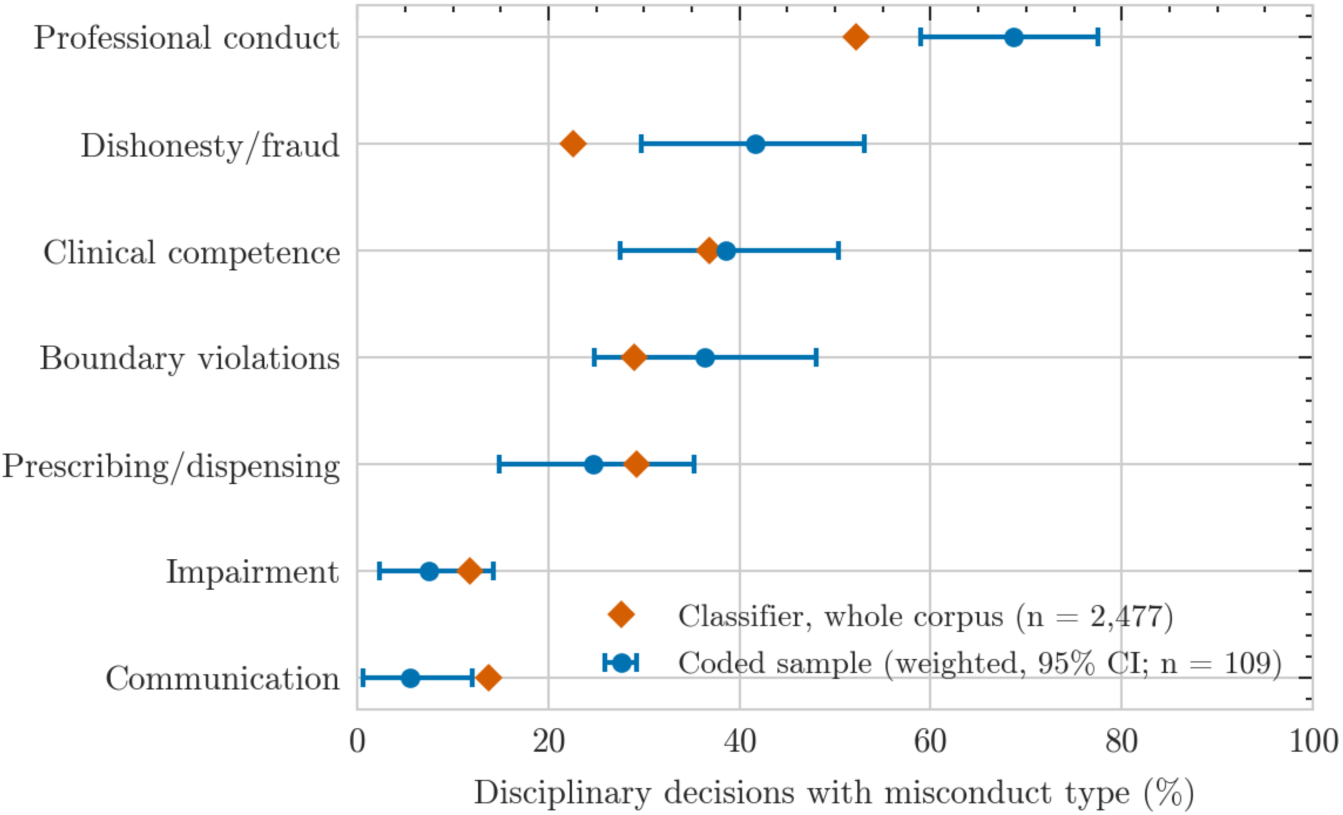
Prevalence of misconduct types among disciplinary decisions: coded sample (weighted, 95% CI) and classifier labels for the whole corpus

**Table 2.** Misconduct types, findings, and outcomes among disciplinary decisions in the coded sample, weighted to the corpus, with the classifier’s corpus-wide prevalence for comparison (n = 109 disciplinary decisions of 160)

|  | **Coded sample, weighted %<br>(95% CI)** | **Coded decisions (n)** | **Classifier, whole corpus %** |
|---|---|---|---|
| **All coded decisions (n = 160)** |  |  |  |
| Disciplinary | 70.3 (61.6–78.6) | 109/160 |  |
| **Misconduct type** |  |  |  |
| Clinical competence | 38.6 (27.5–50.3) | 51/109 | 36.8 |
| Prescribing/dispensing | 24.7 (14.9–35.3) | 31/109 | 29.2 |
| Boundary violations | 36.3 (24.8–48.0) | 39/109 | 29.0 |
| Impairment | 7.5 (2.3–14.2) | 13/109 | 11.8 |
| Dishonesty/fraud | 41.6 (29.7–53.1) | 39/109 | 22.6 |
| Communication | 5.5 (0.6–12.0) | 5/109 | 13.7 |
| Professional conduct | 68.7 (58.9–77.5) | 61/109 | 52.2 |
| **Finding** |  |  |  |
| Professional misconduct | 83.0 (74.7–90.4) | 84/109 |  |
| Unsatisfactory professional conduct | 10.0 (4.2–17.2) | 16/109 |  |
| Unprofessional conduct | 3.0 (0.5–5.9) | 5/109 |  |
| No finding | 4.0 (0.1–9.1) | 4/109 |  |
| **Outcome** |  |  |  |
| Cancellation | 30.6 (19.8–42.4) | 38/109 |  |
| Suspension | 20.8 (11.9–31.1) | 20/109 |  |
| Conditions | 40.0 (28.7–51.6) | 48/109 |  |
| Reprimand | 70.9 (61.0–79.9) | 59/109 |  |
| Fine | 6.6 (1.4–13.1) | 7/109 |  |
| Caution | 1.5 (0.0–3.5) | 3/109 |  |
| Undertaking | 0.1 (0.0–0.3) | 1/109 |  |
| No adverse finding | 2.0 (0.0–4.9) | 2/109 |  |
| Other orders (costs, disqualification, prohibition, other) | 39.2 (27.5–51.1) | 41/109 |  |
| **Profession** |  |  |  |
| Medical practitioner | 41.7 (30.8–53.0) | 68/109 |  |
| Nurse or midwife | 29.7 (18.8–41.0) | 21/109 |  |
| Other profession | 28.7 (17.8–40.0) | 20/109 |  |

*Note:* Coded-sample estimates are weighted to the sampling pool (weight = pool size of the period / 40), with 95% confidence intervals from 10,000 bootstrap resamples within periods. Misconduct types, out-comes, and findings are shares of disciplinary decisions; misconduct types and outcomes are multi-label, so they do not sum to 100%. The classifier column is the share of the 2,477 decisions the classifier la-belled disciplinary (exploratory).

Professional misconduct, the most serious finding, was made in 83.0% of disciplinary decisions (74.7–90.4); unsatisfactory professional conduct in 10.0%, unprofessional conduct in 3.0%, and no finding in 4.0%. Reprimand was the most common order (70.9%; 61.0–79.9), followed by conditions on registration (40.0%), cancellation of registration (30.6%; 19.8–42.4), and suspension (20.8%); fines were ordered in 6.6%, and non-standard orders such as costs, disqualification, or prohibition in 39.2%. Reprimand usually accompanied another order rather than standing alone. Medical practitioners were the subject of 41.7% of disciplinary decisions, nurses and midwives of 29.7%, and other professions of 28.7%.

Neither added check changed the picture: the classifier’s prevalence in the 600 excluded decisions was within 6 percentage points of its prevalence in the pool for every misconduct type, and re-weighting so that each practitioner counted about once moved each type by 2.2 percentage points or less, and reprimand from 70.9% to 77.5% (Supplementary Table S32).

Agreement with the independent second coder on the 60-decision subset was substantial or better for most fields (Supplementary Table S33). Kappa was 0.79 (95% CI 0.60–0.95) for disciplinary status and 0.76 for the finding, and between 0.84 and 0.96 for profession, boundary violations, prescribing misconduct, and the main orders. It was moderate for dishonesty or fraud (0.59; 0.33–0.82) and professional conduct (0.60; 0.39–0.80), and lower for non-standard orders (0.37); communication failures were too rare for kappa. On the subset (unweighted, so these shares differ from the weighted estimates above), the second coder found professional conduct in 37.0% of the decisions they coded as disciplinary, against 58.5% for the author (difference -21.6 percentage points; 95% CI -36.6 to -6.6); no other misconduct type differed by more than 10.3 points (Supplementary Table S34). Of the 90 field-level disagreements, 81 were definitional or concerned the scope of the decision, and only 9 reflected different readings of the text (Supplementary Table S35). All five disagreements on disciplinary status concerned procedural, review, interim, costs, or with-drawn-complaint decisions that the author coded as non-disciplinary. Among the 14 decisions both coders classed as boundary violations, the author recorded an emotional breach in 11 and the second coder in 1.

### C. How Well Automated Labels Reproduce the Coding

On the 45 gold-standard test decisions, the classifier’s misconduct macro-F1 was 0.72 (Table 3; by period in Supplementary Table S30). On the coded sample it was 0.70 (95% CI 0.61–0.77), with disciplinary accuracy 0.98 (0.94–1.00), finding accuracy 0.89, and outcome micro-F1 0.74. Performance differed widely between categories. Boundary violations (F1 0.88) and prescribing misconduct (0.83) were classified well. Professional conduct (0.76) and clinical competence (0.70) were classified with high precision (0.89 and 0.84) but missed about a third and two-fifths of true cases (recall 0.66 and 0.59). Dishonesty or fraud had high precision (0.83) but low recall (0.54), so almost half of coded fraud matters were missed. Impairment (0.54) and communication failures (0.24, precision 0.21, on 5 coded cases) were classified poorly.

**Table 3.** Performance of the classifier on both test sets and of the language-model coder on the coded sample (95% CIs; second test set weighted to the corpus)

| **Measure** | **Classifier, first test set (n = 45)** | **Classifier, coded sample (n = 160, weighted)** | **Language-model coder, coded sample (n = 160, weighted)** |
|---|---|---|---|
| Is-disciplinary accuracy | 0.98 | 0.98 (0.94–1.00) | 0.96 (0.92–0.99) |
| Misconduct macro-F1 | 0.72 | 0.70 (0.61–0.77) | 0.77 (0.69–0.83) |
| Misconduct micro-F1 | 0.78 | 0.78 (0.72–0.83) | 0.82 (0.78–0.87) |
| Finding accuracy | 0.87 | 0.89 (0.83–0.94) | 0.93 (0.88–0.97) |
| Outcome micro-F1 | 0.69 | 0.74 (0.68–0.80) | 0.94 (0.91–0.97) |
| Professional conduct F1 | 0.90 (0.78–0.98) | 0.76 (0.64–0.85) | 0.77 (0.65–0.87) |
| Dishonesty/fraud F1 | 0.53 (0.17–0.80) | 0.66 (0.46–0.81) | 0.85 (0.73–0.94) |
| Clinical competence F1 | 0.83 (0.64–0.96) | 0.70 (0.54–0.83) | 0.81 (0.68–0.91) |
| Boundary violations F1 | 0.86 (0.57–1.00) | 0.88 (0.76–0.96) | 0.91 (0.80–0.98) |
| Prescribing/dispensing F1 | 0.80 (0.50–1.00) | 0.83 (0.67–0.95) | 0.90 (0.77–0.99) |
| Impairment F1 | 0.44 (0.00–0.80) | 0.54 (0.08–0.86) | 0.61 (0.25–0.90) |
| Communication F1 | 0.43 (0.00–0.71) | 0.24 (0.00–0.59) | 0.34 (0.00–0.65) |

*Note:* The first test set is the 45 held-out gold-standard decisions; the coded sample is the representative sample of 160 decisions coded by the author. Confidence intervals are bootstrap percentile intervals; the first-test-set summary measures were reported without intervals.

These error patterns explain the gap between the two sets of estimates in Figure 2. The classifier put dishonesty or fraud at 22.6% of disciplinary decisions against 41.6% in the coded sample, and professional conduct at 52.2% against 68.7%; it put communication failures at 13.7% against 5.5%. Its estimates for clinical competence (36.8%), boundary violations (29.0%), and prescribing (29.2%) fell within the coded sample’s confidence intervals. Regex outcome extraction put cancellation at 39.7% of disciplinary decisions against 30.6% in the coded sample, consistent with its tendency to over-detect cancellation on the first test set. Rogan-Gladen adjustment is unstable with sensitivity as low as 0.29 and is given only as a sensitivity analysis (Supplementary Table S28).

Classifier error also changed over time for one category. Among coded fraud matters, the odds that the classifier detected the matter rose 2.83-fold per decade (95% CI 1.11–7.25). For the other categories with at least 10 coded cases, neither detection nor false positives showed clear dependence on year (Supplementary Table S29). Because communication failures had only 5 coded cases, their error could not be checked over time.

The logged language-model coder was run once on the same 160 decisions, with no failed or refused items. Its weighted macro-F1 was 0.77 (0.69–0.83), with disciplinary accuracy 0.96, finding accuracy 0.93, and outcome micro-F1 0.94. It was much better than the classifier on dishonesty or fraud (F1 0.85), and its detection of fraud did not change with year (odds ratio 0.64 per decade, 0.12–3.35). Its advantage over the classifier in macro-F1 was 0.07, with a 95% confidence interval of -0.01 to 0.15. It therefore met the accuracy thresholds but not the requirement that its advantage be clearly greater than zero, and under the protocol the coded sample became the primary evidence on prevalence.

### D. Change over Time

In the coded sample, the share of disciplinary decisions involving breaches of professional conduct rose from 57.1% in 1999–2009 to 84.4% in 2020–2026 (odds ratio 2.08 per decade, 95% CI 1.16–3.75, p = 0.014; Table 4). Clinical competence failures fell from 57.1% to 31.2% (odds ratio 0.56 per decade, 0.32–1.00, p = 0.051). Neither change is significant after correction across the seven misconduct types (q = 0.10 and 0.18), so both are best read alongside the corpus-wide trends below, which run in the same directions. Dishonesty or fraud was common at both ends of the period (42.9% in 1999–2009 and 50.0% in 2020–2026, with 13.8% in 2010–2014) and showed no clear trend (odds ratio 1.42, 0.79–2.56). There was no evidence of change for boundary violations (0.97, 0.54–1.72), prescribing (0.79, 0.43–1.46), impairment (0.49, 0.20–1.18), or communication failures, but with 20 to 32 disciplinary decisions per period these comparisons could detect only large changes.

**Table 4.**
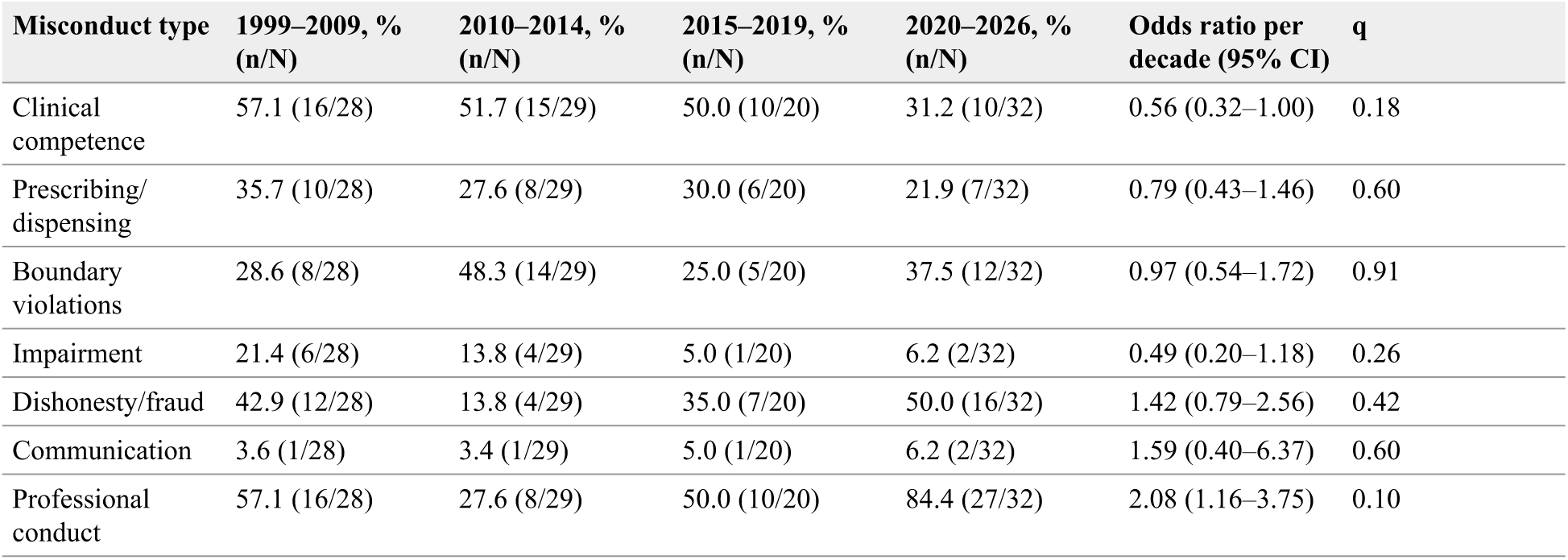
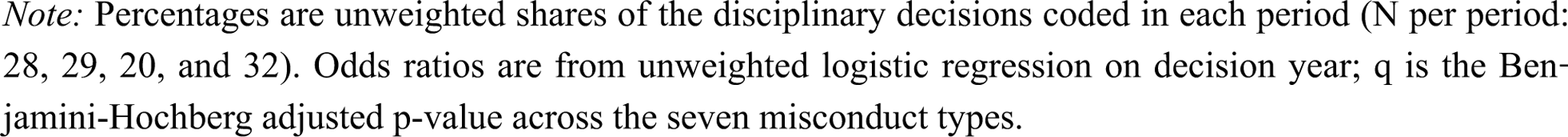
Misconduct types among disciplinary decisions in the coded sample by period, with the odds ratio per decade from logistic regression on decision year

| **Misconduct type** | **1999–2009, % (n/N)** | **2010–2014, % (n/N)** | **2015–2019, % (n/N)** | **2020–2026, % (n/N)** | **Odds ratio per decade (95% CI)** | **q** |
|---|---|---|---|---|---|---|
| Clinical competence | 57.1 (16/28) | 51.7 (15/29) | 50.0 (10/20) | 31.2 (10/32) | 0.56 (0.32–1.00) | 0.18 |
| Prescribing/dispensing | 35.7 (10/28) | 27.6 (8/29) | 30.0 (6/20) | 21.9 (7/32) | 0.79 (0.43–1.46) | 0.60 |
| Boundary violations | 28.6 (8/28) | 48.3 (14/29) | 25.0 (5/20) | 37.5 (12/32) | 0.97 (0.54–1.72) | 0.91 |
| Impairment | 21.4 (6/28) | 13.8 (4/29) | 5.0 (1/20) | 6.2 (2/32) | 0.49 (0.20–1.18) | 0.26 |
| Dishonesty/fraud | 42.9 (12/28) | 13.8 (4/29) | 35.0 (7/20) | 50.0 (16/32) | 1.42 (0.79–2.56) | 0.42 |
| Communication | 3.6 (1/28) | 3.4 (1/29) | 5.0 (1/20) | 6.2 (2/32) | 1.59 (0.40–6.37) | 0.60 |
| Professional conduct | 57.1 (16/28) | 27.6 (8/29) | 50.0 (10/20) | 84.4 (27/32) | 2.08 (1.16–3.75) | 0.10 |

*Note:* Percentages are unweighted shares of the disciplinary decisions coded in each period (N per period: 28, 29, 20, and 32). Odds ratios are from unweighted logistic regression on decision year; q is the Ben-jamini-Hochberg adjusted p-value across the seven misconduct types.

The number of disciplinary decisions increased strongly across complete years from 1999 to 2025 (Mann-Kendall tau = 0.877, FDR-adjusted *p* < 0.001; Sen’s slope 10.5 decisions per year). This growth outpaced the registered workforce (Figure 3; Table 5). Across the ten professions regulated since 2010, the rate rose from 1.25 disciplinary decisions per 10,000 registrants in 2011–2013 to 2.36 in 2023–2025, an increase of about 5% a year (rate ratio 1.050 per year, 95% CI 1.029–1.071). Rates rose for medical practitioners (4.54 to 6.34 per 10,000; rate ratio 1.027, 1.004–1.051) and most steeply for nurses and midwives (0.39 to 1.38; 1.100, 1.063–1.137). Chiropractic also rose (rate ratio 1.162, 1.066–1.268), but on only 38 decisions. Rates for pharmacy, psychology, and dental practitioners showed no clear trend. The all-profession result was unchanged when the pandemic sub-register was included (rate ratio 1.049) and stronger when decisions of unknown profession were allocated (1.066) (Supplementary Table S31).

**Figure 3.**
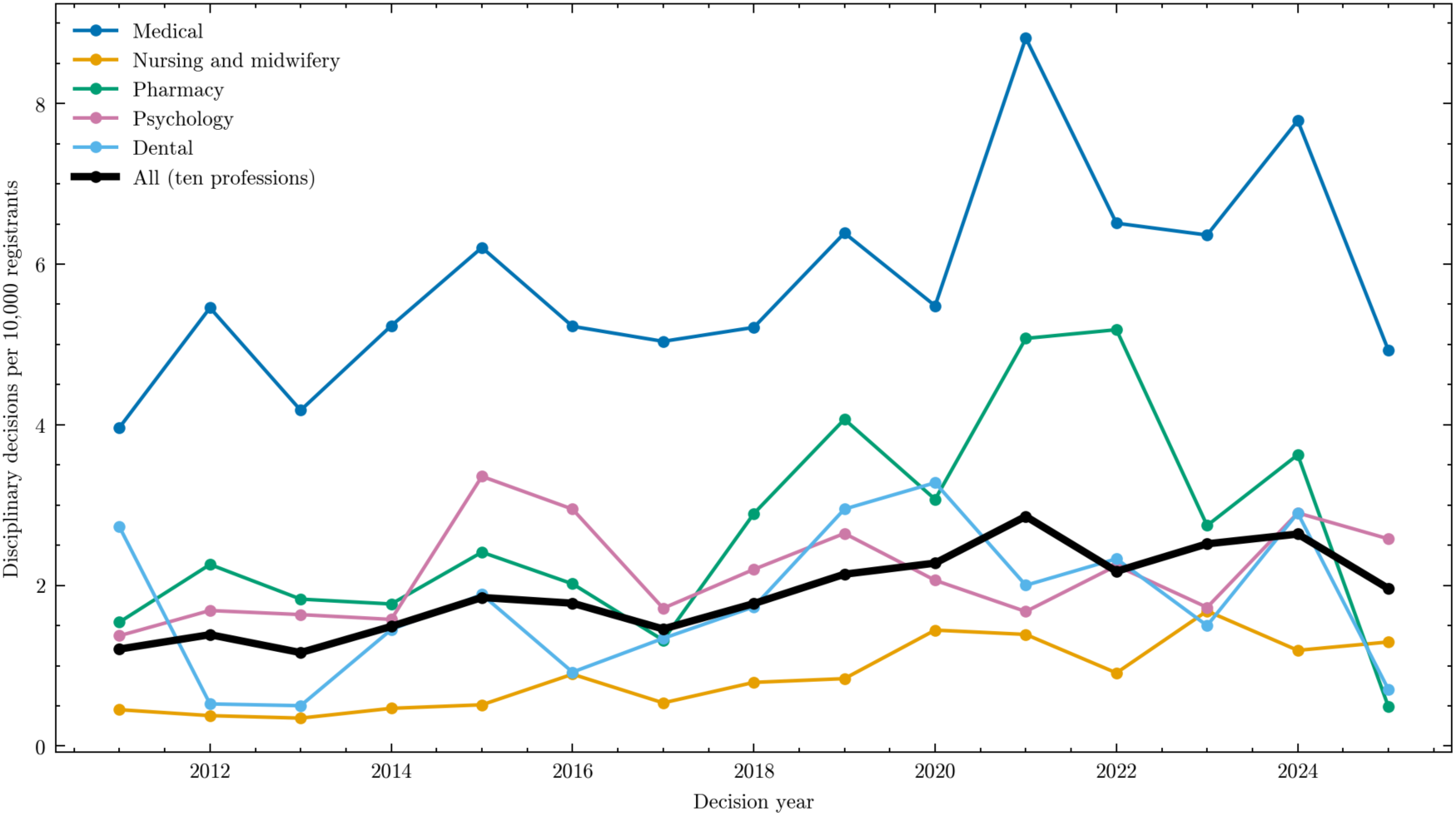
Disciplinary tribunal decisions per 10,000 registered practitioners by profession, 2011–2025 (chiropractic, with 38 decisions, is in Table 5 but not plotted)

**Table 5.**
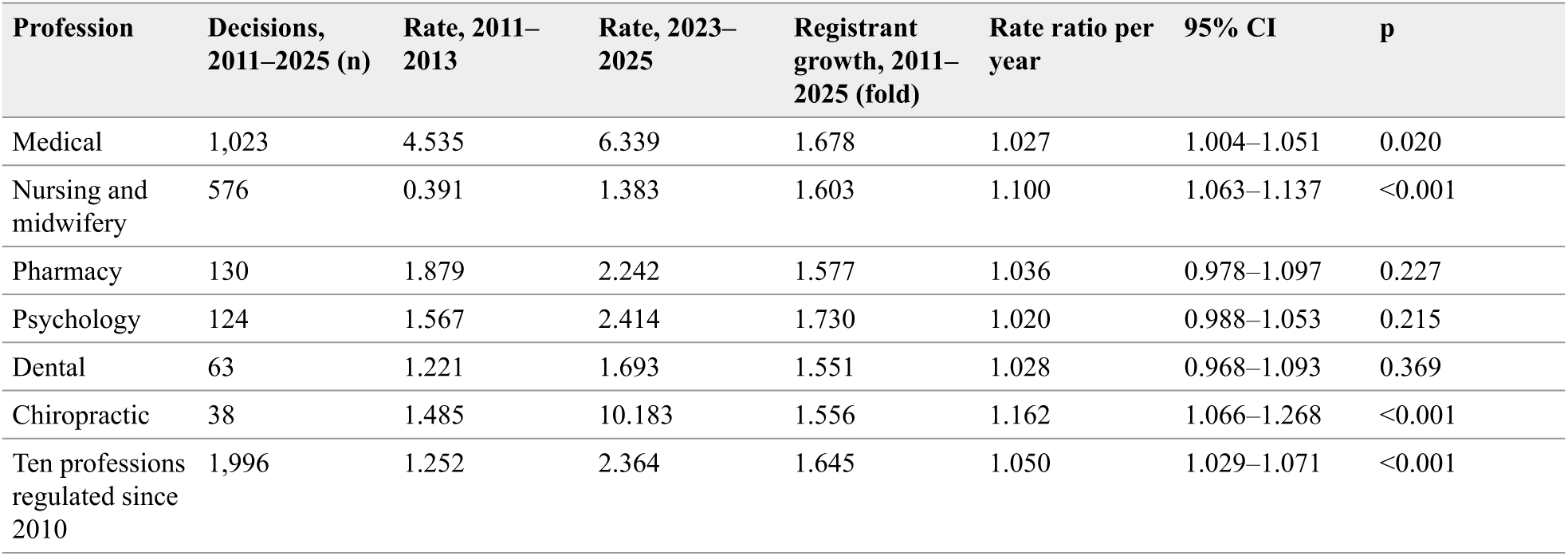
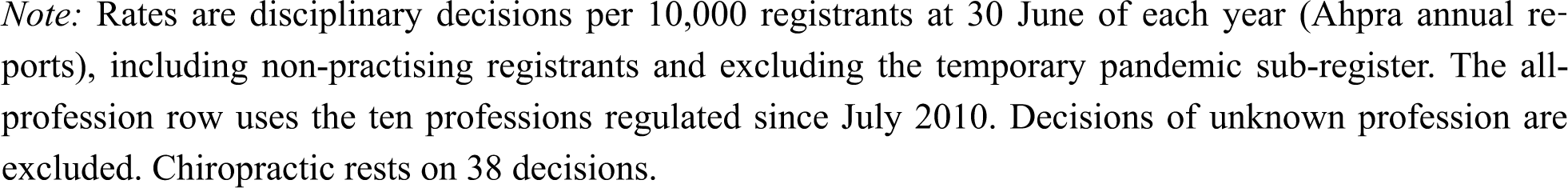
Disciplinary decisions per 10,000 registrants, 2011–2013 and 2023–2025, with registrant growth and rate ratio per year (quasi-Poisson regression)

The classifier’s annual misconduct-type proportions (Supplementary Figure S15) showed significant increases for dishonesty or fraud (tau = 0.462, FDR-adjusted *p* = 0.001) and communication failures (tau = 0.590, *p* < 0.001), and no significant trend for the other categories over the full period. Excluding the years before 2005, when annual counts were small, professional conduct increased (tau = 0.394) and clinical competence decreased (tau = -0.528) (Supplementary Table S24), the same directions as in the coded sample. Neither full-period trend can be called robust under the pre-specified check. For dishonesty or fraud, coded and predicted prevalence moved in the same direction, but the classifier detected 25% of coded fraud matters in each of the two earliest periods and 56% to 57% in the two latest (Figure 4), so part of its rising trend reflects changing error. The communication-failure trend could not be checked (5 coded cases) and rests on a category the classifier labels poorly, so it is not interpretable. A three-period comparison around the COVID-19 pandemic showed shifts in five of seven categories that mirror these longer-term changes and cannot be separated from them (Supplementary Table S11).

**Figure 4.**
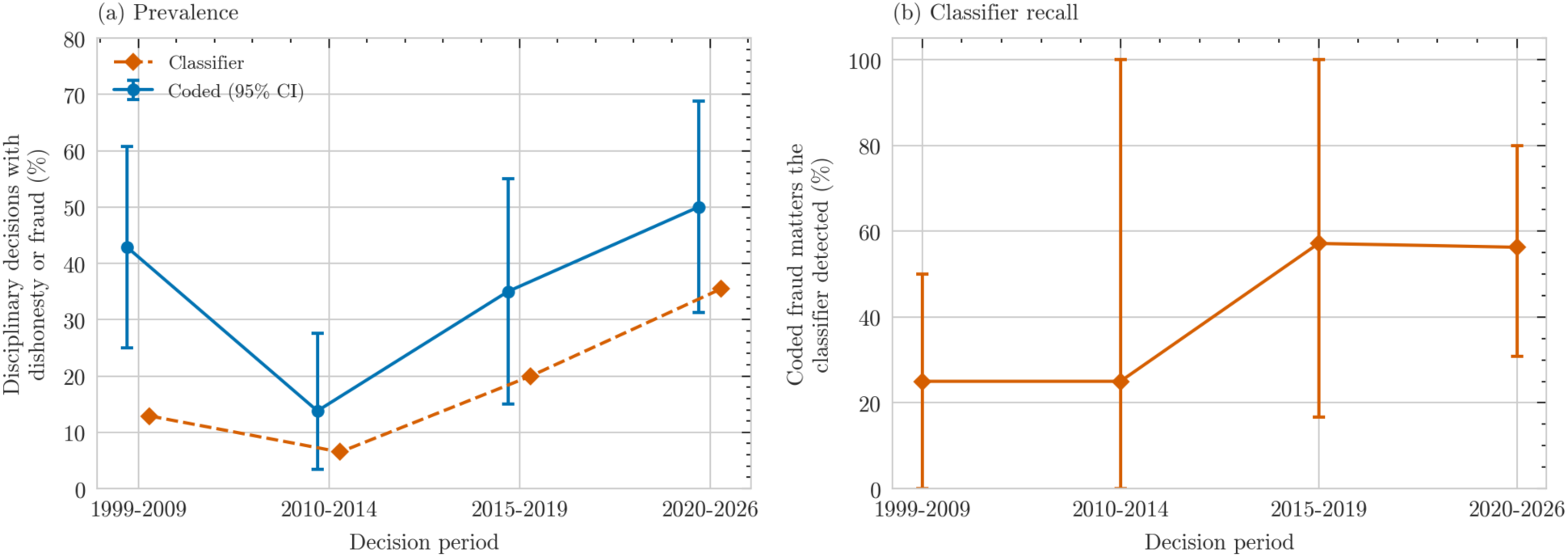
Dishonesty or fraud in the coded sample by period: (a) coded prevalence (95% CI) and classifier-predicted prevalence; (b) share of coded fraud matters the classifier detected (recall, 95% CI)

### E. Exploratory Corpus-Wide Comparisons

The analyses in this section use the automated labels for all 2,477 disciplinary decisions and are exploratory; Table 3 shows how far each can be trusted. Co-occurrence of misconduct types is reported in Supplementary Tables S21 to S23.

Cancellation varied by profession (chi-square = 106.25, df = 9, FDR-adjusted *p* < 0.001, Cramér’s V = 0.209); among professions with more than 100 disciplinary decisions it was highest for pharmacists (55.6%) and lowest for medical practitioners (35.2%), although outcome extraction over-detects cancellation (Table 6). Boundary violations were labelled in 74.4% of psychologist decisions, consistent with Kre-mer’s finding for psychologists.^10^ The most severe outcome varied by jurisdiction (chi-square = 545.01, df = 56, FDR-adjusted *p* < 0.001, V = 0.177), most strongly between Queensland and both New South Wales (V = 0.491) and Victoria (V = 0.440); six of seven misconduct types also varied by jurisdiction, but boundary violations did not (V = 0.058, FDR-adjusted *p* = 0.329) (Supplementary Tables S9 and S10).

**Table 6.**
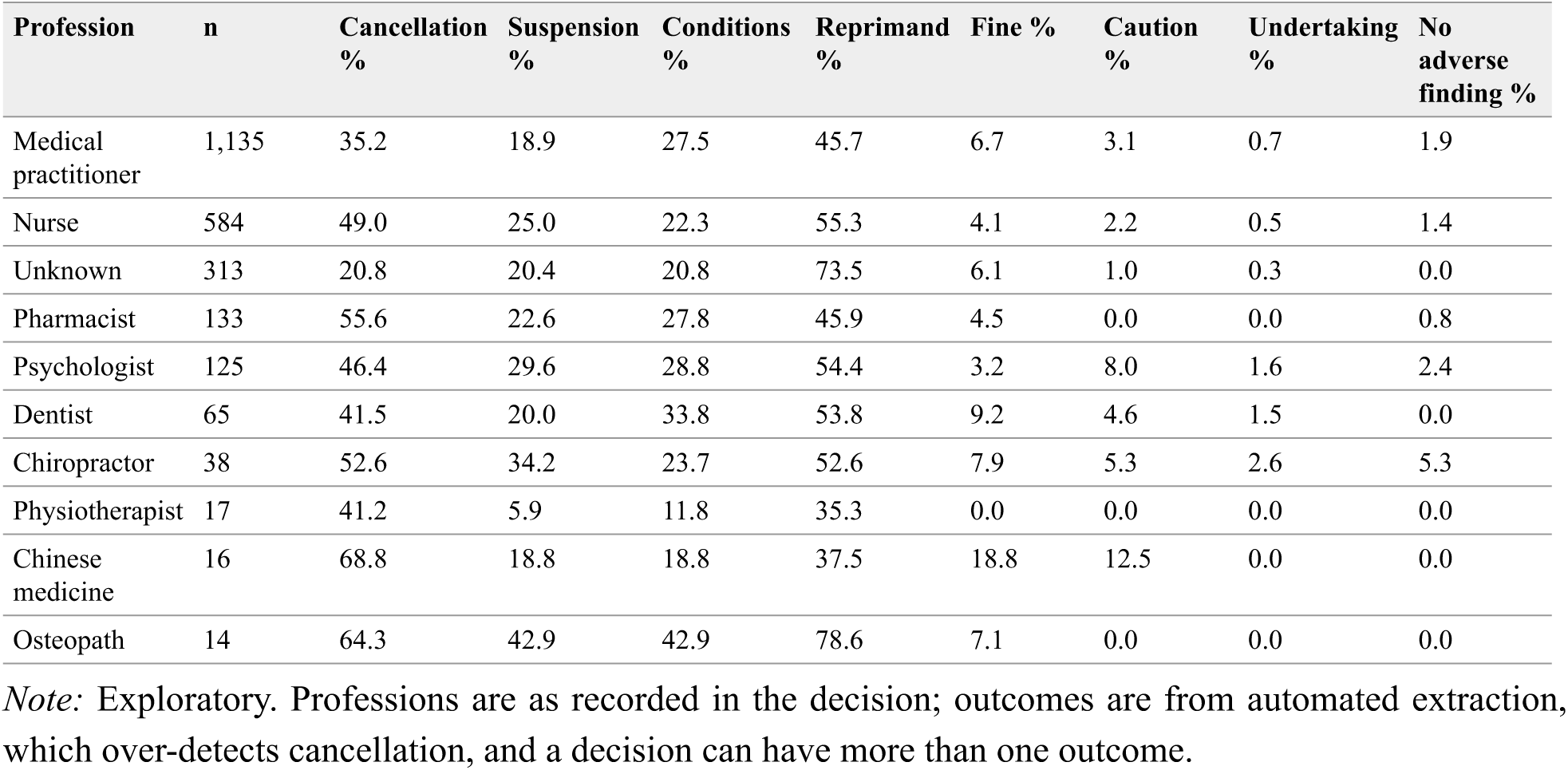
Disciplinary outcome rates by profession, top 10 professions (automated labels; N = 2,440)

Of the 724 decisions labelled as prescribing misconduct, a category the classifier labels well (F1 0.83 on the coded sample), 464 (64.1%) mentioned an opioid, most often oxycodone (31.4%), and of the 15 most frequently mentioned medications, eight are in Schedule 8 of the current Poisons Standard and four are Schedule 4 benzodiazepines subject to Appendix D (Table 7). Mentions include incidental references, so these are upper bounds. Opioid-related prescribing decisions were more often labelled as ending in cancellation than other prescribing decisions (42.0% vs 32.3%; FDR-adjusted *p* = 0.018; Supplementary Table S5). For boundary violations, also labelled well (F1 0.88), the keyword subcategories were accurate for sexual breaches against the coded sample (precision and recall 0.97) but not for financial breaches (precision 0.21), so the coded sample’s 5.5% is the better financial estimate (Supplementary Table S6).

**Table 7.**
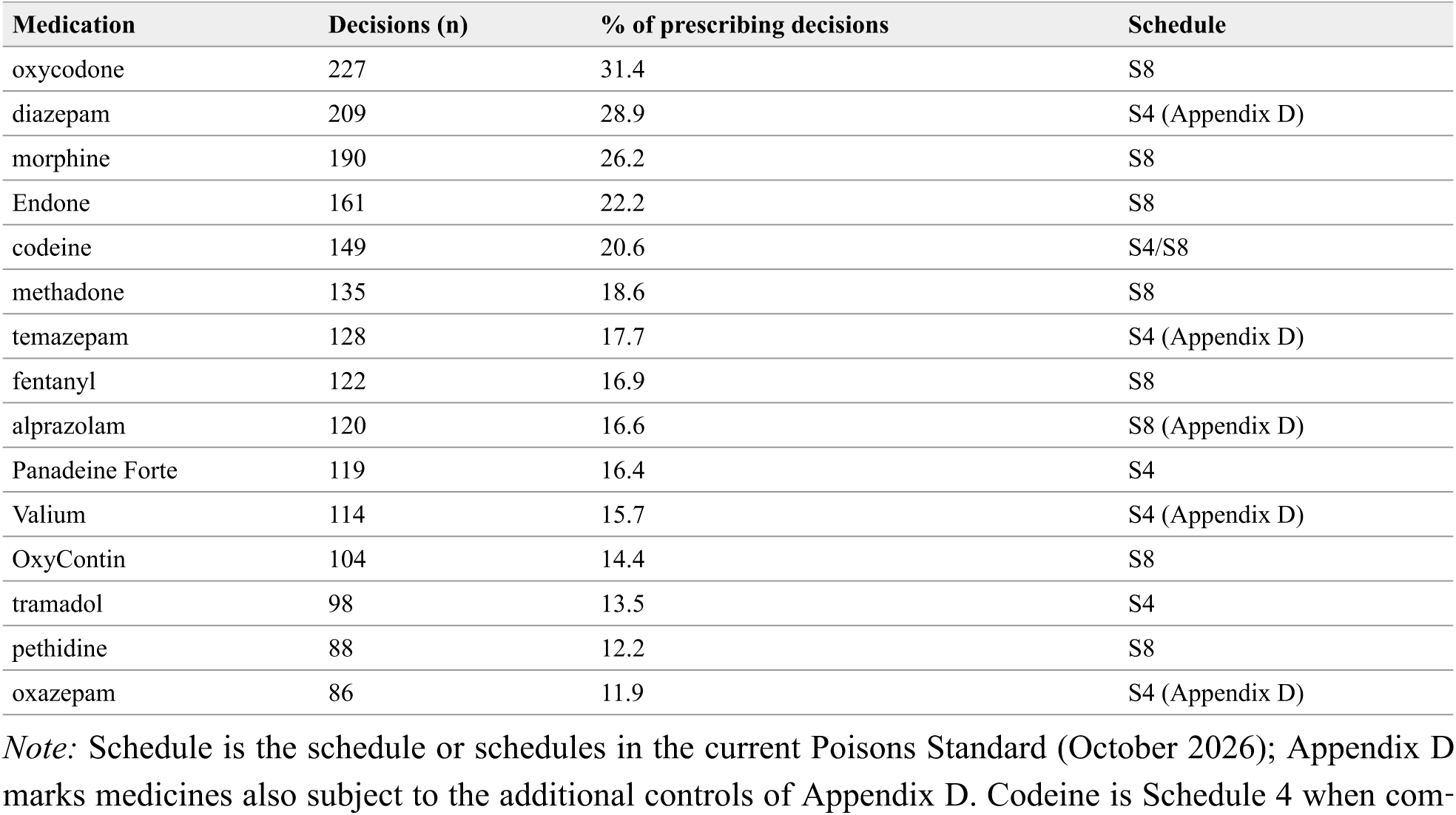
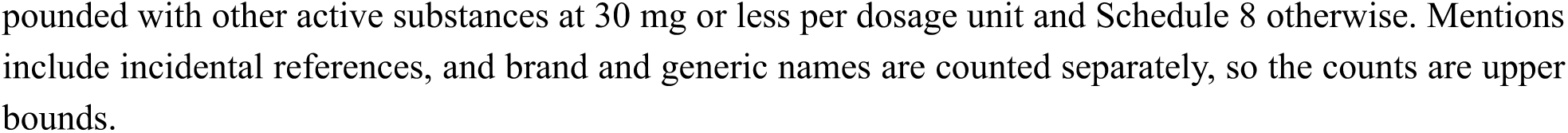
Top 15 medications mentioned in prescribing misconduct decisions (N = 724). Schedule: schedule(s) in the current Poisons Standard (October 2026); App D, also subject to the additional controls of Appendix D. Codeine is Schedule 4 when compounded with other active substances at 30 mg or less per dosage unit and Schedule 8 otherwise.

| Medication | Decisions (n) | % of prescribing decisions | Schedule |
|---|---|---|---|
| oxycodone | 227 | 31.4 | S8 |
| diazepam | 209 | 28.9 | S4 (Appendix D) |
| morphine | 190 | 26.2 | S8 |
| Endone | 161 | 22.2 | S8 |
| codeine | 149 | 20.6 | S4/S8 |
| methadone | 135 | 18.6 | S8 |
| temazepam | 128 | 17.7 | S4 (Appendix D) |
| fentanyl | 122 | 16.9 | S8 |
| alprazolam | 120 | 16.6 | S8 (Appendix D) |
| Panadeine Forte | 119 | 16.4 | S4 |
| Valium | 114 | 15.7 | S4 (Appendix D) |
| OxyContin | 104 | 14.4 | S8 |
| tramadol | 98 | 13.5 | S4 |
| pethidine | 88 | 12.2 | S8 |
| oxazepam | 86 | 11.9 | S4 (Appendix D) |

*Note:* Schedule is the schedule or schedules in the current Poisons Standard (October 2026); Appendix D marks medicines also subject to the additional controls of Appendix D. Codeine is Schedule 4 when com-pounded with other active substances at 30 mg or less per dosage unit and Schedule 8 otherwise. Mentions include incidental references, and brand and generic names are counted separately, so the counts are upper bounds.

## IV. Discussion

### Principal Findings

This study gives the first estimates of the make-up of Australian health practitioner tribunal discipline that are representative of the published decisions across all professions, jurisdictions, and a 27-year period. Breaches of professional conduct, a broad category that includes breaches of registration conditions and conduct outside practice, were found in more than two-thirds of disciplinary decisions in the author’s coding (an independent coder applied this residual category less often), and dishonesty or fraud in about two in five; clinical competence failures and boundary violations each featured in more than a third. Professional misconduct, the most serious finding, was made in 83% of disciplinary decisions, consistent with the referral threshold that directs the most serious matters to tribunals. Professional conduct breaches appear to have become more common over time, and clinical competence failures less common, although neither change is certain. Disciplinary decisions grew faster than the registered workforce, roughly doubling per registrant between 2011–2013 and 2023–2025. Automated labels understated dishonesty and professional conduct, and for dishonesty their error changed over time.

### Dishonesty and Professional Conduct

The prominence of dishonesty is the largest difference between the coded sample and the automated labels: the classifier would have put fraud fifth among seven categories at 23%, where the coded sample puts it second at 42%. Dishonesty in these decisions covers false records, misleading statements to employers or regulators, misrepresented qualifications, and fraudulent billing. It matters for outcome as well as frequency: decision-makers in UK healthcare regulation regard dishonesty, like sexual misconduct, as less remediable than clinical failings,^38^ so its share bears directly on how often removal from practice is likely. The independent second coder found dishonesty slightly more often than the author on the reliability subset (37.0% against 29.3% of disciplinary decisions, a difference within chance), so its prominence does not rest on the author’s coding alone. Dishonesty was common in both the earliest and the latest periods of the coded sample, and the sample shows no clear trend. The classifier’s rising fraud trend is at least partly an artefact of the classifier’s improving detection of fraud in later decisions.

The rise in professional conduct breaches is better supported than the fraud trend but not established: their share in the coded sample rose from 57% of disciplinary decisions before 2010 to 84% from 2020, a change not significant once seven misconduct types are tested, and the automated labels agree in direction once the sparse early years are excluded. The rise may reflect more matters about compliance with earlier regulatory action and about conduct beyond clinical care, as well as changes in how such conduct is charged; the 2022 paramount principle commenced too late in the corpus to account for it.^6^

Professional conduct is also the category on which the two coders agreed least. On the reliability subset the second coder found it in 37% of disciplinary decisions rather than 59%. In 6 of the 9 decisions where only the author coded it, the conduct was dishonest, and the author coded it as professional conduct as well as fraud. As a broad residual category, its prevalence depends on how widely it is applied, and the 68.7% estimate reflects the author’s reading of the guidelines.

### Clinical Competence and Impairment

Clinical competence failures fell from more than half of disciplinary decisions before 2015 to under a third from 2020, although the evidence is at the margin of statistical significance. More clinical performance concerns may now be managed by National Boards through conditions or performance assessment; the 2022 discretion not to refer matters to a tribunal is an unlikely explanation, since the 2025 review found it had not significantly affected referrals.^8^ Impairment featured in 7.5% of disciplinary decisions. About 1% of doctors outside New South Wales were the subject of a health impairment notification over a decade,^39^ but the National Law’s health assessment and Board action pathways may keep most such concerns from tribunals,^40^ and the 2020 narrowing of treating practitioners’ mandatory notification duties for practition-ers’ own health may have reduced them further.^41^

### Boundary Violations

Boundary violations featured in more than a third of disciplinary decisions, and sexual breaches in 84% of these. This accords with Kremer’s finding that boundary violations dominate psychologist decisions,^10^ and with Millbank’s finding that sexual misconduct made up one in five serious tribunal matters under the National Law and attracted the most severe outcomes.^42^ Upstream, sexual misconduct notifications concerned a small proportion of practitioners (0.2% over six years) but led to sanctions more often than other notifications.^43^ There was no evidence that the share of boundary violations changed over time, either in the coded sample or in the automated labels, despite the introduction of mandatory notification in 2010 and the Medical Board’s sexual boundary guidelines (2011, revised 2018).^44,45^ Because these are shares of tribunal decisions, an increase in the number of boundary matters could be masked by faster growth in other categories. In the author’s coding, emotional boundary breaches accompanied most sexual ones, consistent with the ‘slippery slope’ account, in which sexual exploitation of patients often follows a series of earlier non-sexual boundary crossings.^46^ The independent second coder, however, recorded an emotional breach in only 1 of the 14 boundary decisions both coders identified, against 11 for the author. The emotional share therefore depends on how broadly the subtype is read and should be treated with caution.

In the exploratory corpus-wide analysis, decisions labelled as boundary violations were not significantly more likely to end in cancellation than other disciplinary decisions (42.0% vs 38.7%; FDR-adjusted *p* = 0.164; Supplementary Table S7), unlike Millbank’s finding that proved sexual misconduct attracted the highest rate of removal from practice of any head of misconduct;^47^ but the boundary category here includes non-sexual breaches, the comparison decisions also often end in cancellation, and the outcome labels over-detect cancellation. Since April 2026, a finding of professional misconduct with a sexual-misconduct basis is recorded permanently on the public register, whatever the sanction and retrospectively to decisions made from July 2010 (the first round of publication covered 107 practitioners).^7^ A non-cancelling outcome in a sexual misconduct matter therefore no longer leaves a lighter public record, which may alter how such matters are charged and contested, and future analyses will need to treat 2026 as a break.

### Prescribing Misconduct

Prescribing misconduct featured in about a quarter of disciplinary decisions, and almost two-thirds of the decisions labelled as prescribing misconduct mentioned an opioid, which bears on Australia’s efforts to reduce pharmaceutical opioid-related harm^48^ and parallels licensing-board discipline for opioid prescribing in the United States.^49^ Benzodiazepines were also prominent, consistent with their contribution to overdose deaths in Australia and the rescheduling of alprazolam to Schedule 8 in 2014.^50^ The share of prescribing misconduct showed no evidence of change over time, in either the coded sample or the automated labels, despite real-time prescription monitoring (SafeScript in Victoria and the national system) and the rescheduling of codeine to prescription-only status in 2018.^51,52,53^

### Growth Relative to the Workforce

Across the ten professions regulated since 2010, the rate of disciplinary decisions per registrant roughly doubled between 2011–2013 and 2023–2025, and it rose more than threefold for nurses and midwives. The growth may reflect more notifications reaching tribunals, more complete publication of decisions over time, more procedural and appeal decisions per matter, or changes in practitioner conduct, and counts of decisions cannot tell these apart; the denominator also includes non-practising registrants (about 2% to 3% of the total). Whatever its cause, it has regulatory consequences: the 2025 review reported that average investigation times rose by about half between 2018–19 and 2022–23.^8^

### Jurisdictional Variation

The exploratory analysis found large differences in outcomes between jurisdictions, most strongly between Queensland and both New South Wales and Victoria. They may reflect co-regulatory structures (New South Wales has a separate Health Care Complaints Commission, and Queensland a Health Om-budsman),^54^ the mix of matters referred, tribunal composition and practice, and pre-National Law frameworks, as well as genuine differences in how similar cases are treated; separating them would require coding of case severity. The 2025 review recommended longitudinal research to establish whether materially similar cases are treated differently between jurisdictions, and Health Ministers have asked Ahpra to research outcomes for 2020 to 2025.^8^ A combined design like this one, with automated labels for the corpus, representative human coding to measure their error, and coding of case severity, could support that work.

### What Automated Analysis Can and Cannot Show

The study also shows how automated analysis of a legal corpus can go wrong in ways a conventional test set does not reveal. The classifier performed almost the same on its original test set and on the representative sample (macro-F1 0.72 and 0.70), and it identified disciplinary decisions accurately. Yet its corpus-wide estimates understated the second most common misconduct type by almost half, overstated a rare one more than twofold, and produced a rising trend that is partly a product of its own changing error. None of these problems is visible in an overall F1 score. The mechanisms are known in principle: errors in automated labels bias the statistics computed from them even when a classifier scores well on F1,^55^ and a clas-sifier’s error changes when the data it labels differ from the data it was trained on, as decisions written in different periods do.^56^ What this study adds is the size of those effects in a real regulatory corpus, and a practical check that exposes them. The problems become visible only when automated labels are compared with human coding on a sample that represents the corpus and spans its time period. Studies that use machine-learning or language-model labels to describe legal or regulatory corpora, including recent work on health-related court decisions and complaint reports,^19^ should report category-level error on a representative sample and test whether error changes over time before reporting trends. The point is not specific to Australia. Disciplinary actions by United States licensing boards are public records that researchers already review case by case, for example to study discipline for opioid prescribing,^49^ and recent guidance on automated methods in legal epidemiology identifies validation as a central challenge.^21^ A representative, period-stratified validation sample is a practical way to meet it. The human reference needs its own check. Here an independent coder agreed closely with the author on disciplinary status, boundary violations, and prescribing, but only moderately on dishonesty and professional conduct, so the classifier’s error for those two categories is measured against a less certain standard.

A language model, used as a coder with a fixed and logged prompt, performed better than the classifier, particularly for dishonesty (F1 0.85 against 0.66), and its detection of fraud did not drift. Its advantage was not large enough, on 160 decisions, to meet the pre-specified threshold for replacing human coding, and agreement with one author’s coding measures fidelity to the coding guidelines rather than legal correctness. Language-model coders should face the same test as classifiers: category-level error on a representative sample, checked across time.

### Limitations

The main threat to the conclusions is the size and coding of the representative sample. The primary estimates come from 160 decisions coded by one person, so the confidence intervals are wide, rare categories (impairment and communication failures) are imprecise, and only large changes over time can be detected. An independent second coder recoded 60 of the 160 decisions. Agreement was good for disciplinary status, the finding, profession, the main orders, and most misconduct types. It was lower for dishonesty or fraud and professional conduct, the categories that turn most on judgement about breadth, and the two coders applied the emotional boundary subtype very differently. The estimates for professional conduct and for the boundary subtypes therefore depend partly on the author’s interpretation of the guidelines. The second coder spent a median of under two minutes per decision, below the four minutes required at screening. In an exploratory comparison, however, agreement was higher in the faster second session than in the first (Supplementary Appendix H), so the pace does not appear to have degraded the codes. Most disagreements were definitional or about the scope of a decision, which points to rules that could be written more precisely, for example on procedural decisions, where the guidelines’ edge-case rule and the field rule differ; the author applied the field rule throughout, so the primary estimates use one definition. The estimates describe the sampling pool, which excludes the 600 decisions used for the gold standard and for training; those were drawn by profession and jurisdiction, so their exclusion is unlikely to bias the estimates materially.

The sample was also coded twice, and the recode, a departure from the protocol, was made with knowledge of the aggregate results of the first scoring, although without sight of any individual prediction (Supplementary Appendix G).

The corpus-wide analyses are exploratory and, apart from the counts and rates, rest on automated labels with the error rates in Table 3. Associations between groups may be biased by error that differs between them (the classifier’s error by profession on the first test set varied widely, and its error by jurisdiction has not been measured), and outcome extraction over-detects cancellation. The gold standard was checked against the same language model that labelled 300 of the 510 training decisions, so training labels and the first test reference partly share a source, but performance against the author’s blind codes alone was essentially unchanged (macro-F1 0.721 against 0.719; Supplementary Table S26), and the coded sample has no model input.

The corpus may be incomplete. Six legacy state tribunals were unrecoverable, smaller jurisdictions are un-derrepresented, and the corpus includes only published decisions. Matters resolved by consent, surrender of registration, or other non-adjudicated pathways are not included, and tribunals may publish selectively. The findings describe published tribunal decisions, the most serious end of regulatory action, and cannot be generalised to all practitioner misconduct. The unit of analysis is the decision, not the matter: one matter may produce several decisions (for example, liability, sanction, and appeal), which affects comparison with case-based studies such as Millbank’s.

Rates per registrant use annual registrant counts that include non-practising registrants, and depend on the profession recorded in each decision, which was unknown for 311 decisions in 2011–2025. This is a descriptive study, and no causal inference should be drawn from the associations reported.

### Future Directions

Three extensions would strengthen this work. Double coding of the whole representative sample, under guidelines revised where this study’s coders diverged (procedural decisions, the professional conduct category, and the emotional boundary subtype), would make the primary estimates less dependent on one coder’s reading. A larger representative sample, or coding targeted at rare categories, would narrow the intervals and allow change in boundary violations, impairment, and communication failures to be tested. Coding of case severity, representation, and attendance, which Surgenor and colleagues found important in New Zealand,^13^ would allow jurisdictional comparisons that address case mix. Corpus-wide monitoring with automated labels, whether from classifiers or language models, is worth pursuing only with periodic re-validation against representative human coding, especially after legal changes such as the 2026 register reforms.

## V. Conclusion

Across 3,586 Australian tribunal decisions from December 1999 to February 2026, a representative coded sample shows that tribunal discipline is made up largely of breaches of professional conduct and dishonesty, as well as clinical failings and boundary violations (although the professional conduct share depends on how widely that broad category is applied), that most decisions end in a finding of professional misconduct, and that about one in three ends in cancellation. Professional conduct matters appear to have become more common and clinical competence matters less so, and disciplinary decisions have grown much faster than the registered workforce. Automated classification reproduced some of these findings and distorted others, understating dishonesty and producing a fraud trend partly through its own changing error. Published decisions can supply the research on tribunal outcomes over time that the 2025 review asked for, provided that automated labels are checked against representative human coding across the whole period studied.

## Data Availability

All data produced in the present study are available publicly, and instructions to obtain it are contained in the public repository at https://github.com/hayden-farquhar/Health-Practitioner-Misconduct-NLP.

## Acknowledgements

The author gratefully acknowledges the Australasian Legal Information Institute (AustLII) for maintaining open access to Australian legal materials, including the tribunal decisions that form the corpus for this study. The author also acknowledges the Open Australian Legal Corpus hosted on HuggingFace, curated by Umar Butler (Isaacus), which provided New South Wales decisions used in this analysis. The Internet Archive’s Wayback Machine enabled the recovery of decisions from legacy tribunals no longer available on AustLII. This research would not have been possible without these open-access legal information resources. The author thanks A.M., who independently coded the reliability subset as a paid coder recruited through Prolific.

## Ethics Statement

This study analysed publicly available legal decisions published by Australian tribunals. No ethics approval was required. Results are reported in aggregate, and no practitioner is named. The independent second coder was engaged as a paid contributor providing a coding service, not as a research participant; no personal data were collected beyond a pseudonymous platform identifier, which is not reported, and the coder agreed to be acknowledged by initials.

## Data and Code Availability

Analysis code and configuration files are publicly available at https://github.com/hayden-farquhar/Health-Practitioner-Misconduct-NLP. The raw tribunal decisions are publicly available through AustLII and the Open Australian Legal Corpus. The scraped corpus is not included due to size but is reproducible from the pipeline.

## Author Contributions

HF conceived the study, developed the methodology, coded the decisions, recruited and paid the independent second coder, wrote all code, performed all analyses, and wrote the manuscript (sole author). CRediT: Conceptualisation, Methodology, Software, Formal analysis, Investigation, Data curation, Writing – original draft, Writing – review and editing, Visualisation.

## AI Use Disclosure

Claude Sonnet 5.5 (Anthropic; model identifier claude-sonnet-5-5) was used in three logged batch runs with the same fixed prompt: as the second coder of the 300 gold-standard decisions (Methods section B), to label 300 further decisions used as supplementary training data (Methods section D), and once on the 160-decision coded sample to test whether it should replace the classifier (Methods section E). Every request and response was logged, and no model output was edited. All human coding was done by the author, blind to the model’s codes, except the 60-decision reliability subset, which was also coded by an independent paid coder who attested to using no AI tools; every disagreement between the author and the model in the gold standard was adjudicated by the author; the coded sample has no model input. Claude Code (Anthropic) assisted with code development, reference checking, data checks (including the checks that identified the misaligned first coding of the coded sample), and manuscript drafting and editing. It also staged the second coder’s Prolific studies and workbooks, froze the returned files, computed agreement from them, and drafted the descriptive classification of the coders’ disagreements by type (Supplementary Table S35), which the author reviewed; it did not create or change any coder’s codes. No Claude Code output was used as data or as a reference standard. The classifiers themselves are TF-IDF logistic regression models, not language models. The author decided the study design, the author’s coding of every decision, and every adjudication, reviewed and verified all content, and takes responsibility for it.

## Notes

1. *Health Practitioner Regulation National Law Act 2009* (Qld). The National Law operates as a schedule to this Act, applied in each state and territory through corresponding legislation.

2. The 16 regulated professions are: Aboriginal and Torres Strait Islander health practice, Chinese medicine, chiropractic, dental, medical, medical radiation practice, midwifery, nursing, occupational therapy, optometry, osteopathy, paramedicine, pharmacy, physiotherapy, podiatry, and psychology. See Ahpra, *Annual Report 2023/24* (2024).

3. Ahpra and National Boards, *Annual Report 2023/24* (2024) ‘Introduction’ https://www.ahpra.gov.au/Publications/Annual-re-ports/Annual-report-2024/Introduction; Professional Standards Authority for Health and Social Care, *Our Work with Regulators* (Web Page) https://www.professionalstandards.org.uk/organisations-we-oversee/our-work-regulators, accessed 3 November 2026.

4. *Health Practitioner Regulation National Law Act 2009* (Qld) sch pt 8 div 12, s 193 (referral of matters to responsible tribunals).

5. Precision is the share of a classifier’s labels for a misconduct type that the human coder also gave; recall (sensitivity) is the share of the coder’s labels that the classifier also found. F1 combines the two into one score between 0 and 1 (1 is perfect agreement), and macro-F1 averages it across categories. Cohen’s kappa measures agreement between two coders beyond chance (1 is perfect agreement, 0 is chance).

6. *Health Practitioner Regulation National Law and Other Legislation Amendment Act 2022* (Qld) ch 3 pts 2, 26, inserting National Law ss 3A (paramount principle, commenced 21 October 2022) and 193A (decision not to refer to a tribunal, commenced 15 May 2023).

7. *Health Practitioner Regulation National Law and Other Legislation Amendment Act 2025* (Qld) ss 17–21, inserting National Law ss 77A, 198A–198E (reinstatement orders) and 225A–225B (recording of professional misconduct findings based on sexual misconduct), commenced 10 April 2026; Medical Board of Australia, ‘Health Practitioner Register Updated under Landmark Re-forms’ (News, 10 April 2026) https://www.medicalboard.gov.au/News/2026-04-10-Register-updated-under-landmark-re-forms.aspx, accessed 6 November 2026. On reinstatement before these reforms, see Jenni Millbank, ‘Reinstatement of Previously Deregistered Health Professionals in Australia: Legal Determinations of Risk, Patient Safety, and Public Interest’ (2023) 51(1) *Federal Law Review* 3 https://doi.org/10.1177/0067205X221146334.

8. Sue Dawson, *Transforming Health Professions Regulation in Australia: Independent Review Final Report* (Independent Review of Complexity in the National Registration and Accreditation Scheme, July 2025) 72, 84–5 https://www.health.gov.au/sites/default/files/2025-09/transforming-health-professionals-regulation-in-australia-independent-review-final-report.pdf, accessed 6 November 2026; Health Ministers’ Meeting, *Response to the Independent Review of Complexity in the National Registration and Accreditation Scheme* (1 May 2026) action 4.7 https://www.health.gov.au/sites/default/files/2026-05/health-ministers-meeting-re-sponse-to-the-independent-review-of-complexity-in-the-national-registration-and-accreditation-scheme_0.pdf, accessed 6 November 2026.

9. Jenni Millbank, ‘Serious Misconduct of Health Professionals in Disciplinary Tribunals under the National Law 2010–17’ (2020) 44(2) *Australian Health Review* 190 https://doi.org/10.1071/AH18239; see also Jenni Millbank, ‘Health Practitioner Regulation: Has the National Law Produced National Outcomes in Serious Disciplinary Matters?’ (2019) 47(4) *Federal Law Review* 631 https://doi.org/10.1177/0067205X19875028.

10. Paul D Kremer, ‘The Path to Psychologist Misconduct: A Thematic Analysis of Australian Tribunal Decisions (2016–2025)’ (2026) *Australian Psychologist* (advance online publication, 17 September 2026) 1 https://doi.org/10.1080/00050067.2026.2733201.

11. Matthew J Spittal, Marie M Bismark and David M Studdert, ‘Identification of Practitioners at High Risk of Complaints to Health Profession Regulators’ (2019) 19 *BMC Health Services Research* 380 https://doi.org/10.1186/s12913-019-4214-y; Marie M Bismark et al, ‘Identification of Doctors at Risk of Recurrent Complaints: A National Study of Healthcare Complaints in Aus-tralia’ (2013) 22(7) *BMJ Quality & Safety* 532 https://doi.org/10.1136/bmjqs-2012-001691; Matthew J Spittal et al, ‘Outcomes of Notifications to Health Practitioner Boards: A Retrospective Cohort Study’ (2016) 14 *BMC Medicine* 198 https://doi.org/10.1186/s12916-016-0748-6; Marie M Bismark et al, ‘Mandatory Reports of Concerns about the Health, Performance and Conduct of Health Practitioners’ (2014) 201(7) *Medical Journal of Australia* 399 https://doi.org/10.5694/mja14.00210; Katie Elkin, Matthew J Spittal and David M Studdert, ‘Risks of Complaints and Adverse Disciplinary Findings against International Medical Graduates in Victoria and Western Australia’ (2012) 197(8) *Medical Journal of Australia* 448 https://doi.org/10.5694/mja12.10632.

12. Katie J Elkin et al, ‘Doctors Disciplined for Professional Misconduct in Australia and New Zealand, 2000–2009’ (2011) 194(9) *Medical Journal of Australia* 452 https://doi.org/10.5694/j.1326-5377.2011.tb03058.x; see Millbank (n 9) 192 (adapting the Elkin typology).

13. Lois J Surgenor et al, ‘Fifteen Years On: What Patterns Continue to Emerge from New Zealand’s Health Practitioners Disciplinary Tribunal?’ (2020) 28(1) *Journal of Law and Medicine* 165; Lois Surgenor et al, ‘To Impose or Not Impose Penalty Conditions Following Professional Misconduct: What Factors Are Cited by Three Professional Disciplinary Tribunals in New Zealand?’ (2024) 13(6) *Laws* 69 https://doi.org/10.3390/laws13060069.

14. Nikolaos Aletras et al, ‘Predicting Judicial Decisions of the European Court of Human Rights: A Natural Language Processing Perspective’ (2016) 2 *PeerJ Computer Science* e93 https://doi.org/10.7717/peerj-cs.93.

15. Ilias Chalkidis et al, ‘LEGAL-BERT: The Muppets Straight Out of Law School’ in *Findings of the Association for Computational Linguistics: EMNLP 2020* (Association for Computational Linguistics, 2020) 2898 https://doi.org/10.18653/v1/2020.find-ings-emnlp.261.

16. Masha Medvedeva, Michel Vols and Martijn Wieling, ‘Using Machine Learning to Predict Decisions of the European Court of Human Rights’ (2020) 28(2) *Artificial Intelligence and Law* 237 https://doi.org/10.1007/s10506-019-09255-y.

17. See, eg, Jacob Devlin et al, ‘BERT: Pre-Training of Deep Bidirectional Transformers for Language Understanding’ in *Proceedings of the 2019 Conference of the North American Chapter of the Association for Computational Linguistics: Human Language Technologies* (Association for Computational Linguistics, 2019) vol 1, 4171 https://doi.org/10.18653/v1/N19-1423; Haoxi Zhong et al, ‘How Does NLP Benefit Legal System: A Summary of Legal Artificial Intelligence’ in *Proceedings of the 58th Annual Meeting of the Association for Computational Linguistics* (Association for Computational Linguistics, 2020) 5218 https://doi.org/10.18653/v1/2020.acl-main.466; Ilias Chalkidis et al, ‘LexGLUE: A Benchmark Dataset for Legal Language Understanding in English’ in *Proceedings of the 60th Annual Meeting of the Association for Computational Linguistics* (Association for Computational Linguistics, 2022) vol 1, 4310 https://doi.org/10.18653/v1/2022.acl-long.297.

18. Raphael Souza de Oliveira and Erick Giovani Sperandio Nascimento, ‘Analysing Similarities between Legal Court Documents Using Natural Language Processing Approaches Based on Transformers’ (2025) 20(4) *PLOS ONE* e0320244 https://doi.org/10.1371/journal.pone.0320244.

19. Min Ji Kim and Min Choi, ‘Legal Infoveillance of Unlicensed Medical Practices in South Korea through Criminal Court Decisions Using Machine Learning: Retrospective Observational Study’ (2026) 12 *JMIR Public Health and Surveillance* e92591 ht-tps://doi.org/10.2196/92591; Cameron Wells et al, ‘Learning from Complaints about Surgical Care: A Large Language Model-Assisted Sequential Methods Analysis’ (2026) *BMJ Quality & Safety* (advance online publication, 28 January 2026) https://doi.org/10.1136/bmjqs-2025-019556.

20. Alan Hanna and Lezley-Anne Hanna, ‘Topic Analysis of UK Fitness to Practise Cases: What Lessons Can Be Learnt?’ (2019) 7(3) *Pharmacy* 130 https://doi.org/10.3390/pharmacy7030130.

21. Regen Weber-Fares et al, ‘Leveraging Artificial Intelligence and Natural Language Processing in Legal Epidemiology Studies: Opportunities and Challenges’ (2026) 54(3) *Journal of Law, Medicine & Ethics* 368 https://doi.org/10.1017/jme.2025.10224.

22. Erik von Elm et al, ‘The Strengthening the Reporting of Observational Studies in Epidemiology (STROBE) Statement: Guidelines for Reporting Observational Studies’ (2007) 370(9596) *The Lancet* 1453 https://doi.org/10.1016/S0140-6736(07)61602-X; Gary S Collins et al, ‘TRIPOD+AI Statement: Updated Guidance for Reporting Clinical Prediction Models That Use Regression or Machine Learning Methods’ (2024) 385 *BMJ* e078378 https://doi.org/10.1136/bmj-2023-078378.

23. Umar Butler, *Open Australian Legal Corpus* (Dataset, Hugging Face, 2024) https://doi.org/10.57967/hf/2784, accessed 11 February 2026.

24. Albert Weichselbraun, ‘Inscriptis: A Python-Based HTML to Text Conversion Library Optimized for Knowledge Extraction from the Web’ (2021) 6(66) *Journal of Open Source Software* 3557 https://doi.org/10.21105/joss.03557.

25. Ahpra and National Boards, *Annual Report 2023/24* (2024) table 16 (‘Types of concerns raised’) and ‘Notifications’ https://www.ahpra.gov.au/Publications/Annual-reports/Annual-report-2024/Notifications (accessed 4 November 2026); Elkin et al (n 12) box 4; Millbank (n 9) 192.

26. Meysam Alizadeh et al, ‘Open-Source LLMs for Text Annotation: A Practical Guide for Model Setting and Fine-Tuning’ (2025) 8(1) *Journal of Computational Social Science* 17 https://doi.org/10.1007/s42001-024-00345-9; Fabrizio Gilardi, Meysam Alizadeh and Mael Kubli, ‘ChatGPT Outperforms Crowd Workers for Text-Annotation Tasks’ (2023) 120(30) *Proceedings of the National Academy of Sciences* e2305016120 https://doi.org/10.1073/pnas.2305016120.

27. Petter Törnberg, ‘Large Language Models Outperform Expert Coders and Supervised Classifiers at Annotating Political Social Media Messages’ (2025) 43(6) *Social Science Computer Review* 1181 https://doi.org/10.1177/08944393241286471.

28. Key coding rules: boundary violations were reserved for practitioner–patient relationship breaches, with criminal conduct outside practice and breach of registration conditions coded as professional conduct; disqualification of unregistered practitioners was coded as ‘other’, not cancellation; and pharmacist dispensing misconduct was coded as prescribing misconduct. The one post-adjudication amendment added a communication failure that both coders had left uncoded; it was decided by the author and is recorded with its date and reason, with the frozen blind codes, the model’s logged outputs, and the adjudication record. The final gold standard comprised 213 disciplinary and 87 non-disciplinary decisions.

29. Stefan Palan and Christian Schitter, ‘Prolific.ac: A Subject Pool for Online Experiments’ (2018) 17 *Journal of Behavioral and Experimental Finance* 22 https://doi.org/10.1016/j.jbef.2017.12.004.

30. J Richard Landis and Gary G Koch, ‘The Measurement of Observer Agreement for Categorical Data’ (1977) 33(1) *Biometrics* 159 https://doi.org/10.2307/2529310.

31. See, eg, Aletras et al (n 14) (N-gram features for judicial decision prediction); Fabian Pedregosa et al, ‘Scikit-Learn: Machine Learning in Python’ (2011) 12 *Journal of Machine Learning Research* 2825 https://jmlr.org/papers/v12/pedregosa11a.html (the scikit-learn library used for vectorisation and classification).

32. Chalkidis et al (n 15).

33. *Therapeutic Goods (Poisons Standard—October 2026) Instrument 2026* (Cth) Index, app D https://www.legislation.gov.au/F2026L01327/latest/text (accessed 5 November 2026).

34. Maarten Grootendorst, ‘BERTopic: Neural Topic Modeling with a Class-Based TF-IDF Procedure’ (2022) *arXiv preprint* arXiv:2203.05794.

35. Ahpra and National Boards, *Annual Report* (2010/11 to 2025/26), tables of registered practitioners by profession and principal place of practice https://www.ahpra.gov.au/Publications/Annual-reports/Annual-report-archive, accessed 6 November 2026.

36. Walter J Rogan and Beth Gladen, ‘Estimating Prevalence from the Results of a Screening Test’ (1978) 107(1) *American Journal of Epidemiology* 71 https://doi.org/10.1093/oxfordjournals.aje.a112510.

37. A calibrated classifier’s predicted probabilities match observed frequencies. Here only the final labels were used, so the analyses depend on the error rates in Table 3 rather than on calibration.

38. Tristan Price et al, ‘Role of Remediation in Cases of Serious Misconduct before UK Healthcare Regulators: A Qualitative Study’ (2025) 34(2) *BMJ Quality & Safety* 110 https://doi.org/10.1136/bmjqs-2024-017187.

39. Marie M Bismark et al, ‘Health Impairment Notifications about Doctors to the Australian Medical Regulator, 2012–2022: A Retrospective Cohort Study’ (2026) 224(1) *Medical Journal of Australia* e70131 https://doi.org/10.5694/mja2.70131.

40. *Health Practitioner Regulation National Law Act 2009* (Qld) sch pt 8 divs 9 (health and performance assessments), 10 (action by National Board) and 11 (panels).

41. *Health Practitioner Regulation National Law and Other Legislation Amendment Act 2019* (Qld) ss 17–19, inserting National Law ss 141A–141C (mandatory notifications by treating practitioners), commenced 1 March 2020.

42. Millbank (n 9) 194–5; see also Jenni Millbank, ‘Female Health Practitioners Disciplined for Sexual Misconduct’ (2020) 43(4) *University of New South Wales Law Journal* 1244 https://doi.org/10.53637/QZEC5520.

43. Marie M Bismark et al, ‘Sexual Misconduct by Health Professionals in Australia, 2011–2016: A Retrospective Analysis of Notifications to Health Regulators’ (2020) 213(5) *Medical Journal of Australia* 218 https://doi.org/10.5694/mja2.50706.

44. *Health Practitioner Regulation National Law Act 2009* (Qld) sch ss 140–143 (mandatory notifications by practitioners, employers, and education providers).

45. Medical Board of Australia, *Guidelines: Sexual Boundaries in the Doctor–Patient Relationship* (12 December 2018) (replacing guidelines in effect from 28 October 2011) https://www.medicalboard.gov.au/Codes-Guidelines-Policies/Sexual-boundaries-guidelines.aspx.

46. Cherrie A Galletly, ‘Crossing Professional Boundaries in Medicine: The Slippery Slope to Patient Sexual Exploitation’ (2004) 181(7) *Medical Journal of Australia* 380 https://doi.org/10.5694/j.1326-5377.2004.tb06334.x; Lisa Lampe et al, ‘Being a “Good” Doctor: Understanding and Managing Professional Boundaries Is Challenging and Can Lead to Stress and Burnout’ (2023) 31(6) *Australasian Psychiatry* 764 https://doi.org/10.1177/10398562231191662.

47. Millbank (n 9) 194.

48. Australian Institute of Health and Welfare, *Opioid Harm in Australia: And Comparisons between Australia and Canada* (Cat No HSE 210, November 2018) https://www.aihw.gov.au/reports/illicit-use-of-drugs/opioid-harm-in-australia/summary; Penington Institute, *Australia’s Annual Overdose Report 2023* (August 2023) https://www.penington.org.au/wp-content/uploads/2023/08/PEN_Annual-Overdose-Report-2023_FINAL.pdf.

49. Carol L Galletly et al, ‘Out of Bounds: Physician Licensing Board Disciplinary Cases Related to Opioid Prescribing’ (2024) 52(3) *Journal of Law, Medicine & Ethics* 679 https://doi.org/10.1017/jme.2024.138.

50. Penington Institute (n 48) 8; Rachel Sutherland et al, ‘Alprazolam Use among a Sample of Australians Who Inject Drugs: Trends up to Six Years Post Regulatory Changes’ (2020) 79 *International Journal of Drug Policy* 102721 https://doi.org/10.1016/j.drugpo.2020.102721.

51. Department of Health (Vic), *SafeScript* (Web Page) https://www.health.vic.gov.au/drugs-and-poisons/safescript, accessed 3 November 2026.

52. Department of Health, Disability and Ageing (Cth), *National Real Time Prescription Monitoring* (Web Page) https://www.health.gov.au/our-work/national-real-time-prescription-monitoring, accessed 3 November 2026.

53. Penington Institute (n 48); Therapeutic Goods Administration, *Codeine Advice and Support If You Have Chronic Pain* (Web Page, 4 April 2018) https://www.tga.gov.au/resources/explore-topic/prescription-opioids-hub/codeine-advice-and-support-if-you-have-chronic-pain, accessed 17 November 2026 (codeine became prescription-only on 1 February 2018).

54. *Health Care Complaints Act 1993* (NSW); *Health Ombudsman Act 2013* (Qld).

55. Nathan TeBlunthuis, Valerie Hase and Chung-Hong Chan, ‘Misclassification in Automated Content Analysis Causes Bias in Regression. Can We Fix It? Yes We Can!’ (2024) 18(3) *Communication Methods and Measures* 278 https://doi.org/10.1080/19312458.2023.2293713.

56. Jose G Moreno-Torres et al, ‘A Unifying View on Dataset Shift in Classification’ (2012) 45(1) *Pattern Recognition* 521 ht-tps://doi.org/10.1016/j.patcog.2011.06.019.

