## Supplementary Materials for "What Australian Health Practitioner Tribunals Decide, and What Automated Analysis Misses: 3,586 Decisions, 1999 to February 2026"

**Contents.** Appendix A: misconduct taxonomy. Appendix B: keyword filtering. Appendix C: supplementary tables (S1 to S26 use the automated labels for the whole corpus and are exploratory; S27 to S32 report the coded sample). Appendix D: supplementary figures (S1 to S15). Appendix E: classifier technical details. Appendix F: topic model. Appendix G: the coded sample and its re-coding. Appendix H: independent second coder (Tables S33 to S35).

### Appendix A: Full Misconduct Taxonomy

The misconduct taxonomy comprises 7 Level 1 (L1) categories and 34 Level 2 (L2) subcategories. Definitions and examples are provided below. Misconduct categories are applied as multi-label classifications: a single decision may be assigned to multiple categories. Detailed annotation guidelines and coding rules are available as a separate document.

#### A.1 Clinical Competence (CLIN)

**Definition:** Deficiencies in clinical knowledge, skill, judgement, or care delivery that fall below the expected standard for the practitioner's profession and level of experience.

| Code | Subcategory | Definition |
|---|---|---|
| CLIN_DIAG | Diagnostic failure | Failure to diagnose, misdiagnosis, or delayed diagnosis of a condition |
| CLIN_TREAT | Inappropriate treatment | Treatment that is not clinically indicated, is contraindicated, or falls below accepted standards |
| CLIN_ASSESS | Inadequate assessment | Failure to conduct an adequate clinical assessment, history, or examination |
| CLIN_RECORD | Inadequate record-keeping | Clinical records that are absent, incomplete, inaccurate, or do not meet professional standards |
| CLIN_INFCON | Infection control failure | Failure to maintain infection control or hygiene standards |
| CLIN_OTHER | Other clinical competence issue | Clinical competence issues not captured by the above subcategories |

**Inclusion:** Errors or deficiencies directly related to clinical care delivery, diagnosis, treatment planning, or clinical decision-making.

**Exclusion:** Prescribing-specific issues (PRESC). Communication failures during clinical encounters (COMM). Impairment-related clinical errors (code under both CLIN and IMPR).

#### A.2 Prescribing Misconduct (PRESC)

**Definition:** Inappropriate, excessive, or fraudulent prescribing, dispensing, or supply of medications, including failure to monitor patients on medication, prescribing or dispensing without adequate assessment, and self-prescribing or self-supply.

| Code | Subcategory | Definition |
|---|---|---|
| PRESC_OPIOID | Opioid prescribing | Inappropriate prescribing of opioid medications (Schedule 8 or equivalent) |
| PRESC_BENZO | Benzodiazepine prescribing | Inappropriate prescribing of benzodiazepines (Schedule 4 Appendix D or equivalent) |
| PRESC_OTHER_S8 | Other Schedule 8 prescribing | Inappropriate prescribing of Schedule 8 substances other than opioids |
| PRESC_GENERAL | General prescribing failure | Prescribing failures not specific to controlled substances |
| PRESC_SELF | Self-prescribing | Prescribing medication for oneself, family members, or close associates |
| PRESC_MONITOR | Monitoring failure | Failure to appropriately monitor patients on medication |

**Inclusion:** All matters primarily concerning prescribing practices, medication management, or drug supply.

**Exclusion:** Drug-related impairment of the practitioner (IMPR). Clinical decisions about non-pharmacological treatment (CLIN).

#### A.3 Boundary Violations (BOUND)

**Definition:** Breaches of professional boundaries in the practitioner–patient relationship, including sexual, emotional, financial, and social boundary violations.

| Code | Subcategory | Definition |
|---|---|---|
| BOUND_SEXUAL | Sexual boundary violation | Sexual contact, sexual relationship, or sexualised behaviour with a patient or former patient |
| BOUND_EMOTIONAL | Emotional boundary violation | Inappropriate emotional involvement with a patient, including dual relationships and emotional dependency |
| BOUND_FINANCIAL | Financial boundary violation | Inappropriate financial dealings with a patient |
| BOUND_SOCIAL | Social/digital boundary violation | Inappropriate social or online interactions with patients |

**Inclusion:** All matters involving inappropriate relationships or interactions between practitioner and patient/client that breach professional boundaries.

**Exclusion:** Communication failures that do not involve boundary crossing (COMM). Financial fraud not involving patients (FRAUD).

#### A.4 Impairment (IMPR)

**Definition:** The practitioner’s ability to practise is impaired by a physical or mental health condition, substance use disorder, or cognitive impairment.

| Code | Subcategory | Definition |
|---|---|---|
| IMPR_SUBSTANCE | Substance use disorder | Practitioner has a substance use disorder or is practising while affected by alcohol or drugs |
| IMPR_MENTAL | Mental health impairment | Practitioner has a mental health condition that impairs their ability to practise safely |
| IMPR_PHYSICAL | Physical health impairment | Practitioner has a physical health condition or cognitive |

| Code | Subcategory | Definition |
|---|---|---|
| | | impairment affecting their ability to practise |
| IMPR_NONCOMPLIANCE | Non-compliance with health monitoring | Failure to comply with health-related conditions on registration or monitoring programs |

**Inclusion:** All matters where the practitioner’s health or substance use is a primary concern affecting their fitness to practise.

**Exclusion:** Prescribing misconduct not related to the practitioner’s own substance use (PRESC). Dishonesty to obtain drugs for patients (FRAUD).

#### A.5 Dishonesty and Fraud (FRAUD)

**Definition:** Deliberate dishonesty, fraud, or deception in the practitioner’s professional conduct, including Medicare fraud, falsification of records, and misrepresentation of qualifications.

| Code | Subcategory | Definition |
|---|---|---|
| FRAUD_BILLING | Billing or Medicare fraud | Fraudulent billing, including billing for services not provided, upcoding, or Medicare fraud |
| FRAUD_RECORDS | Falsification of records | Deliberate alteration, fabrication, or destruction of clinical or professional records |
| FRAUD_QUALS | Misrepresentation of qualifications | Misrepresenting qualifications, registration status, or professional credentials |
| FRAUD_RESEARCH | Research misconduct | Fabrication, falsification, or plagiarism in research activities |
| FRAUD_OTHER | Other dishonesty | Other forms of professional dishonesty not captured above |

**Inclusion:** All forms of professional dishonesty, fraud, deception, or misrepresentation.

**Exclusion:** Inadequate record-keeping due to poor practice rather than deliberate falsification (CLIN_RECORD). Financial boundary violations with patients (BOUND_FINANCIAL).

#### A.6 Communication Failures (COMM)

**Definition:** Failures in professional communication with patients, families, colleagues, or regulatory bodies, including failures to obtain informed consent, inadequate disclosure, and failure to respond to complaints.

| Code | Subcategory | Definition |
|---|---|---|
| COMM_CONSENT | Informed consent failure | Failure to obtain adequate informed consent before treatment or procedures |
| COMM_DISCLOSURE | Inadequate disclosure | Failure to provide patients with adequate information about their condition, treatment, or outcomes |
| COMM_NOTIFY | Failure to report or notify | Failure to make mandatory notifications or reports to regulatory bodies, employers, or relevant authorities |
| COMM_CONDUCT | Unprofessional communication | Rude, aggressive, intimidating, or otherwise unprofessional communication with patients, families, or colleagues |

**Inclusion:** All matters where communication failures are the primary conduct concern.

**Exclusion:** Record-keeping failures (CLIN_RECORD). Dishonest communication (FRAUD). Boundary-related communication (BOUND).

#### A.7 Professional Conduct (PROF)

**Definition:** Breaches of general professional obligations and standards of conduct not captured by other categories, including failure to comply with conditions, advertising breaches, and practice management failures.

| Code | Subcategory | Definition |
|---|---|---|
| PROF_CONDITIONS | Breach of conditions | Failure to comply with conditions imposed on the practitioner’s registration |
| PROF_ADVERTISING | Advertising breach | Misleading, deceptive, or non-compliant advertising of health services |
| PROF_INSURANCE | Insurance or registration failure | Failure to maintain professional indemnity insurance or registration requirements |
| PROF_SUPERVISION | Supervision failure | Inadequate supervision of students, junior practitioners, or support staff |
| PROF_OTHER | Other professional conduct | Professional conduct issues not captured by other subcategories |

**Inclusion:** General professional conduct matters, breach of registration conditions, advertising, practice management, and matters not fitting other categories.

**Exclusion:** Clinical competence issues (CLIN). Communication failures (COMM). Boundary violations (BOUND).

---

### Appendix B: Keyword Filtering System

Decisions from tribunals with mixed jurisdiction (e.g., NCAT, VCAT, QCAT) were filtered using a two-tier keyword matching system to exclude non-health practitioner decisions. The system used 60 health-related terms organised into two tiers:

**Strong keywords** (one match sufficient): AHPRA, HCCC, Health Care Complaints Commission, Medical Board, Nursing and Midwifery Board, Pharmacy Board, Psychology Board, Dental Board, Chiropractic Board, Physiotherapy Board, Optometry Board, Osteopathy Board, Podiatry Board, Medical Radiation Practice Board, Occupational Therapy Board, Paramedicine Board, Chinese Medicine Board, Aboriginal and Torres Strait Islander Health Practice Board, Health Professional, Health Practitioner Regulation National Law, health practitioner tribunal, medical tribunal, nursing tribunal.

**General keywords** (two or more matches required): patient, clinical, practitioner, registration, medical, nursing, pharmacy, dental, psychology, chiropractic, physiotherapy, optometry, osteopathy, podiatry, midwifery, paramedic, health, doctor, nurse, pharmacist, dentist, psychologist, chiropractor, physiotherapist, surgeon, specialist, GP, general practitioner, hospital, clinic, treatment, diagnosis, prescription, medication, therapeutic, impairment, misconduct, unprofessional conduct, professional misconduct.

Keyword matching was case-insensitive and applied to the full text of each decision. This approach was designed to be inclusive (high sensitivity) at the cost of some false positives, which were manually reviewed during corpus assembly. The full keyword list and matching logic are available in the code repository.

---

### Appendix C: Supplementary Tables

The following tables were generated during the statistical analysis but are not included in the main paper. Tables S1 to S24 are based on the classifier's automated labels for the whole corpus; they are exploratory and should be read with the classifier's performance on the coded sample (main text Table 3; Table S30).

**Table S1. Decisions by Year (1999–2026; 2026 includes decisions to 9 February only)**

| Year | N | Year | N | Year | N |
|---|---|---|---|---|---|
| 1999 | 1 | 2009 | 8 | 2018 | 129 |
| 2001 | 2 | 2010 | 35 | 2019 | 178 |
| 2002 | 5 | 2011 | 65 | 2020 | 213 |
| 2003 | 2 | 2012 | 82 | 2021 | 255 |
| 2004 | 11 | 2013 | 67 | 2022 | 213 |
| 2005 | 11 | 2014 | 92 | 2023 | 229 |
| 2006 | 10 | 2015 | 114 | 2024 | 257 |
| 2007 | 14 | 2016 | 114 | 2025 | 234 |
| 2008 | 5 | 2017 | 100 | 2026 | 20 |

**Table S2. Misconduct Type by Profession**

| Code | Misconduct type | Overall N | Overall % | Medical practitioner | Nurse | Unknown | Pharmacist | Psychologist | Dentist | Chiropractor | Physiotherapist |
|---|---|---|---|---|---|---|---|---|---|---|---|
| CLIN | Clinical competence | 911 | 36.8 | 580 | 109 | 41 | 28 | 64 | 42 | 11 | 9 |
| PRESC | Prescribing | 724 | 29.2 | 418 | 84 | 97 | 104 | 3 | 11 | 2 | 1 |
| BOUND | Boundary violations | 719 | 29.0 | 362 | 118 | 86 | 3 | 93 | 5 | 16 | 12 |
| PROF | Professional conduct | 1293 | 52.2 | 480 | 420 | 130 | 93 | 48 | 53 | 22 | 8 |
| FRAUD | Dishonesty/ fraud | 559 | 22.6 | 177 | 162 | 102 | 32 | 37 | 15 | 14 | 3 |
| IMPR | Impairment | 292 | 11.8 | 135 | 84 | 42 | 10 | 6 | 7 | 0 | 1 |
| COMM | Communication | 339 | 13.7 | 219 | 33 | 13 | 4 | 22 | 15 | 11 | 8 |

*Note: Table shows the eight most represented professions (including unknown). Excluded due to small sample sizes: Chinese medicine (n = 16), Osteopath (n = 14), Paramedic (n = 14), Podiatrist (n = 9), Occupational therapist (n = 5), Midwife (n = 5), Optometrist (n = 4).*

**Table S3. Misconduct Co-occurrence Matrix**

| Misconduct type | Clinical competence | Prescribing | Boundary violations | Professional conduct | Dishonesty/fraud | Impairment | Communication |
|---|---|---|---|---|---|---|---|
| Clinical competence | 911 | 371 | 309 | 398 | 98 | 48 | 293 |
| Prescribing | 371 | 724 | 82 | 390 | 192 | 166 | 73 |
| Boundary violations | 309 | 82 | 719 | 192 | 81 | 34 | 192 |
| Professional conduct | 398 | 390 | 192 | 1293 | 437 | 178 | 154 |
| Dishonesty/fraud | 98 | 192 | 81 | 437 | 559 | 119 | 37 |
| Impairment | 48 | 166 | 34 | 178 | 119 | 292 | 8 |
| Communication | 293 | 73 | 192 | 154 | 37 | 8 | 339 |

**Table S4. Profession by Finding**

| Profession | N | Professional misconduct | Professional misconduct % | Unsatisfactory professional conduct | Unsatisfactory professional conduct % | Unprofessional conduct | Unprofessional conduct % | No finding | No finding % |
|---|---|---|---|---|---|---|---|---|---|
| Medical practitioner | 1135 | 944 | 83.2 | 56 | 4.9 | 37 | 3.3 | 98 | 8.6 |
| Nurse | 584 | 482 | 82.5 | 51 | 8.7 | 19 | 3.3 | 32 | 5.5 |
| Unknown | 313 | 277 | 88.5 | 5 | 1.6 | 10 | 3.2 | 21 | 6.7 |
| Pharmacist | 133 | 96 | 72.2 | 21 | 15.8 | 10 | 7.5 | 6 | 4.5 |
| Psychologist | 125 | 111 | 88.8 | 4 | 3.2 | 5 | 4.0 | 5 | 4.0 |
| Dentist | 65 | 56 | 86.2 | 5 | 7.7 | 3 | 4.6 | 1 | 1.5 |
| Chiropractor | 38 | 33 | 86.8 | 4 | 10.5 | 1 | 2.6 | 0 | 0.0 |
| Physiotherapist | 17 | 13 | 76.5 | 0 | 0.0 | 1 | 5.9 | 3 | 17.6 |
| Chinese medicine | 16 | 13 | 81.2 | 1 | 6.2 | 1 | 6.2 | 1 | 6.2 |
| Osteopath | 14 | 13 | 92.9 | 0 | 0.0 | 0 | 0.0 | 1 | 7.1 |

**Table S5. Opioid vs Non-Opioid Outcome Comparison**

| Group | N | Cancellation % | Suspension % | Conditions % | Reprimand % | Fine % | Caution % | Undertaking % | No adverse finding % |
|---|---|---|---|---|---|---|---|---|---|
| Opioid | 464 | 42.0 | 21.6 | 30.2 | 51.5 | 5.8 | 1.5 | 1.7 | 0.6 |

| Group | N | Cancellation % | Suspension % | Conditions % | Reprimand % | Fine % | Caution % | Undertaking % | No adverse finding % |
|---|---|---|---|---|---|---|---|---|---|
| Non-opioid | 260 | 32.3 | 23.5 | 31.9 | 53.8 | 6.2 | 2.3 | 0.8 | 0.4 |

**Table S6. Boundary Violation Subcategories**

| Subcategory | N | % of BOUND decisions |
|---|---|---|
| Sexual boundary violation | 657 | 91.4 |
| Emotional boundary violation | 575 | 80.0 |
| Financial boundary violation | 410 | 57.0 |
| Other/unclassified | 15 | 2.1 |

*Note: Keyword-based and exploratory (Methods D). Against the coded sample, the financial keywords had precision 0.21 (Results E); the coded sample's weighted rates among boundary-violation decisions are in Table S27.*

**Table S7. Boundary Violations vs Other Misconduct: Outcome Comparison**

| Group | N | Cancellation % | Suspension % | Conditions % | Reprimand % | Fine % | Caution % |
|---|---|---|---|---|---|---|---|
| Boundary violations | 719 | 42.0 | 23.6 | 19.9 | 48.4 | 4.5 | 2.5 |
| Other misconduct | 1758 | 38.7 | 20.9 | 27.8 | 53.6 | 6.3 | 2.8 |

**Table S8. Misconduct Type by Year**

| Year | Total | Clinical competence | Prescribing | Boundary violations | Professional conduct | Dishonesty/fraud | Impairment | Communication |
|---|---|---|---|---|---|---|---|---|
| 1999 | 1 | 0 | 0 | 0 | 1 | 1 | 0 | 0 |
| 2000 | 0 | 0 | 0 | 0 | 0 | 0 | 0 | 0 |
| 2001 | 2 | 1 | 0 | 0 | 1 | 0 | 1 | 0 |
| 2002 | 5 | 2 | 1 | 1 | 2 | 1 | 2 | 0 |
| 2003 | 2 | 0 | 0 | 1 | 1 | 0 | 0 | 0 |
| 2004 | 11 | 4 | 6 | 2 | 5 | 2 | 2 | 1 |
| 2005 | 11 | 4 | 3 | 4 | 3 | 0 | 3 | 0 |
| 2006 | 10 | 5 | 4 | 2 | 6 | 0 | 2 | 0 |
| 2007 | 14 | 9 | 8 | 3 | 8 | 1 | 3 | 1 |
| 2008 | 5 | 3 | 1 | 2 | 1 | 1 | 0 | 0 |
| 2009 | 8 | 4 | 4 | 2 | 4 | 1 | 2 | 0 |
| 2010 | 35 | 14 | 9 | 13 | 16 | 7 | 1 | 2 |
| 2011 | 65 | 25 | 16 | 20 | 23 | 4 | 2 | 5 |

| Year | Total | Clinical competence | Prescribing | Boundary violations | Professional conduct | Dishonesty/fraud | Impairment | Communication |
|---|---|---|---|---|---|---|---|---|
| 2012 | 82 | 42 | 25 | 18 | 29 | 13 | 11 | 10 |
| 2013 | 67 | 28 | 18 | 18 | 25 | 8 | 3 | 3 |
| 2014 | 92 | 43 | 23 | 19 | 34 | 15 | 10 | 5 |
| 2015 | 114 | 56 | 31 | 34 | 42 | 15 | 15 | 17 |
| 2016 | 114 | 47 | 34 | 33 | 40 | 21 | 18 | 7 |
| 2017 | 100 | 49 | 33 | 30 | 31 | 17 | 15 | 13 |
| 2018 | 129 | 50 | 43 | 36 | 68 | 25 | 18 | 17 |
| 2019 | 178 | 61 | 58 | 56 | 88 | 54 | 22 | 26 |
| 2020 | 213 | 48 | 71 | 55 | 127 | 67 | 30 | 14 |
| 2021 | 255 | 101 | 89 | 69 | 160 | 65 | 26 | 53 |
| 2022 | 213 | 89 | 73 | 72 | 126 | 56 | 31 | 41 |
| 2023 | 229 | 78 | 56 | 65 | 140 | 57 | 30 | 40 |
| 2024 | 257 | 79 | 65 | 83 | 160 | 59 | 26 | 58 |
| 2025 | 234 | 57 | 44 | 77 | 136 | 61 | 16 | 24 |
| 2026 | 20 | 6 | 8 | 3 | 16 | 8 | 3 | 2 |

**Table S9. Jurisdiction by Outcome**

| Jurisdiction | N | Cancellation % | Suspension % | Conditions % | Reprimand % | Fine % | Caution % | Undertaking % | No adverse finding % |
|---|---|---|---|---|---|---|---|---|---|
| NSW | 931 | 49.5 | 6.1 | 17.7 | 21.1 | 1.4 | 1.3 | 0.0 | 1.6 |
| VIC | 646 | 54.8 | 43.8 | 42.4 | 78.5 | 11.1 | 6.8 | 1.9 | 2.5 |
| QLD | 646 | 15.5 | 23.8 | 22.4 | 68.9 | 6.5 | 0.9 | 0.6 | 0.3 |
| SA | 123 | 30.9 | 16.3 | 12.2 | 65.9 | 3.3 | 0.8 | 0.0 | 0.8 |
| WA | 57 | 21.1 | 8.8 | 12.3 | 33.3 | 10.5 | 1.8 | 0.0 | 5.3 |
| ACT | 37 | 21.6 | 27.0 | 40.5 | 64.9 | 8.1 | 10.8 | 2.7 | 0.0 |
| NT | 27 | 22.2 | 18.5 | 29.6 | 48.1 | 7.4 | 0.0 | 0.0 | 0.0 |
| TAS | 10 | 40.0 | 40.0 | 30.0 | 60.0 | 0.0 | 0.0 | 0.0 | 0.0 |

**Table S10. Jurisdiction by Misconduct Type**

| Jurisdiction | N | Clinical competence % | Prescribing % | Boundary violations % | Professional conduct % | Dishonesty/fraud % | Impairment % | Communication % |
|---|---|---|---|---|---|---|---|---|
| NSW | 931 | 51.7 | 35.8 | 29.3 | 57.6 | 12.1 | 15.6 | 21.7 |
| VIC | 646 | 38.5 | 22.6 | 31.7 | 57.1 | 31.1 | 8.0 | 13.2 |
| QLD | 646 | 16.3 | 28.2 | 26.2 | 39.0 | 26.0 | 10.4 | 2.3 |
| SA | 123 | 22.0 | 28.5 | 27.6 | 77.2 | 41.5 | 14.6 | 14.6 |
| WA | 57 | 49.1 | 24.6 | 28.1 | 33.3 | 21.1 | 3.5 | 19.3 |
| ACT | 37 | 37.8 | 18.9 | 21.6 | 18.9 | 5.4 | 10.8 | 8.1 |
| NT | 27 | 14.8 | 22.2 | 33.3 | 48.1 | 44.4 | 14.8 | 11.1 |
| TAS | 10 | 30.0 | 10.0 | 50.0 | 20.0 | 0.0 | 0.0 | 20.0 |

**Table S11. COVID-19 Period Comparison**

| Period | N | CLIN % | PRESC % | BOUND % | PROF % | FRAUD % | IMPR % | COMM % | CANCEL % | REPRIMAND % |
|---|---|---|---|---|---|---|---|---|---|---|
| Pre-COVID (<2020) | 1045 | 42.8 | 30.3 | 28.1 | 41.0 | 17.8 | 12.4 | 10.2 | 35.6 | 41.8 |
| COVID (2020–2021) | 468 | 31.8 | 34.2 | 26.5 | 61.3 | 28.2 | 12.0 | 14.3 | 41.7 | 53.4 |
| Post-COVID (2022+) | 953 | 32.4 | 25.8 | 31.5 | 60.7 | 25.3 | 11.1 | 17.3 | 43.7 | 62.9 |

*Note: Total N = 2,466 (of 2,477 disciplinary decisions). 11 decisions were excluded due to missing date information.*

**Table S12. Error Analysis**

| Decision | Profession | Fields in error | Misconduct, coded | Misconduct, predicted | False positives | False negatives | Finding, coded | Finding, predicted | Outcome, coded | Outcome, predicted | Text length (characters) |
|---|---|---|---|---|---|---|---|---|---|---|---|
| qcat_2012_186 | pharmacist | misconduct, outcome | CLIN, PRESC | PRESC | | CLIN | | | CONDITIONS, SUSPEND | | 17466 |
| nswcatod_2018_103 | other | outcome | | | | | | | | REPRIMAND | 47239 |
| qcat_2023_432 | nurse | misconduct, finding, outcome | COMM, PROF | | | COMM, PROF | UPC_SAT | NONE | CAUTION, CONDITIONS | | 33073 |
| nswcatod_2015_139 | osteopath | misconduct, outcome | BOUND, CLIN, COMM | BOUND | | CLIN, COMM | | | CANCEL, CONDITIONS | CANCEL | 22563 |

| Decision | Profession | Fields in error | Misconduct, coded | Misconduct, predicted | False positives | False negatives | Finding, coded | Finding, predicted | Outcome, coded | Outcome, predicted | Text length (characters) |
|---|---|---|---|---|---|---|---|---|---|---|---|
| qcat_2021_51 | other | misconduct | CLIN, FRAUD, IMPR, PRESC | | | CLIN, FRAUD, IMPR, PRESC | | | | | 19011 |
| vcat_2021_1209 | medical_practitioner | misconduct, outcome | BOUND, PROF | BOUND, COMM, PROF | COMM | | | | CONDITIONS, REPRIMAND | CANCEL, CONDITIONS, REPRIMAND, SUSPEND | 53416 |
| nswmt_2012_13 | medical_practitioner | misconduct | BOUND, CLIN, PRESC | BOUND, CLIN, IMPR, PRESC | IMPR | | | | | | 43234 |
| vcat_2025_563 | pharmacist | outcome | | | | | | | CANCEL, REPRIMAND | CANCEL, CONDITIONS, REPRIMAND, SUSPEND | 99094 |
| nswcatod_2018_72 | pharmacist | misconduct, finding | FRAUD, PRESC, PROF | PRESC, PROF | | FRAUD | PM | UPC_SAT | | | 73824 |
| nswcatod_2021_26 | medical_practitioner | finding, outcome | | | | | UPC_SAT | PM | | CANCEL | 39571 |
| vcat_2024_358 | osteopath | outcome | | | | | | | REPRIMAND | CANCEL, CONDITIONS, REPRIMAND, SUSPEND | 87350 |
| nswcatod_2018_28 | medical_practitioner | misconduct, outcome | BOUND, CLIN, PRESC | CLIN, PRESC | | BOUND | | | | CANCEL | 147552 |
| nswcatod_2024_76 | medical_practitioner | misconduct, outcome | BOUND, CLIN | BOUND, CLIN, COMM | COMM | | | | | CANCEL | 35462 |
| nswcatod_2020_65 | chiropractor | misconduct, outcome | CLIN | CLIN, COMM | COMM | | | | CONDITIONS, REPRIMA | CANCEL, CONDITIONS, | 71042 |

| Decision | Profession | Fields in error | Misconduct, coded | Misconduct, predicted | False positives | False negatives | Finding, coded | Finding, predicted | Outcome, coded | Outcome, predicted | Text length (characters) |
|---|---|---|---|---|---|---|---|---|---|---|---|
| | | | | | | | | | ND | REPRIMAND | |
| nswcatod_2019_67 | nurse | misconduct | BOUND, PROF | BOUND | | PROF | | | | | 62692 |
| vcat_2021_68 | nurse | outcome | | | | | | | REPRIMAND | CONDITIONS, REPRIMAND | 54142 |
| tashpt_2014_4 | nurse | misconduct | FRAUD, PRESC, PROF | | | FRAUD, PRESC, PROF | | | | | 8117 |
| nswcatod_2022_169 | chiropractor | finding | | | | | PM | UPC_SAT | | | 84587 |
| vcat_2012_1615 | chinese_medicine_practitioner | misconduct, outcome | CLIN, PRESC | CLIN | | PRESC | | | CAUTION, CONDITIONS | CAUTION | 12463 |
| qcat_2024_465 | nurse | misconduct | FRAUD, IMPR, PROF | PROF | | FRAUD, IMPR | | | | | 26787 |
| nswcatod_2022_166 | pharmacist | misconduct | CLIN, IMPR, PRESC, PROF | CLIN, FRAUD, PRESC, PROF | FRAUD | IMPR | | | | | 73702 |
| vcat_2011_2002 | podiatrist | is_disciplinary, misconduct, finding, outcome | CLIN | N/A | N/A | CLIN | UPC_SAT | NONE | CONDITIONS | | 13618 |
| vcat_2018_1044 | nurse | outcome | | | | | | | | REPRIMAND | 35516 |
| vcat_2022_959 | psychologist | misconduct, outcome | BOUND, CLIN | BOUND, CLIN, COMM | COMM | | | | REPRIMAND | | 38537 |
| vcat_2024_232 | psychologist | misconduct, outcome | CLIN, COMM, FRAUD, PROF | CLIN, COMM, PROF | | FRAUD | | | CANCEL, REPRIMAND | CANCEL, CONDITIONS, REPRIMAND, SUSPEND, | 258014 |

| Decision | Profession | Fields in error | Misconduct, coded | Misconduct, predicted | False positives | False negatives | Finding, coded | Finding, predicted | Outcome, coded | Outcome, predicted | Text length (characters) |
|---|---|---|---|---|---|---|---|---|---|---|---|
| | | | | | | | | | | UNDERTAKING | |
| nswcatod_2023_31 | medical_practitioner | misconduct | BOUND, CLIN, COMM, FRAUD, PRESC, PROF | CLIN, PRESC, PROF | | BOUND, COMM, FRAUD | | | | | 49659 |
| qcat_2025_153 | medical_practitioner | misconduct, outcome | CLIN, COMM, PROF | PROF | | CLIN, COMM | | | REPRIMAND | CANCEL, REPRIMAND | 45991 |
| qcat_2012_613 | pharmacist | finding | | | | | NONE | UPC_GEN | | | 4374 |
| nswcatod_2020_141 | nurse | outcome | | | | | | | CANCEL | CANCEL, CONDITIONS | 112246 |
| qcat_2014_553 | psychologist | misconduct, outcome | PROF | | | PROF | | | CONDITIONS, REPRIMAND, SUSPEND | CONDITIONS | 14016 |
| vcat_2022_1042 | podiatrist | outcome | | | | | | | CONDITIONS, REPRIMAND, SUSPEND | CANCEL, CONDITIONS, REPRIMAND, SUSPEND | 65552 |
| qcat_2022_131 | medical_practitioner | misconduct | PROF | IMPR, PROF | IMPR | | | | | | 49588 |

**Table S13. Sensitivity: Excluding Appeals**

| Misconduct type | With appeals (%) | Without appeals (%) | Difference (pp) |
|---|---|---|---|
| Clinical competence | 36.8 | 36.3 | -0.5 |
| Prescribing | 29.2 | 29.1 | -0.1 |
| Boundary violations | 29.0 | 29.3 | 0.3 |
| Professional conduct | 52.2 | 52.4 | 0.2 |
| Dishonesty/fraud | 22.6 | 23.2 | 0.6 |

| Misconduct type | With appeals (%) | Without appeals (%) | Difference (pp) |
|---|---|---|---|
| Impairment | 11.8 | 11.8 | 0.0 |
| Communication | 13.7 | 13.4 | -0.3 |

**Table S14. Sensitivity: Pre-2015 vs Post-2015 Misconduct**

| Misconduct type | Pre-2015 N | Pre-2015 % | Post-2015 N | Post-2015 % | Chi2 | p-value |
|---|---|---|---|---|---|---|
| Clinical competence | 184 | 44.9 | 721 | 35.1 | 13.74 | 0.0002098 |
| Prescribing | 118 | 28.8 | 605 | 29.4 | 0.04 | 0.8393 |
| Boundary violations | 105 | 25.6 | 613 | 29.8 | 2.73 | 0.09854 |
| Professional conduct | 159 | 38.8 | 1134 | 55.2 | 36.1 | 1.877e-09 |
| Dishonesty/fraud | 54 | 13.2 | 505 | 24.6 | 24.66 | 6.842e-07 |
| Impairment | 42 | 10.2 | 250 | 12.2 | 1.03 | 0.3113 |
| Communication | 27 | 6.6 | 312 | 15.2 | 20.55 | 5.8e-06 |

**Table S15. Sensitivity: Pre-2015 vs Post-2015 Outcomes**

| Outcome | Pre-2015 % | Post-2015 % |
|---|---|---|
| CANCEL | 24.6 | 42.9 |
| SUSPEND | 17.8 | 22.6 |
| CONDITIONS | 26.8 | 25.4 |
| REPRIMAND | 40.2 | 54.5 |
| FINE | 5.9 | 5.7 |
| CAUTION | 7.1 | 1.9 |
| UNDERTAKING | 1.0 | 0.6 |
| NO_ADVERSE | 2.4 | 1.3 |

**Table S16. Sensitivity: Classification Thresholds**

| Threshold | Macro F1 | Micro F1 | Hamming Loss |
|---|---|---|---|
| 0.0 | 0.719 | 0.783 | 0.097 |
| 0.1 | 0.706 | 0.775 | 0.1 |
| 0.2 | 0.703 | 0.764 | 0.103 |
| 0.3 | 0.703 | 0.764 | 0.103 |
| 0.5 | 0.672 | 0.733 | 0.111 |
| 0.7 | 0.65 | 0.721 | 0.114 |
| 1.0 | 0.633 | 0.703 | 0.119 |

| Threshold | Macro F1 | Micro F1 | Hamming Loss |
|---|---|---|---|

**Table S17. Sensitivity: Per-Profession Classifier Performance**

| Profession | N (test) | Macro F1 | Micro F1 |
|---|---|---|---|
| medical_practitioner | 12 | 0.624 | 0.8 |
| nurse | 9 | 0.638 | 0.733 |
| pharmacist | 6 | 0.458 | 0.818 |
| physiotherapist | 3 | 0.25 | 1.0 |
| psychologist | 3 | 0.417 | 0.769 |

*Note: Per-profession test set sizes sum to 33, not the full test set of 45, because the remaining 12 test decisions involved professions not listed above (including unknown profession and professions with fewer than 3 test instances).*

**Table S18. Error-Adjusted Prevalence Estimates (Rogan-Gladen)**

Prevalence estimates adjusted for known classifier sensitivity and specificity using the Rogan-Gladen estimator. Sensitivity and specificity come from 5 to 22 positive test instances per category, so the adjusted figures are highly uncertain (for example, dishonesty/fraud rests on 10 test instances with sensitivity 0.40). These first-test-set estimates are superseded by the coded-sample estimates in Table S27; Table S28 re-estimates the adjustment using the coded sample.

| Code | Observed N | Observed % | Sensitivity | Specificity | Adjusted % | Direction of bias |
|---|---|---|---|---|---|---|
| CLIN | 911 | 36.8 | 0.706 | 1.0 | 52.1 | underestimate (low recall, no FP) |
| PRESC | 724 | 29.2 | 0.667 | 1.0 | 43.8 | underestimate (low recall, no FP) |
| BOUND | 719 | 29.0 | 0.75 | 1.0 | 38.7 | underestimate (low recall, no FP) |
| IMPR | 292 | 11.8 | 0.4 | 0.95 | 19.4 | uncertain (both FP and FN) |
| FRAUD | 559 | 22.6 | 0.4 | 0.971 | 53.1 | uncertain (both FP and FN) |
| COMM | 339 | 13.7 | 0.429 | 0.895 | 9.8 | uncertain (both FP and FN) |
| PROF | 1293 | 52.2 | 0.818 | 1.0 | 63.8 | underestimate (low recall, no FP) |

**Table S19. Bootstrap 95% Confidence Intervals for Per-Class F1**

| Code | F1 | 95% CI lower | 95% CI upper | Support (n) |
|---|---|---|---|---|
| CLIN | 0.8276 | 0.643 | 0.96 | 17 |

| Code | F1 | 95% CI lower | 95% CI upper | Support (n) |
|---|---|---|---|---|
| PRESC | 0.8 | 0.5 | 1.0 | 9 |
| BOUND | 0.8571 | 0.571 | 1.0 | 8 |
| IMPR | 0.4444 | 0.0 | 0.8 | 5 |
| FRAUD | 0.5333 | 0.167 | 0.8 | 10 |
| COMM | 0.4286 | 0.0 | 0.714 | 7 |
| PROF | 0.9 | 0.783 | 0.979 | 22 |

Overall: macro-F1 = 0.719, micro-F1 = 0.783 (test set, n = 45, scored once).

**Table S20. FDR Correction Summary**

| Test | Source | p-value | Rank | BH threshold | Adj. p | Sig. | FDR sig. |
|---|---|---|---|---|---|---|---|
| Jur. vs outcome | chi square | 2.60e-81 | 1 | 0.00079 | 1.64e-79 | Yes | Yes |
| jurisdiction pw. NSW vs QLD | chi square | 2.81e-77 | 2 | 0.00159 | 8.85e-76 | Yes | Yes |
| Prof. vs mc CLIN | chi square | 1.04e-62 | 3 | 0.00238 | 2.18e-61 | Yes | Yes |
| Prof. vs mc PRESC | chi square | 1.58e-62 | 4 | 0.00317 | 2.49e-61 | Yes | Yes |
| jurisdiction pw. VIC vs QLD | chi square | 1.47e-49 | 5 | 0.00397 | 1.85e-48 | Yes | Yes |
| Prof. vs mc BOUND | chi square | 3.43e-46 | 6 | 0.00476 | 3.60e-45 | Yes | Yes |
| Jur. vs mc CLIN | chi square | 1.44e-45 | 7 | 0.00556 | 1.30e-44 | Yes | Yes |
| Prof. vs mc PROF | chi square | 1.67e-39 | 8 | 0.00635 | 1.31e-38 | Yes | Yes |
| Jur. vs mc FRAUD | chi square | 3.72e-25 | 9 | 0.00714 | 2.61e-24 | Yes | Yes |
| Prof. vs mc COMM | chi square | 4.73e-25 | 10 | 0.00794 | 2.98e-24 | Yes | Yes |
| Jur. vs mc COMM | chi square | 9.14e-24 | 11 | 0.00873 | 5.23e-23 | Yes | Yes |
| Prof. vs outcome | chi square | 2.00e-23 | 12 | 0.00952 | 1.05e-22 | Yes | Yes |
| Jur. vs mc PROF | chi square | 3.12e-23 | 13 | 0.01032 | 1.51e-22 | Yes | Yes |
| covid vs mc PROF | chi square | 1.56e-21 | 14 | 0.01111 | 7.02e-21 | Yes | Yes |
| Prof. vs CANCEL | chi square | 8.51e-19 | 15 | 0.01190 | 3.58e-18 | Yes | Yes |
| Prof. vs REPRIMAND | chi square | 4.46e-15 | 16 | 0.01270 | 1.75e-14 | Yes | Yes |
| jurisdiction pw. | chi square | 1.12e-13 | 17 | 0.01349 | 4.15e-13 | Yes | Yes |

| Test | Source | p-value | Rank | BH threshold | Adj. p | Sig. | FDR sig. |
|---|---|---|---|---|---|---|---|
| NSW vs VIC | | | | | | | |
| Prof. vs mc FRAUD | chi square | 5.83e-12 | 18 | 0.01429 | 2.04e-11 | Yes | Yes |
| covid vs outcome | chi square | 3.49e-11 | 19 | 0.01508 | 1.16e-10 | Yes | Yes |
| trend TOTAL | mann kendall | 1.48e-10 | 20 | 0.01587 | 4.68e-10 | Yes | Yes |
| jurisdiction pw. VIC vs WA | chi square | 1.72e-10 | 21 | 0.01667 | 5.17e-10 | Yes | Yes |
| jurisdiction pw. NSW vs SA | chi square | 1.17e-09 | 22 | 0.01746 | 3.34e-09 | Yes | Yes |
| Temp. Professional conduct | temporal split | 1.88e-09 | 23 | 0.01825 | 5.14e-09 | Yes | Yes |
| jurisdiction pw. QLD vs WA | chi square | 1.98e-08 | 24 | 0.01905 | 5.19e-08 | Yes | Yes |
| jurisdiction pw. NSW vs WA | chi square | 3.22e-08 | 25 | 0.01984 | 8.12e-08 | Yes | Yes |
| jurisdiction pw. VIC vs SA | chi square | 1.47e-07 | 26 | 0.02063 | 3.55e-07 | Yes | Yes |
| covid vs mc CLIN | chi square | 5.39e-07 | 27 | 0.02143 | 1.26e-06 | Yes | Yes |
| Temp. Dishonesty/fraud | temporal split | 6.84e-07 | 28 | 0.02222 | 1.54e-06 | Yes | Yes |
| Prof. vs finding | chi square | 2.19e-06 | 29 | 0.02302 | 4.63e-06 | Yes | Yes |
| covid vs mc FRAUD | chi square | 2.20e-06 | 30 | 0.02381 | 4.63e-06 | Yes | Yes |
| Jur. vs mc PRESC | chi square | 2.68e-06 | 31 | 0.02460 | 5.45e-06 | Yes | Yes |
| Temp. Communication | temporal split | 5.80e-06 | 32 | 0.02540 | 1.14e-05 | Yes | Yes |
| trend COMM | mann kendall | 1.32e-05 | 33 | 0.02619 | 2.52e-05 | Yes | Yes |
| covid vs mc COMM | chi square | 2.50e-05 | 34 | 0.02698 | 4.62e-05 | Yes | Yes |
| jurisdiction pw. QLD vs SA | chi square | 6.16e-05 | 35 | 0.02778 | 1.11e-04 | Yes | Yes |
| Jur. vs mc IMPR | chi square | 1.39e-04 | 36 | 0.02857 | 2.44e-04 | Yes | Yes |
| Temp. Clinical competence | temporal split | 2.10e-04 | 37 | 0.02937 | 3.57e-04 | Yes | Yes |

| Test | Source | p-value | Rank | BH threshold | Adj. p | Sig. | FDR sig. |
|---|---|---|---|---|---|---|---|
| Prof. vs CAUTION | chi square | 5.58e-04 | 38 | 0.03016 | 9.25e-04 | Yes | Yes |
| trend FRAUD | mann kendall | 7.64e-04 | 39 | 0.03095 | 1.23e-03 | Yes | Yes |
| covid vs mc PRESC | chi square | 3.14e-03 | 40 | 0.03175 | 4.95e-03 | Yes | Yes |
| Prof. vs SUSPEND | chi square | 4.24e-03 | 41 | 0.03254 | 6.51e-03 | Yes | Yes |
| Prof. vs mc IMPR | chi square | 1.01e-02 | 42 | 0.03333 | 1.51e-02 | Yes | Yes |
| opioid vs CANCEL | chi square | 1.25e-02 | 43 | 0.03413 | 1.83e-02 | Yes | Yes |
| Prof. vs CONDITIONS | chi square | 4.69e-02 | 44 | 0.03492 | 6.71e-02 | Yes | No |
| jurisdiction pw. SA vs WA | chi square | 5.87e-02 | 45 | 0.03571 | 8.22e-02 | No | No |
| trend CLIN | mann kendall | 8.69e-02 | 46 | 0.03651 | 1.19e-01 | No | No |
| trend BOUND | mann kendall | 9.52e-02 | 47 | 0.03730 | 1.27e-01 | No | No |
| Temp. Boundary violations | temporal split | 9.85e-02 | 48 | 0.03810 | 1.27e-01 | No | No |
| covid vs mc BOUND | chi square | 9.90e-02 | 49 | 0.03889 | 1.27e-01 | No | No |
| Prof. vs FINE | chi square | 1.10e-01 | 50 | 0.03968 | 1.38e-01 | No | No |
| trend PRESC | mann kendall | 1.22e-01 | 51 | 0.04048 | 1.51e-01 | No | No |
| fisher boundary vs CANCEL | chi square | 1.36e-01 | 52 | 0.04127 | 1.64e-01 | No | No |
| boundary vs CANCEL | chi square | 1.44e-01 | 53 | 0.04206 | 1.71e-01 | No | No |
| trend PROF | mann kendall | 1.50e-01 | 54 | 0.04286 | 1.74e-01 | No | No |
| boundary vs SUSPEND | chi square | 1.52e-01 | 55 | 0.04365 | 1.74e-01 | No | No |
| Prof. vs NO ADVERSE | chi square | 1.60e-01 | 56 | 0.04444 | 1.81e-01 | No | No |
| Jur. vs mc BOUND | chi square | 2.97e-01 | 57 | 0.04524 | 3.29e-01 | No | No |
| Temp. Impairment | temporal split | 3.11e-01 | 58 | 0.04603 | 3.38e-01 | No | No |
| trend IMPR | mann kendall | 3.81e-01 | 59 | 0.04683 | 4.07e-01 | No | No |

| Test | Source | p-value | Rank | BH threshold | Adj. p | Sig. | FDR sig. |
|---|---|---|---|---|---|---|---|
| opioid vs SUSPEND | chi square | 6.17e-01 | 60 | 0.04762 | 6.48e-01 | No | No |
| covid vs mc IMPR | chi square | 6.58e-01 | 61 | 0.04841 | 6.77e-01 | No | No |
| Prof. vs UNDERTAKING | chi square | 6.66e-01 | 62 | 0.04921 | 6.77e-01 | No | No |
| Temp. Prescribing | temporal split | 8.39e-01 | 63 | 0.05000 | 8.39e-01 | No | No |

Benjamini-Hochberg FDR correction was applied to all 63 statistical tests. Of the 44 tests significant before correction, 43 remained significant after correction (lost: profession_vs_CONDITIONS). Results reported as significant in the manuscript are significant after FDR correction; the Mann-Kendall trends for BOUND and PROF and the boundary violation–cancellation association were not significant before or after correction.

#### Table S21. Phi Coefficient Matrix for Misconduct Type Co-occurrence

Phi coefficients (equivalent to Pearson correlation for binary variables) between the seven misconduct types. Positive values indicate co-occurrence; negative values indicate inverse association.

| | CLIN | PRESC | BOUND | PROF | FRAUD | IMPR | COMM |
|---|---|---|---|---|---|---|---|
| CLIN | 1.0 | 0.19 | 0.08 | -0.13 | -0.22 | -0.15 | 0.41 |
| PRESC | 0.19 | 1.0 | -0.25 | 0.02 | 0.06 | 0.22 | -0.07 |
| BOUND | 0.08 | -0.25 | 1.0 | -0.33 | -0.17 | -0.14 | 0.24 |
| PROF | -0.13 | 0.02 | -0.33 | 1.0 | 0.28 | 0.06 | -0.05 |
| FRAUD | -0.22 | 0.06 | -0.17 | 0.28 | 1.0 | 0.16 | -0.11 |
| IMPR | -0.15 | 0.22 | -0.14 | 0.06 | 0.16 | 1.0 | -0.12 |
| COMM | 0.41 | -0.07 | 0.24 | -0.05 | -0.11 | -0.12 | 1.0 |

#### Table S22. Misconduct Multi-Label Combinations (n >= 10)

| combination | n_types | n | pct |
|---|---|---|---|
| PROF | 1 | 317 | 12.8 |
| BOUND | 1 | 244 | 9.9 |
| PROF + FRAUD | 2 | 209 | 8.4 |
| CLIN + PRESC + PROF | 3 | 112 | 4.5 |
| CLIN + PRESC | 2 | 111 | 4.5 |
| CLIN | 1 | 109 | 4.4 |
| CLIN + BOUND + COMM | 3 | 98 | 4.0 |
| PRESC | 1 | 80 | 3.2 |

| combination | n_types | n | pct |
|---|---|---|---|
| CLIN + BOUND | 2 | 71 | 2.9 |
| CLIN + PROF | 2 | 64 | 2.6 |
| BOUND + PROF | 2 | 61 | 2.5 |
| PRESC + PROF + FRAUD | 3 | 55 | 2.2 |
| PRESC + PROF + FRAUD + IMPR | 4 | 54 | 2.2 |
| PRESC + PROF | 2 | 52 | 2.1 |
| PROF + IMPR | 2 | 45 | 1.8 |
| CLIN + PROF + COMM | 3 | 40 | 1.6 |
| CLIN + COMM | 2 | 35 | 1.4 |
| IMPR | 1 | 31 | 1.3 |
| PRESC + FRAUD + IMPR | 3 | 29 | 1.2 |
| CLIN + BOUND + PROF + COMM | 4 | 29 | 1.2 |
| CLIN + PRESC + PROF + COMM | 4 | 29 | 1.2 |
| PRESC + PROF + IMPR | 3 | 26 | 1.0 |
| BOUND + PROF + FRAUD | 3 | 23 | 0.9 |
| CLIN + PROF + FRAUD | 3 | 22 | 0.9 |
| PRESC + IMPR | 2 | 22 | 0.9 |
| PROF + FRAUD + IMPR | 3 | 21 | 0.8 |
| FRAUD | 1 | 20 | 0.8 |
| CLIN + PRESC + BOUND | 3 | 19 | 0.8 |
| BOUND + FRAUD | 2 | 19 | 0.8 |
| CLIN + BOUND + PROF | 3 | 18 | 0.7 |
| BOUND + PROF + | 3 | 16 | 0.6 |

| combination | n_types | n | pct |
|---|---|---|---|
| COMM | | | |
| PRESC + FRAUD | 2 | 15 | 0.6 |
| PRESC + BOUND | 2 | 14 | 0.6 |
| CLIN + PRESC + PROF + FRAUD | 4 | 13 | 0.5 |
| CLIN + PRESC + COMM | 3 | 12 | 0.5 |
| CLIN + BOUND + FRAUD | 3 | 10 | 0.4 |
| BOUND + COMM | 2 | 10 | 0.4 |
| CLIN + PRESC + BOUND + PROF | 4 | 10 | 0.4 |
| CLIN + PRESC + PROF + IMPR | 4 | 10 | 0.4 |
| CLIN + PRESC + BOUND + COMM | 4 | 10 | 0.4 |

**Table S23. Misconduct Profile Clusters (Ward's Hierarchical Clustering)**

| cluster | n | pct_of_disciplinary | dominant_misconduct | top_professions | cancellation_rate_pct |
|---|---|---|---|---|---|
| 1 | 222 | 9.0 | PRESC (91.0%), PROF (48.2%), FRAUD (40.5%) | Medical Practitioner (72), Pharmacist (69), Unknown (46) | 32.4 |
| 2 | 277 | 11.2 | IMPR (100.0%), PROF (59.2%), PRESC (55.2%), FRAUD (40.1%), CLIN (13.0%), BOUND (8.7%) | Medical Practitioner (125), Nurse (84), Unknown (41) | 50.5 |
| 3 | 317 | 12.8 | PROF (100.0%) | Nurse (143), Medical Practitioner (90), Unknown (41) | 43.8 |
| 4 | 278 | 11.2 | FRAUD (100.0%), PROF (95.0%), CLIN (24.8%), PRESC (11.9%), COMM (6.1%) | Nurse (88), Medical Practitioner (84), Unknown (42) | 47.8 |
| 5 | 519 | 21.0 | BOUND (100.0%), CLIN (28.5%), PROF (25.4%), FRAUD (14.3%), PRESC (10.2%) | Medical Practitioner (225), Nurse (102), Psychologist (77) | 41.4 |

| cluster | n | pct_of_disciplinary | dominant_misconduct | top_professions | cancellation_rate_pct |
|---|---|---|---|---|---|
| 6 | 302 | 12.2 | COMM (100.0%), CLIN (86.8%), BOUND (57.0%), PROF (44.0%), PRESC (19.9%) | Medical Practitioner (201), Nurse (31), Psychologist (16) | 44.0 |
| 7 | 173 | 7.0 | CLIN (100.0%), PROF (37.0%) | Medical Practitioner (83), Nurse (53), Dentist (14) | 27.7 |
| 8 | 223 | 9.0 | CLIN (100.0%), PRESC (100.0%), PROF (50.2%) | Medical Practitioner (176), Pharmacist (19), Nurse (12) | 34.1 |

Hierarchical clustering (Ward's method) on the binary misconduct type matrix identified 8 clusters (silhouette score = 0.4). Each cluster is characterised by its dominant misconduct types, top professions, and cancellation rate.

#### Table S24. Mann-Kendall Sensitivity: Excluding Pre-2005 Years

| misconduct_type | range | n_years | year_start | year_end | tau | p_value | slope | trend | significant |
|---|---|---|---|---|---|---|---|---|---|
| CLIN | full_range | 27 | 1999 | 2026 | -0.236 | 0.08691 | -0.524 | no trend | False |
| CLIN | 2005_onwards | 22 | 2005 | 2026 | -0.528 | 0.000642 | -1.2222 | decreasing | True |
| PRESC | full_range | 27 | 1999 | 2026 | 0.214 | 0.122368 | 0.5627 | no trend | False |
| PRESC | 2005_onwards | 22 | 2005 | 2026 | 0.022 | 0.910124 | 0.052 | no trend | False |
| BOUND | full_range | 27 | 1999 | 2026 | 0.231 | 0.095221 | 0.4712 | no trend | False |
| BOUND | 2005_onwards | 22 | 2005 | 2026 | 0.065 | 0.693012 | 0.1174 | no trend | False |
| PROF | full_range | 27 | 1999 | 2026 | 0.199 | 0.149985 | 0.6031 | no trend | False |
| PROF | 2005_onwards | 22 | 2005 | 2026 | 0.394 | 0.011155 | 1.5509 | increasing | True |
| FRAUD | full_range | 27 | 1999 | 2026 | 0.462 | 0.000764 | 1.0584 | increasing | True |
| FRAUD | 2005_onwards | 22 | 2005 | 2026 | 0.645 | 3e-05 | 1.2765 | increasing | True |
| IMPR | full_range | 27 | 1999 | 2026 | -0.123 | 0.38078 | -0.2748 | no trend | False |
| IMPR | 2005_onwards | 22 | 2005 | 2026 | -0.113 | 0.480668 | -0.25 | no trend | False |
| COMM | full_range | 27 | 1999 | 2026 | 0.59 | 1.3e-05 | 0.7054 | increasing | True |
| COMM | 2005_onwards | 22 | 2005 | 2026 | 0.567 | 0.000235 | 0.8643 | increasing | True |
| TOTAL | full_range | 27 | 1999 | 2025 | 0.877 | 0.0 | 10.5455 | increasing | True |
| TOTAL | 2005_onwards | 21 | 2005 | 2025 | 0.857 | 0.0 | 13.6125 | increasing | True |

Sensitivity analysis re-running Mann-Kendall trend tests on years 2005–2026 only (excluding the early period 1999–2004 when fewer than 10 disciplinary decisions were issued per year). The significant increasing trends for FRAUD, COMM, and total volume persisted. Two trends that were not significant over the full range became significant: PROF increased (tau = 0.394, $p$ = 0.011) and CLIN decreased (tau = -0.528, $p$ = 0.0006). BOUND showed no trend in either range (full range tau = 0.231, $p$ = 0.095; 2005 onwards tau = 0.065, $p$ = 0.693). $p$-values here are unadjusted.

#### Table S25. Inter-Rater Agreement: Author's Blind Codes vs Logged Model Pass (N = 300)

Cohen's kappa with 95% bootstrap confidence intervals (2,000 resamples), computed on the frozen blind codes before adjudication. Misconduct, boundary subtype, and outcome fields are binary (code present or absent).

| Field | Kappa | 95% CI | % agreement | Positives (author / model) |
|---|---|---|---|---|
| is_disciplinary | 0.968 | 0.931–0.992 | 98.7 | 213 / 213 |
| finding | 0.943 | 0.906–0.973 | 96.7 | |
| profession | 0.953 | 0.925–0.977 | 96.0 | |
| CLIN | 0.776 | 0.701–0.846 | 89.7 | 117 / 98 |
| PRESC | 0.897 | 0.833–0.949 | 96.3 | 73 / 66 |
| BOUND | 0.903 | 0.839–0.958 | 96.7 | 70 / 62 |
| IMPR | 0.869 | 0.748–0.959 | 98.0 | 25 / 25 |
| FRAUD | 0.871 | 0.804–0.931 | 95.3 | 71 / 71 |
| COMM | 0.597 | 0.446–0.728 | 90.7 | 43 / 37 |
| PROF | 0.747 | 0.67–0.818 | 87.3 | 150 / 120 |
| BOUND_SEXUAL | 0.94 | 0.88–0.987 | 98.3 | 52 / 49 |
| BOUND_EMOTIONAL | 0.684 | 0.545–0.793 | 93.0 | 48 / 27 |
| BOUND_FINANCIAL | 0.762 | 0.575–0.92 | 97.7 | 18 / 13 |
| CANCEL | 0.908 | 0.838–0.967 | 97.3 | 49 / 57 |
| SUSPEND | 1.0 | 1.0–1.0 | 100.0 | 35 / 35 |
| CONDITIONS | 0.956 | 0.914–0.991 | 98.3 | 75 / 78 |
| REPRIMAND | 1.0 | 1.0–1.0 | 100.0 | 126 / 126 |
| FINE | 1.0 | 1.0–1.0 | 100.0 | 11 / 11 |
| CAUTION | 0.921 | 0.665–1.0 | 99.7 | 7 / 6 |
| UNDERTAKING | 0.0 | 0.0–0.0 | 99.3 | 2 / 0 |
| NO_ADVERSE | 0.887 | 0.497–1.0 | 99.7 | 5 / 4 |
| OTHER | 0.635 | 0.551–0.719 | 82.0 | 144 / 100 |

### Table S26. Post-Hoc Sensitivity: Test Performance Against the Author's Blind Codes Alone (N = 45)

The classifiers chosen under the pre-specified protocol, scored on the same 45 test decisions against the gold standard and against the author's blind codes with no adjudication or model input. This analysis was added after the test set had been scored; nothing was re-selected on it. The author's codes differed from the gold standard in the misconduct labels of 9 decisions and the outcome labels of 7; disciplinary status and finding did not differ.

| Metric | Gold standard | Author's blind codes only |
|---|---|---|
| Is-disciplinary accuracy | 0.978 | 0.978 |
| Misconduct macro-F1 | 0.719 | 0.721 |
| Misconduct micro-F1 | 0.783 | 0.774 |
| Finding accuracy | 0.867 | 0.867 |
| Outcome regex micro-F1 | 0.694 | 0.694 |
| F1: CLIN | 0.828 | 0.828 |
| F1: PRESC | 0.800 | 0.800 |
| F1: BOUND | 0.857 | 0.750 |
| F1: IMPR | 0.444 | 0.444 |
| F1: FRAUD | 0.533 | 0.625 |
| F1: COMM | 0.429 | 0.500 |
| F1: PROF | 0.900 | 0.857 |

### Table S27. Coded Sample: Weighted Estimates (n = 160 decisions; 109 disciplinary)

Weighted ratio estimates (weight = pool size of the period / 40) with 95% CIs from 10,000 bootstrap resamples within periods. Percentages are of disciplinary decisions, except boundary subtypes (of the 39 coded boundary decisions) and the first row (of all 160).

| Group | Measure | Weighted % (95% CI) | n |
|---|---|---|---|
| All decisions | Disciplinary (share of all decisions) | 70.3 (61.6-78.6) | 109/160 |
| Misconduct type | Clinical competence | 38.6 (27.5-50.3) | 51/109 |
| Misconduct type | Prescribing/dispensing | 24.7 (14.9-35.3) | 31/109 |
| Misconduct type | Boundary violations | 36.3 (24.8-48.0) | 39/109 |
| Misconduct type | Impairment | 7.5 (2.3-14.2) | 13/109 |
| Misconduct type | Dishonesty/fraud | 41.6 (29.7-53.1) | 39/109 |
| Misconduct type | Communication | 5.5 (0.6-12.0) | 5/109 |
| Misconduct type | Professional conduct | 68.7 (58.9-77.5) | 61/109 |
| Boundary subtype | Sexual (among boundary) | 83.6 (70.0-94.5) | 31/39 |
| Boundary subtype | Emotional (among boundary) | 76.4 (58.4-92.4) | 30/39 |

| Group | Measure | Weighted % (95% CI) | n |
|---|---|---|---|
| Boundary subtype | Financial (among boundary) | 5.5 (1.4-9.6) | 4/39 |
| Finding | Professional misconduct | 83.0 (74.7-90.4) | 84/109 |
| Finding | Unsatisfactory professional conduct | 10.0 (4.2-17.2) | 16/109 |
| Finding | Unprofessional conduct | 3.0 (0.5-5.9) | 5/109 |
| Finding | No finding | 4.0 (0.1-9.1) | 4/109 |
| Outcome | Cancellation | 30.6 (19.8-42.4) | 38/109 |
| Outcome | Suspension | 20.8 (11.9-31.1) | 20/109 |
| Outcome | Conditions | 40.0 (28.7-51.6) | 48/109 |
| Outcome | Reprimand | 70.9 (61.0-79.9) | 59/109 |
| Outcome | Fine | 6.6 (1.4-13.1) | 7/109 |
| Outcome | Caution | 1.5 (0.0-3.5) | 3/109 |
| Outcome | Undertaking | 0.1 (0.0-0.3) | 1/109 |
| Outcome | No adverse finding | 2.0 (0.0-4.9) | 2/109 |
| Outcome | Other orders | 39.2 (27.5-51.1) | 41/109 |
| Profession | Medical practitioner | 41.7 (30.8-53.0) | 68/109 |
| Profession | Nurse or midwife | 29.7 (18.8-41.0) | 21/109 |
| Profession | Other profession | 28.7 (17.8-40.0) | 20/109 |

#### Table S28. Rogan-Gladen Adjusted Prevalence Using the Coded Sample (Sensitivity Analysis)

Sensitivity and specificity of the classifier on the coded sample (weighted). Adjusted values are unstable where sensitivity is low and are not primary estimates.

| Misconduct type | Classifier corpus % | Sensitivity | Specificity | Adjusted % |
|---|---|---|---|---|
| Clinical competence | 36.8 | 0.593 | 0.959 | 59.2 |
| Prescribing/dispensing | 29.2 | 0.896 | 0.947 | 28.4 |
| Boundary violations | 29.0 | 0.807 | 0.991 | 35.2 |
| Impairment | 11.8 | 0.5 | 0.981 | 20.6 |
| Dishonesty/fraud | 22.6 | 0.541 | 0.955 | 36.4 |
| Communication | 13.7 | 0.285 | 0.956 | 38.5 |
| Professional conduct | 52.2 | 0.662 | 0.921 | 76.0 |

#### Table S29. Classifier Error over Time and Trend Check on the Coded Sample

Odds ratios per decade from logistic regressions on decision year, for detection among coded positives and for false positives among coded negatives. Prevalence by period (1999-2009, 2010-2014, 2015-2019, 2020-2026) among decisions predicted, or coded, disciplinary. A trend is robust if predicted and coded prevalence move in the same direction and neither error rate shows clear dependence on time (95% CI includes 1).

| Misconduct type | Coded positives | Detection OR per decade | False-positive OR per decade | Predicted % by period | Coded % by period | Same direction | Robust |
|---|---|---|---|---|---|---|---|
| Clinical competence | 51 | 0.58 (0.22-1.51) | 0.63 (0.20-1.98) | 48, 45, 45, 16 | 57, 52, 50, 31 | yes | yes |
| Prescribing/ dispensing | 31 | 1.18 (0.16-8.56) | 0.83 (0.27-2.56) | 36, 23, 35, 26 | 36, 28, 30, 22 | yes | yes |
| Boundary violations | 39 | 1.17 (0.38-3.59) | 3.67 (0.08-170.92) | 23, 26, 30, 32 | 29, 48, 25, 38 | yes | yes |
| Impairment | 13 | 2.01 (0.29-14.18) | 0.84 (0.15-4.67) | 13, 6, 5, 6 | 21, 14, 5, 6 | yes | yes |
| Dishonesty/fraud | 39 | 2.83 (1.11-7.25) | 0.97 (0.19-5.01) | 13, 6, 20, 36 | 43, 14, 35, 50 | yes | no |
| Communication | 5 | descriptive only | 2.83 (0.57-14.09) | 6, 3, 10, 6 | 4, 3, 5, 6 | yes | not assessable (fewer than 10 coded cases) |
| Professional conduct | 61 | 1.41 (0.71-2.82) | 0.57 (0.17-1.85) | 48, 10, 45, 64 | 57, 28, 50, 84 | yes | yes |

#### Table S30. Classifier and Language-Model Coder on the Coded Sample, Overall and by Period

| Set | Disciplinary accuracy | Misconduct macro-F1 | Finding accuracy | Outcome micro-F1 | Dishonesty/fraud F1 |
|---|---|---|---|---|---|
| Weighted (n=160) | 0.977 (0.94-1.00) | 0.696 (0.61-0.77) | 0.886 (0.83-0.94) | 0.739 (0.68-0.80) | 0.656 (0.46-0.81) |
| Period 1999-2009 | 0.925 (0.82-1.00) | 0.717 (0.55-0.81) | 0.775 (0.65-0.90) | 0.559 (0.38-0.69) | 0.375 (0.00-0.67) |
| Period 2010-2014 | 0.950 (0.88-1.00) | 0.677 (0.42-0.79) | 0.725 (0.57-0.85) | 0.688 (0.53-0.80) | 0.333 (0.00-0.80) |
| Period 2015-2019 | 1.000 (1.00-1.00) | 0.808 (0.56-0.90) | 0.900 (0.80-0.97) | 0.623 (0.44-0.76) | 0.727 (0.29-1.00) |
| Period 2020-2026 | 0.975 (0.93-1.00) | 0.645 (0.52-0.75) | 0.925 (0.82-1.00) | 0.783 (0.71-0.85) | 0.667 (0.42-0.85) |
| Combined author reference (n=205) | 0.966 (0.94-0.99) | 0.716 (0.66-0.76) | 0.839 (0.79-0.89) | 0.686 (0.63-0.74) | 0.579 (0.43-0.70) |
| Coder, weighted (n=160) | 0.960 (0.92-0.99) | 0.765 (0.69-0.83) | 0.935 (0.88-0.97) | 0.942 (0.91-0.97) | 0.852 (0.73-0.94) |
| Coder, period 1999-2009 | 0.950 (0.88-1.00) | 0.793 (0.75-0.82) | 0.900 (0.80-0.97) | 0.960 (0.91-1.00) | 0.917 (0.77-1.00) |
| Coder, period 2010-2014 | 0.975 (0.93-1.00) | 0.751 (0.63-0.82) | 0.900 (0.80-0.97) | 0.963 (0.91-1.00) | 0.800 (0.33-1.00) |
| Coder, period 2015-2019 | 0.925 (0.85-1.00) | 0.858 (0.62-0.92) | 0.925 (0.82-1.00) | 0.903 (0.79-0.98) | 0.875 (0.67-1.00) |
| Coder, period 2020-2026 | 0.975 (0.93-1.00) | 0.736 (0.60-0.83) | 0.950 (0.88-1.00) | 0.946 (0.90-0.98) | 0.849 (0.69-0.97) |

**Table S31. Rates per 10,000 Registrants, Ten Professions Regulated Since 2010: Sensitivity Analyses**

| Analysis | Rate 2011-13 | Rate 2023-25 | Rate ratio per year (95% CI) |
|---|---|---|---|
| Primary (excluding pandemic sub-register) | 1.252 | 2.364 | 1.050 (1.029-1.071) |
| Including pandemic sub-register | 1.252 | 2.364 | 1.049 (1.029-1.069) |
| Unknown profession allocated | 1.282 | 2.795 | 1.066 (1.043-1.089) |

**Table S32. Checks on the Coded-Sample Estimates: Practitioner-Level Weighting and Pool Exclusion**

Practitioner-level: each coded disciplinary decision is re-weighted by the inverse of the number of corpus decisions about the same practitioner (identified approximately by the coder’s recorded surname in the citation, or in the opening party details for neutral citations); 24 of 109 coded disciplinary decisions had related decisions (maximum 10). Pool exclusion: classifier prevalence among decisions it labelled disciplinary, in the sampling pool (n = 2,053) and in the 600 excluded labelled decisions (n = 416). Cancellation and reprimand are blank in the classifier columns because the check compares misconduct types only.

| Measure | Coded sample, primary (weighted %) | Coded sample, practitioner-level (weighted %) | Classifier, sampling pool % | Classifier, excluded 600 % |
|---|---|---|---|---|
| Professional conduct | 68.7 | 69.8 | 52.2 | 53.1 |
| Dishonesty/fraud | 41.6 | 43.3 | 21.6 | 27.6 |
| Clinical competence | 38.6 | 36.8 | 36.1 | 39.7 |
| Boundary violations | 36.3 | 37.4 | 28.7 | 31.2 |
| Prescribing/dispensing | 24.7 | 22.5 | 29.0 | 30.8 |
| Impairment | 7.5 | 7.8 | 11.3 | 14.2 |
| Communication | 5.5 | 5.1 | 12.9 | 18.0 |
| Cancellation | 30.6 | 29.7 | | |
| Reprimand | 70.9 | 77.5 | | |

### Appendix D: Supplementary Figures

The following figures were generated during the analysis but are not included in the main paper.

**Figure S1. Decisions per Year by Jurisdiction (Stacked Bar)**

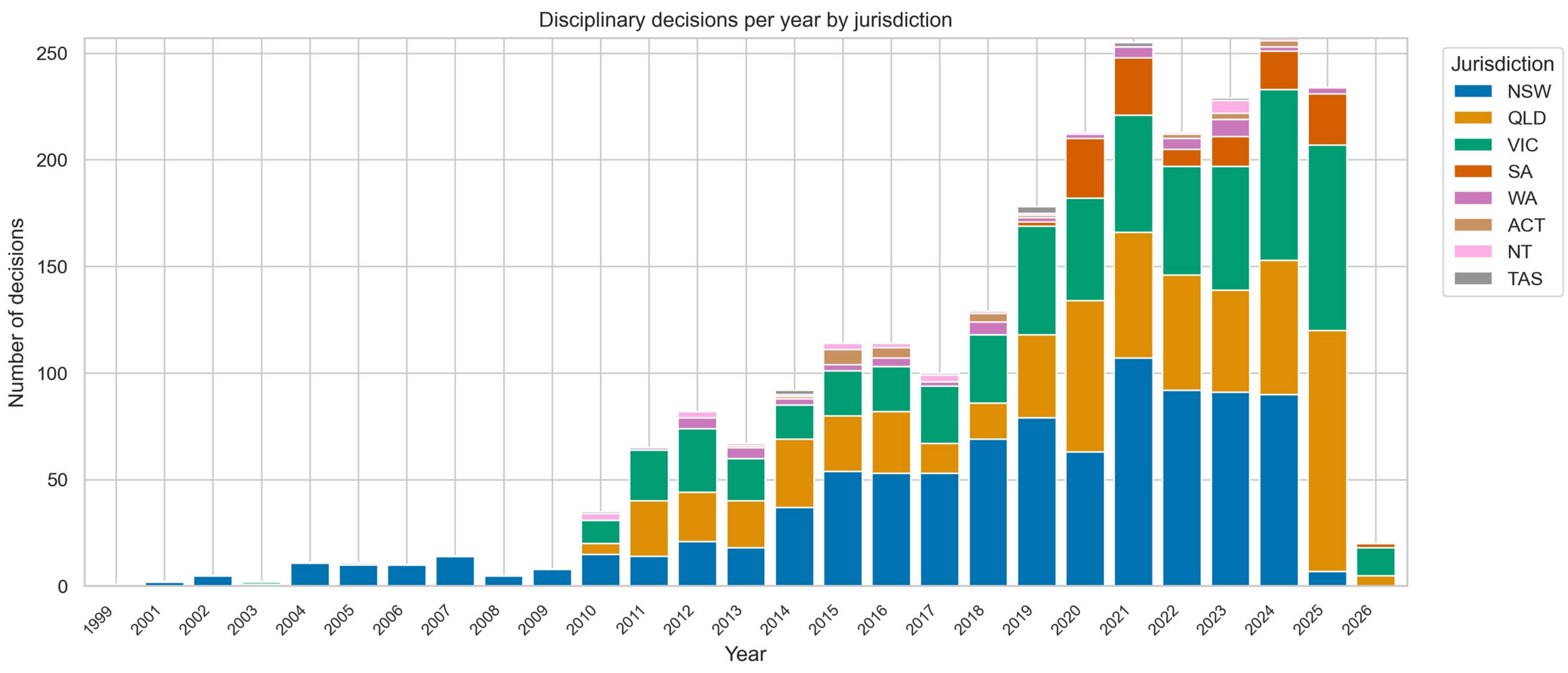

**Figure S2. Cancellation and Suspension Rates by Profession (Grouped Bar)**

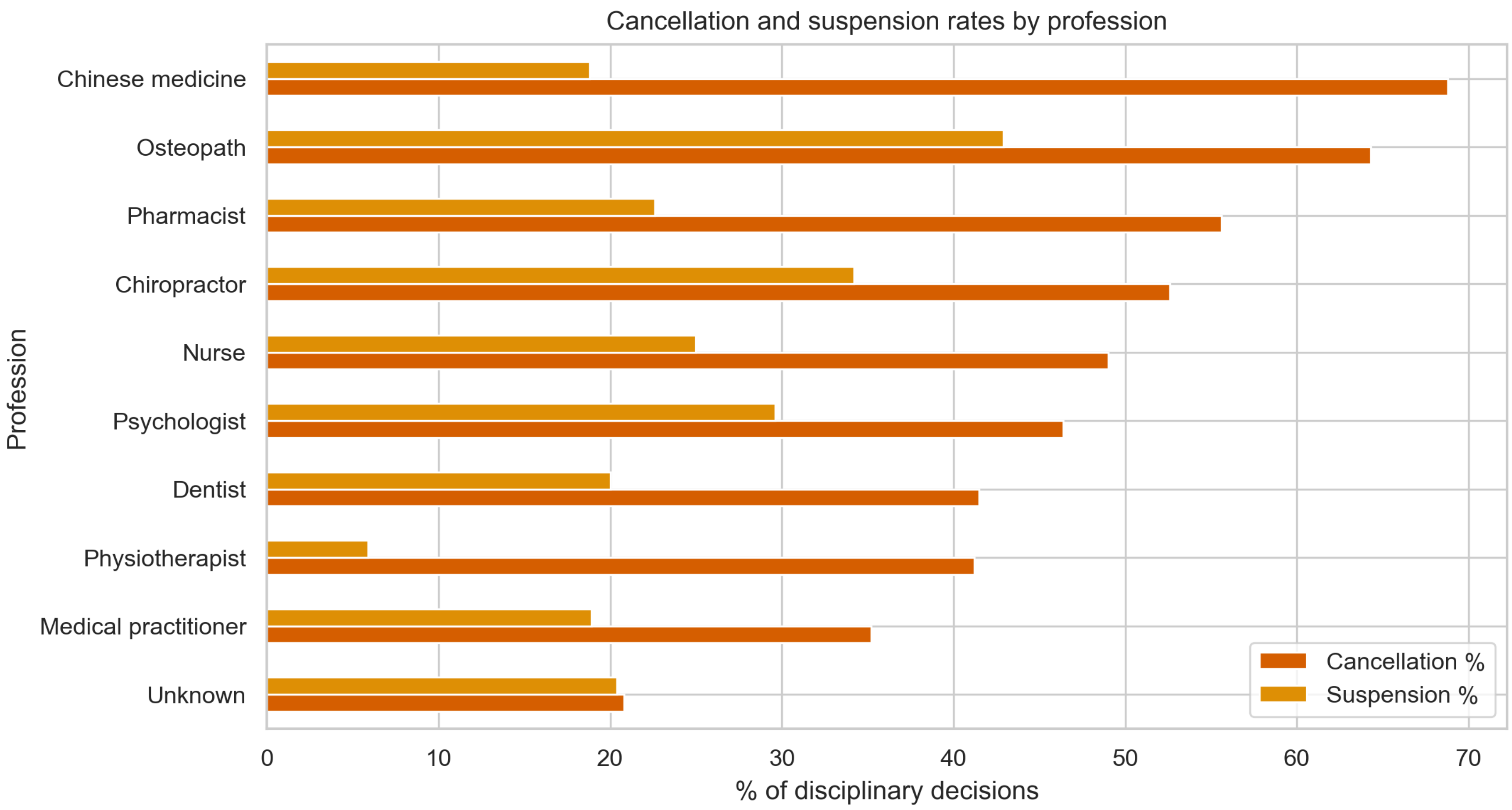

**Figure S3. Misconduct Co-occurrence Heatmap**

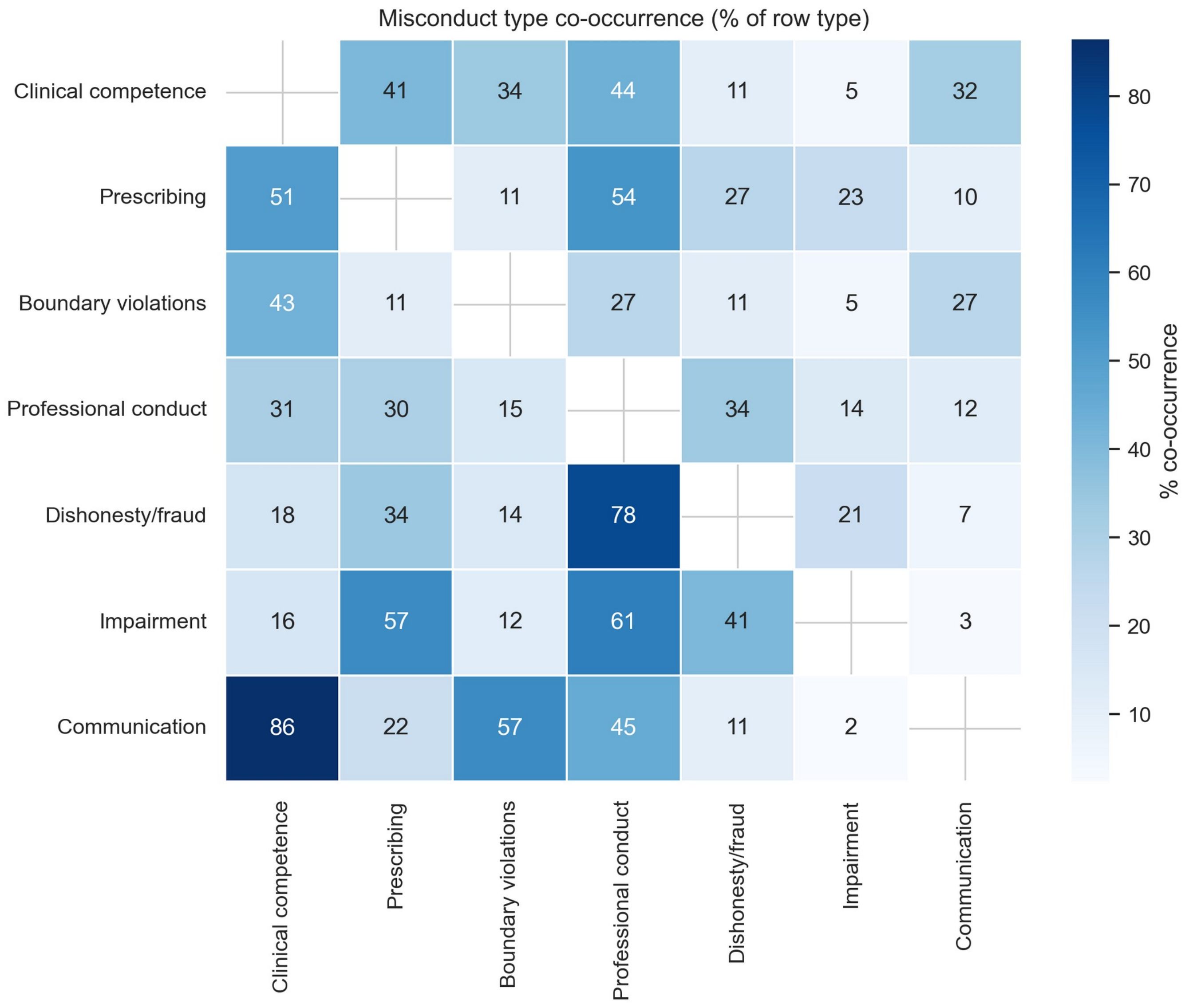

**Figure S4. Misconduct Distribution by Profession (Stacked Bar)**

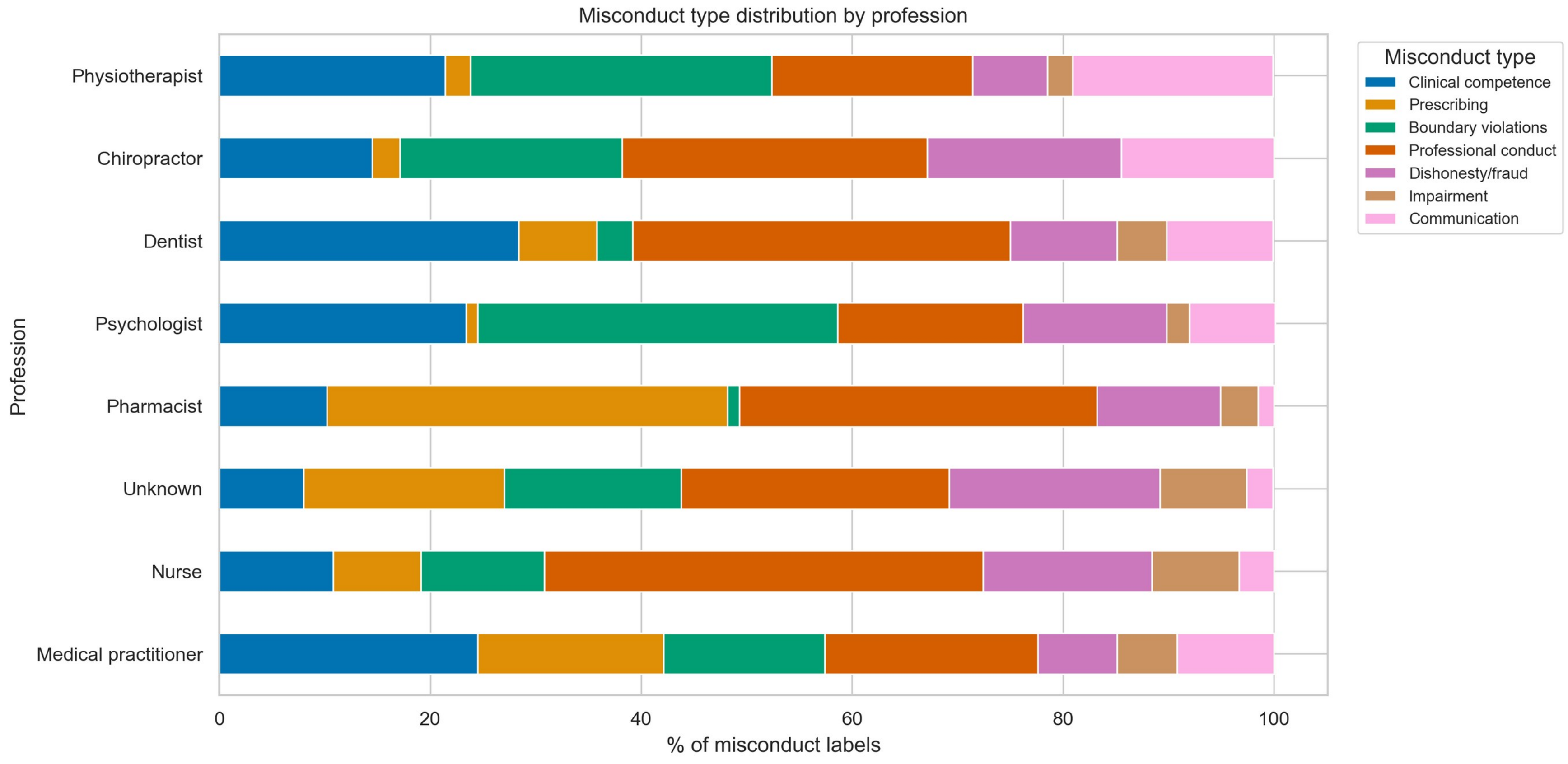

**Figure S5. Top 20 Medications in Prescribing Misconduct Decisions**

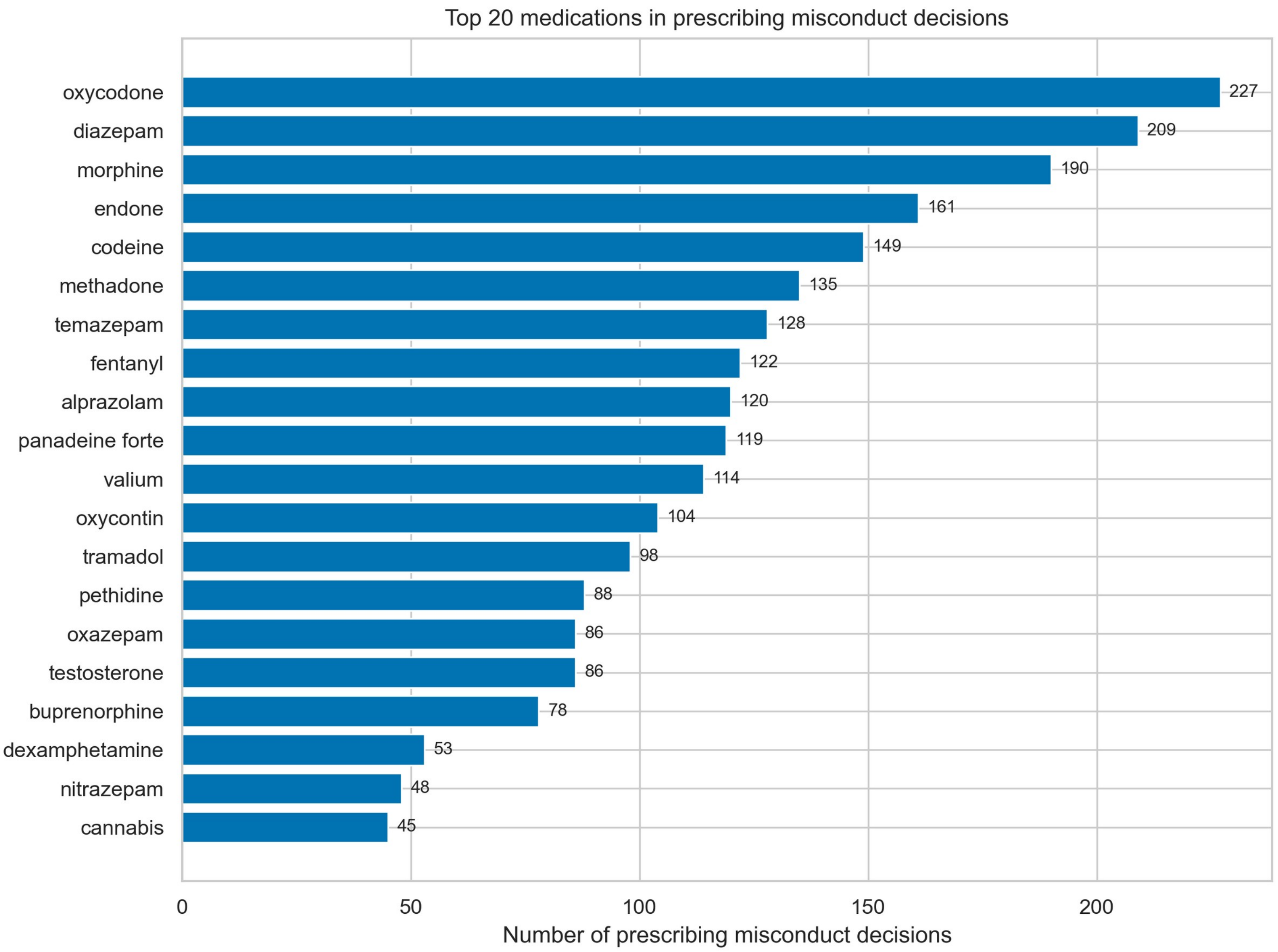

**Figure S6. Annual Decisions with Trend Line**

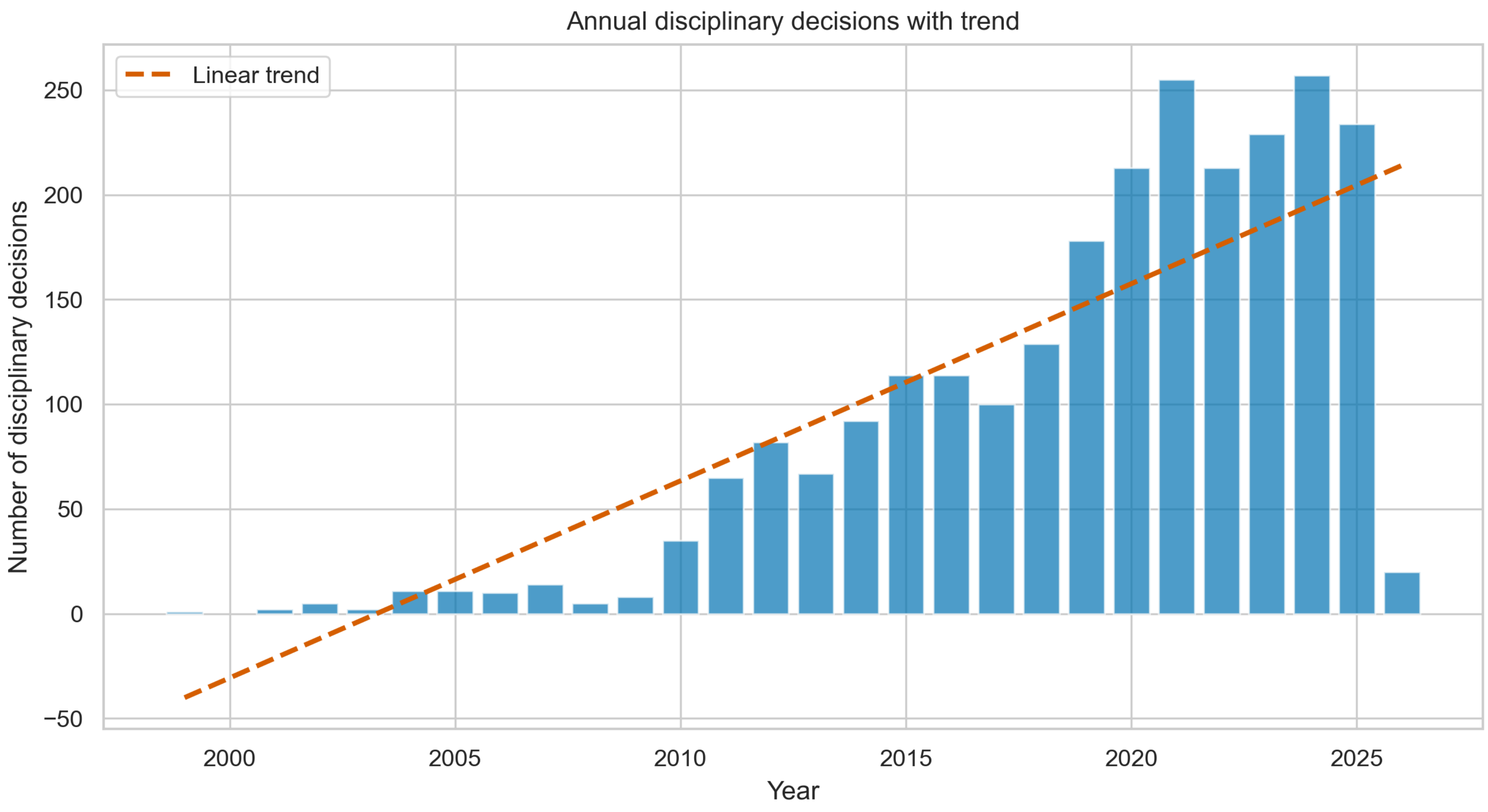

**Figure S7. Outcome Rates by Jurisdiction (Grouped Bar)**

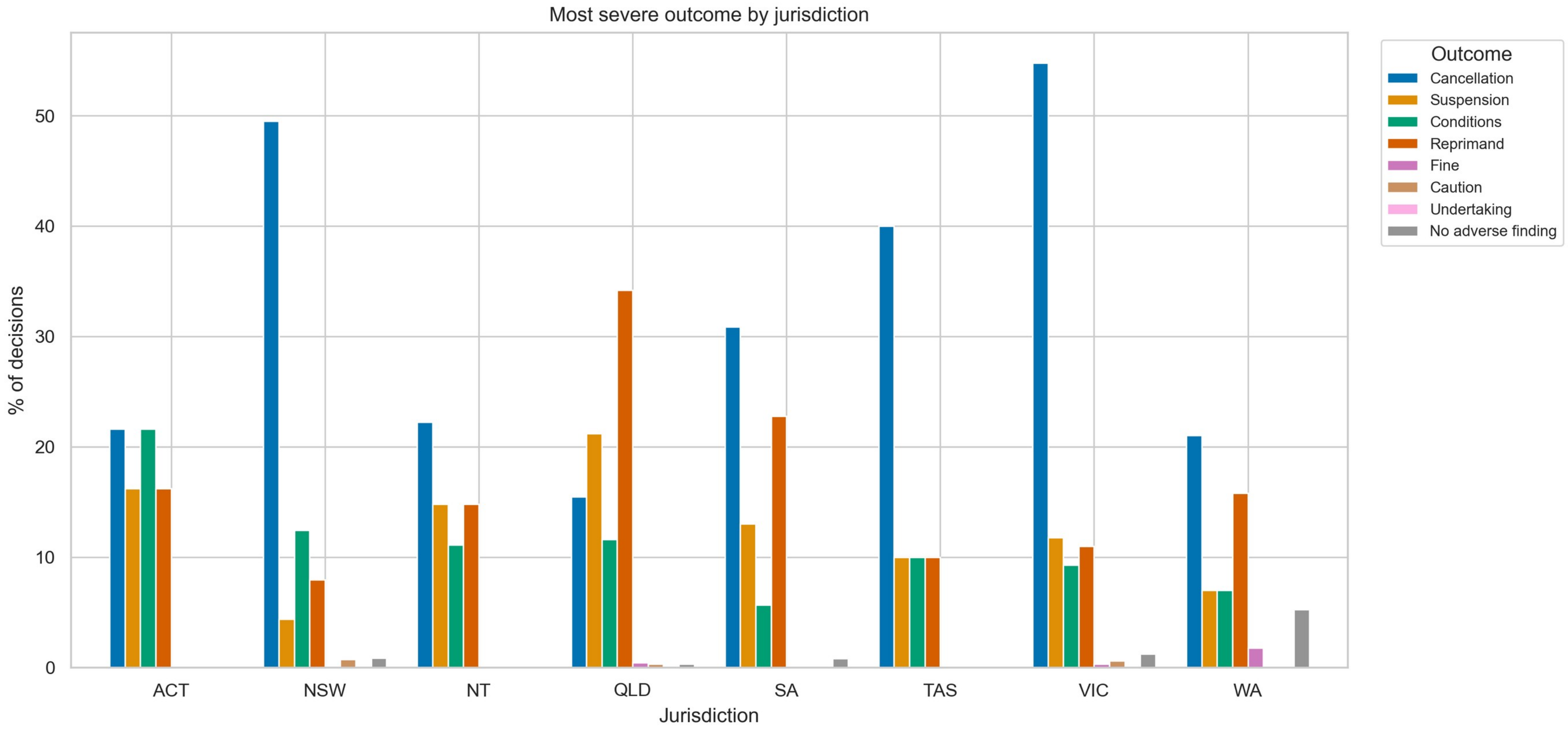

**Figure S8. Misconduct Type Prevalence by COVID-19 Period**

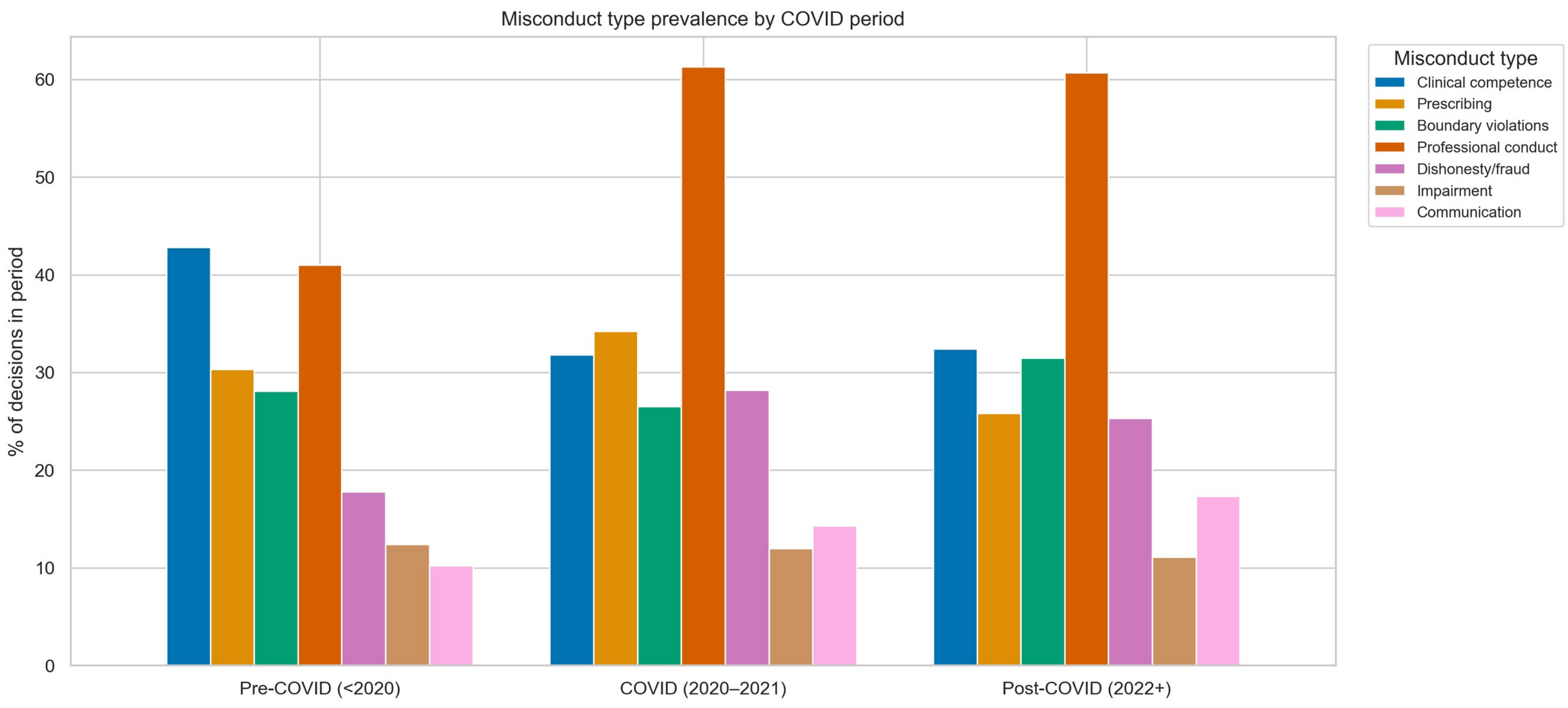

**Figure S9. Confusion Matrix: Is-Disciplinary Classification**

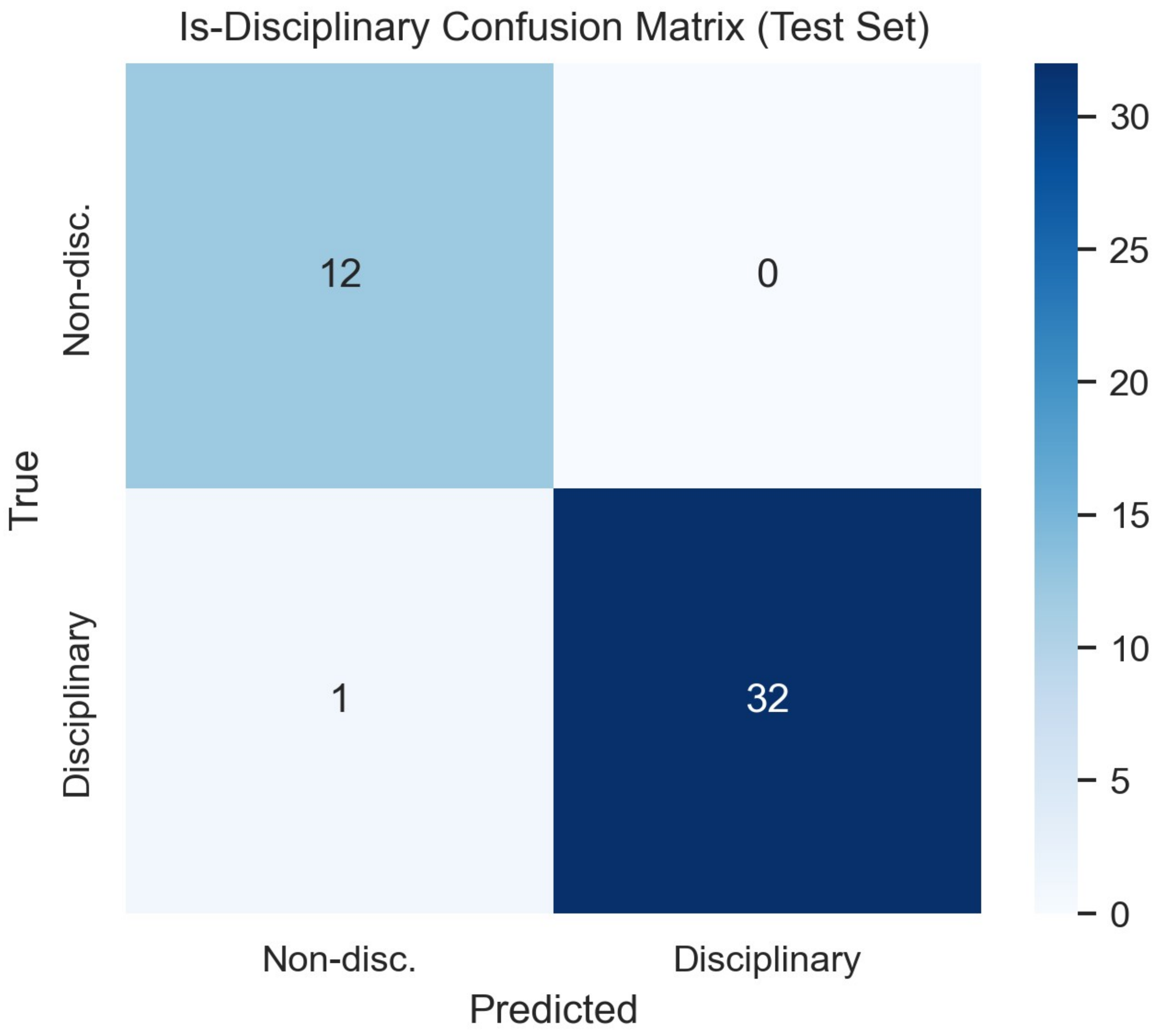

**Figure S10. Confusion Matrices: Misconduct L1 Classification (Per-Class)**

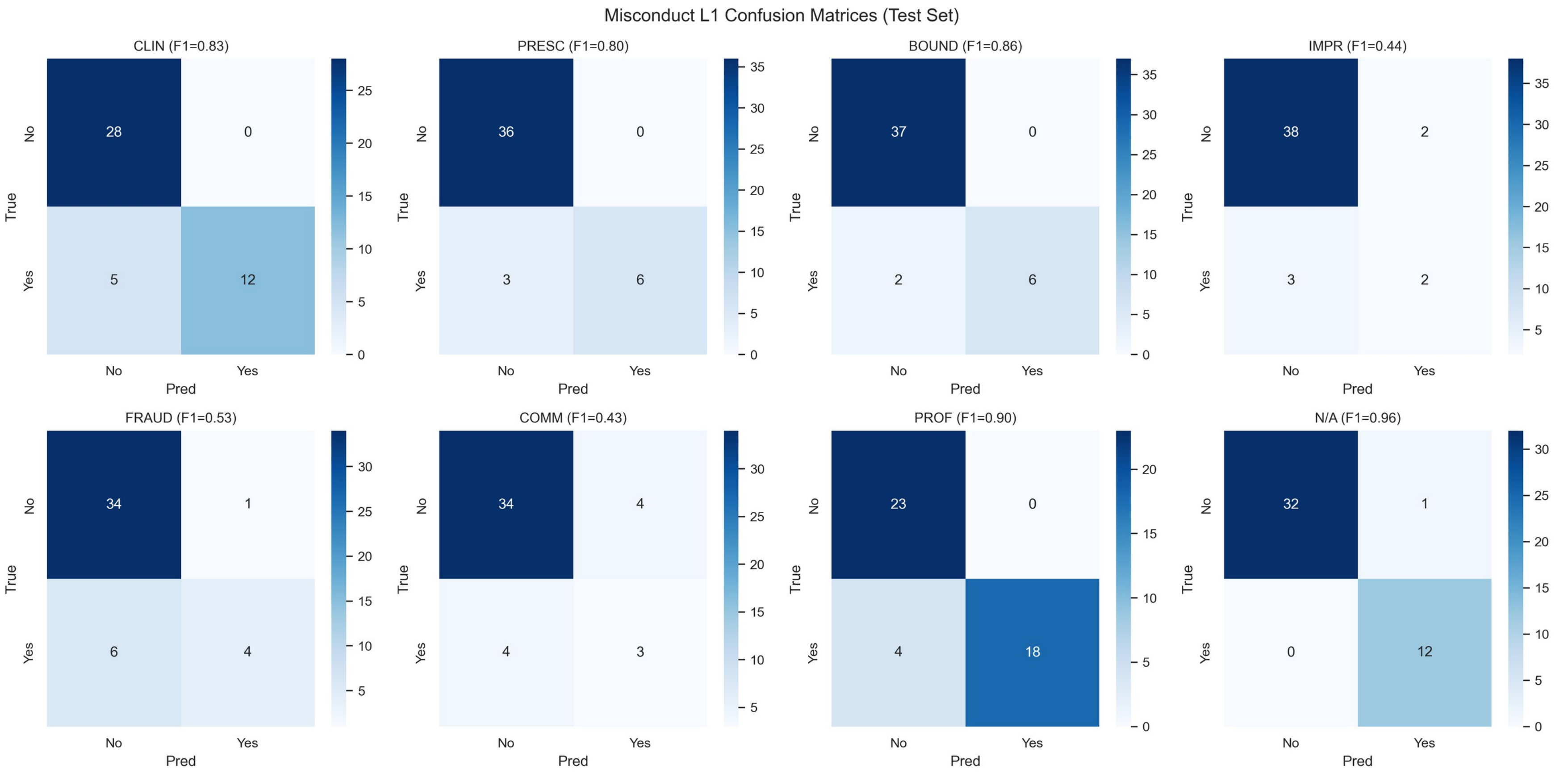

**Figure S11. Confusion Matrix: Finding Classification**

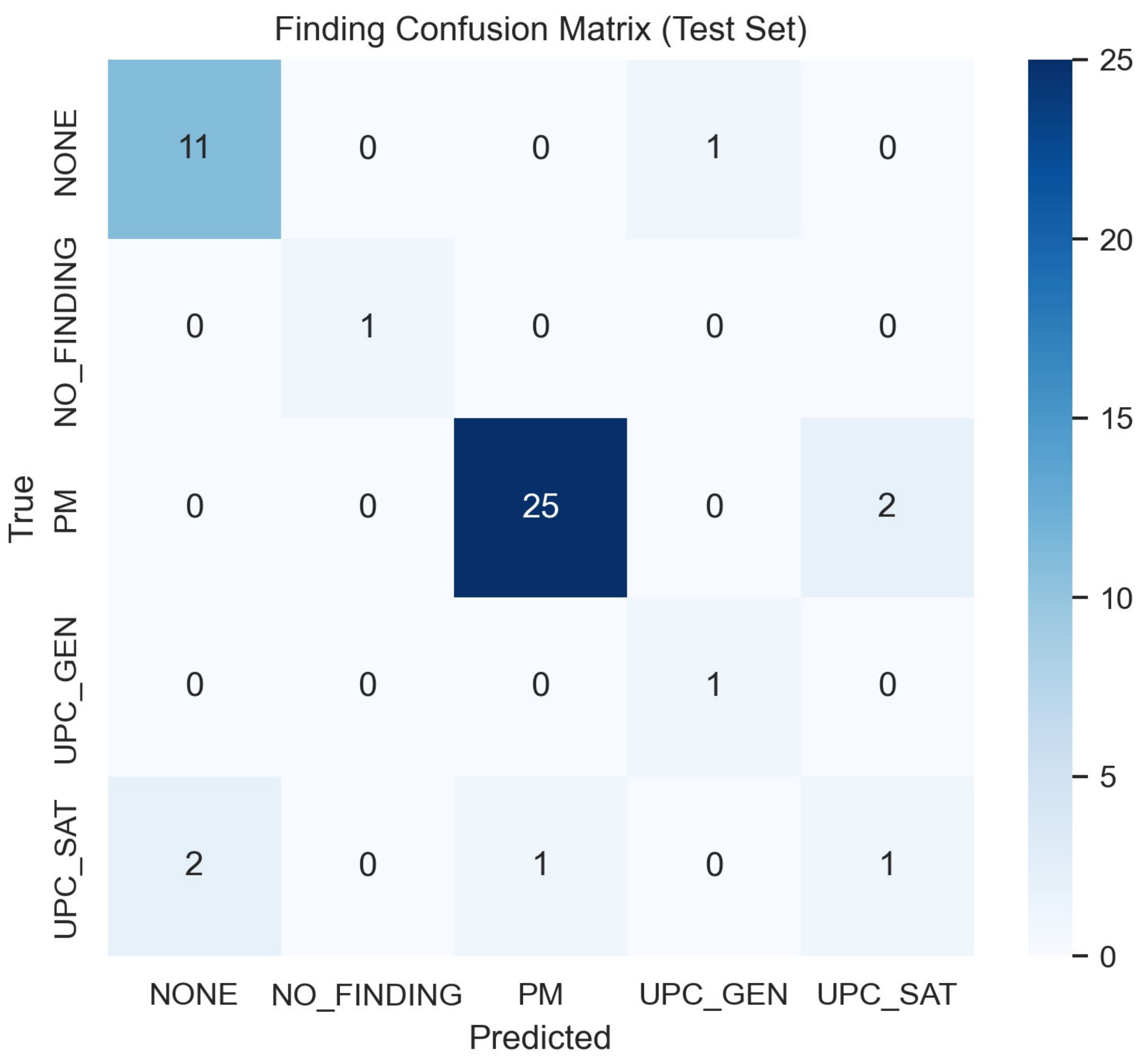

**Figure S12. Classification Performance by Threshold**

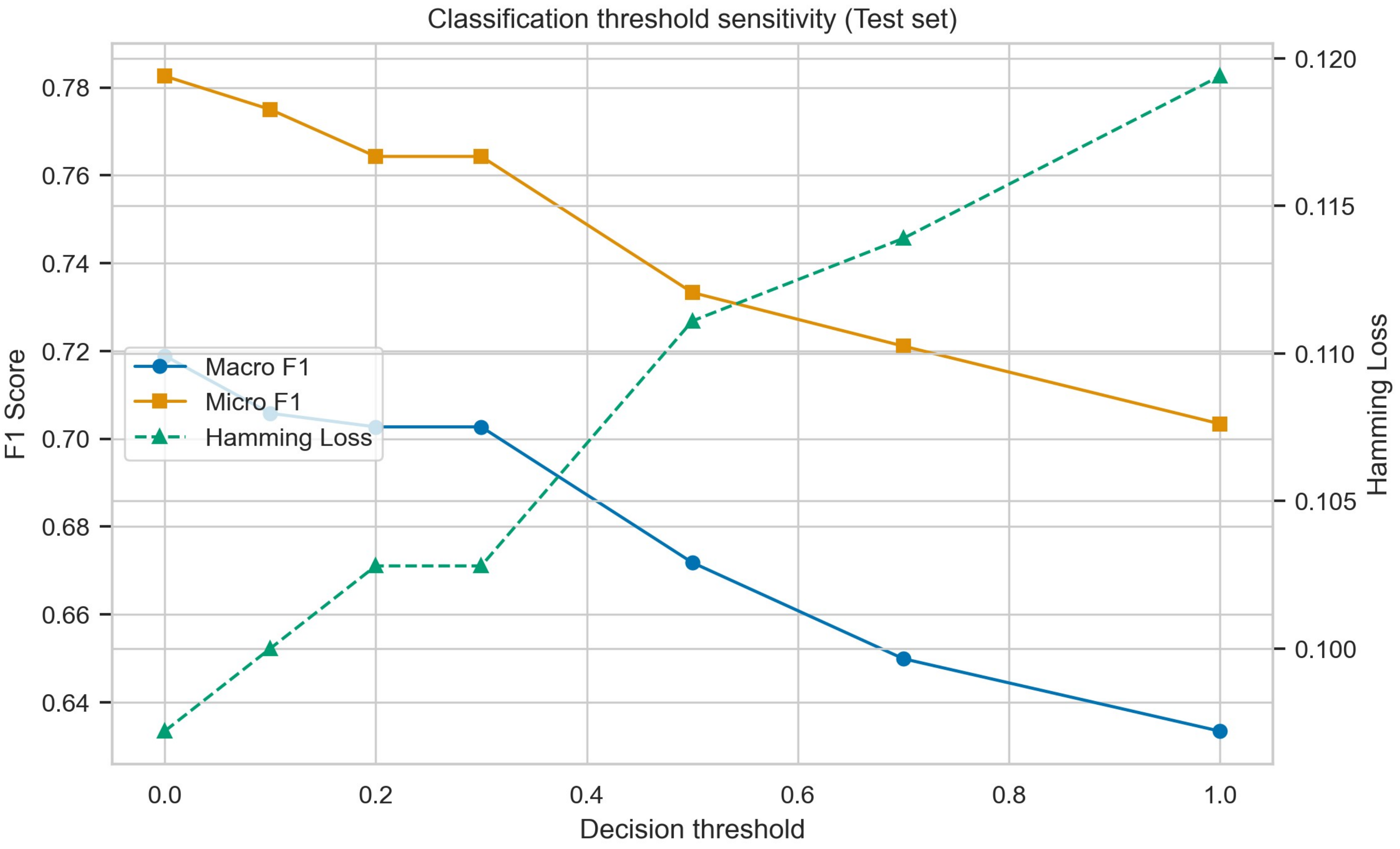

**Figure S13. Corpus Assembly and the Samples Used in the Study (Larger Format of Figure 1)**

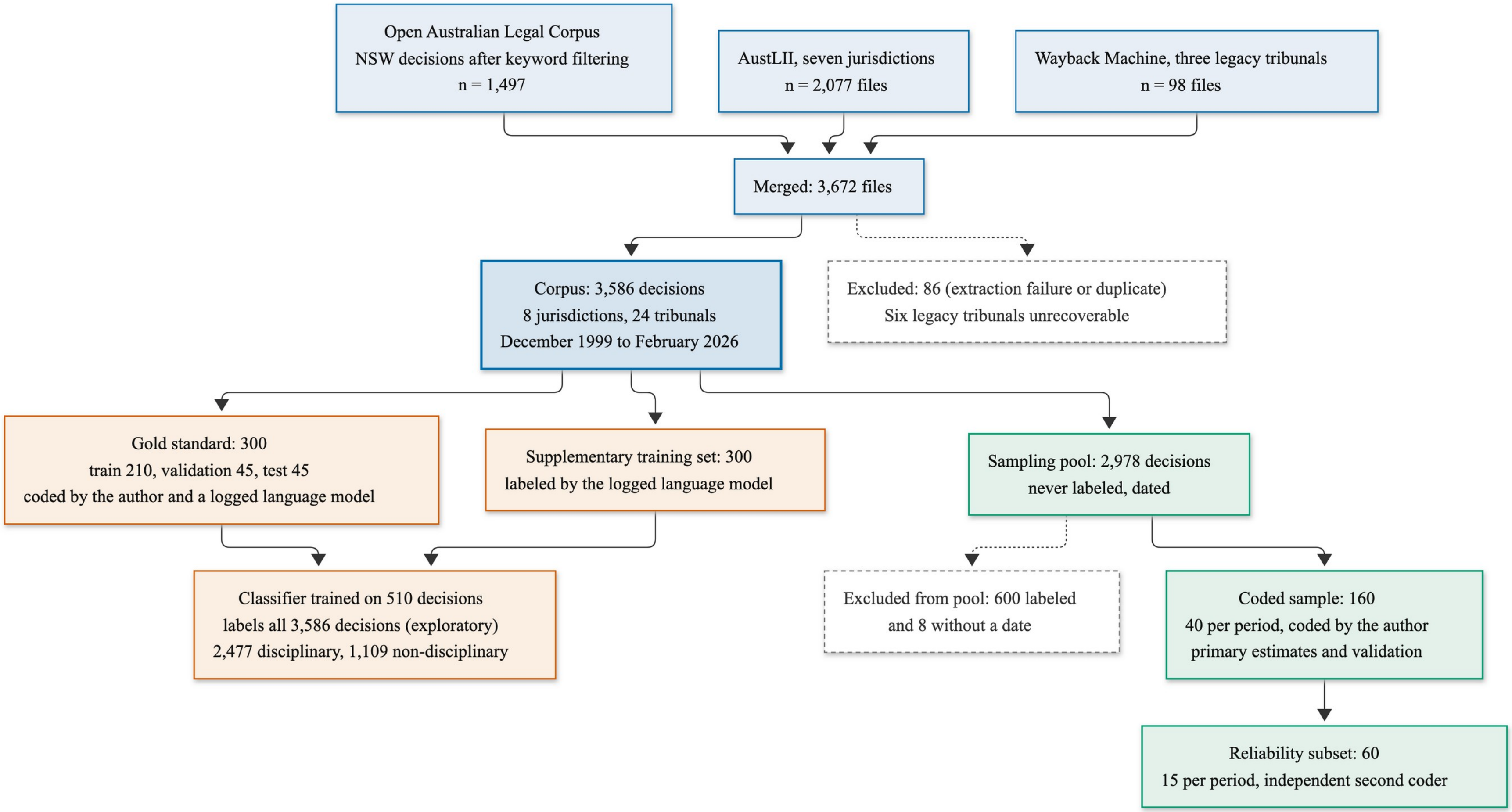

**Figure S14. Misconduct Type Co-occurrence Network**

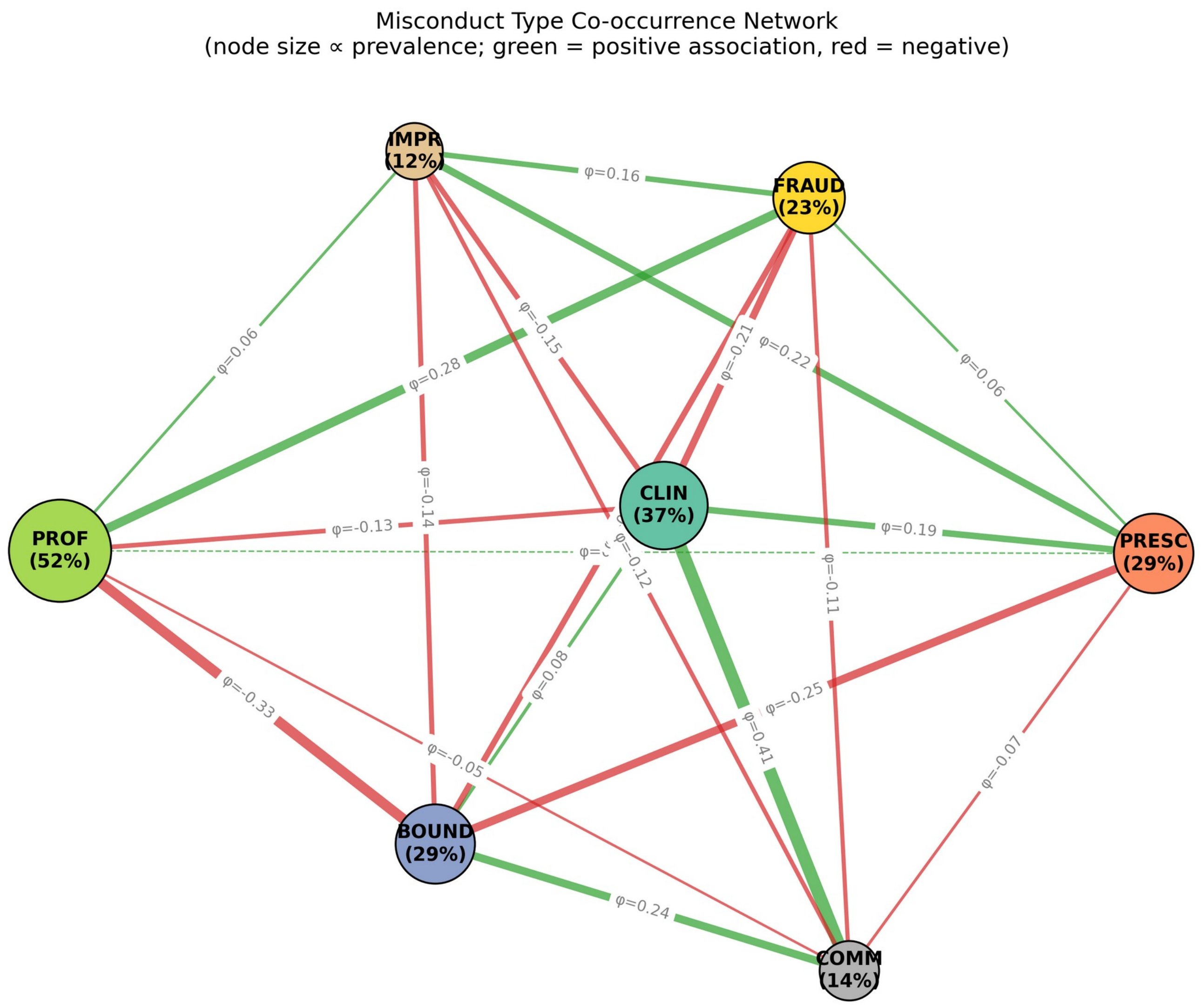

**Figure S15. Classifier-Labelled Misconduct Type Proportions by Year, 1999–2026 (Exploratory; 2026 Includes Decisions to 9 February Only)**

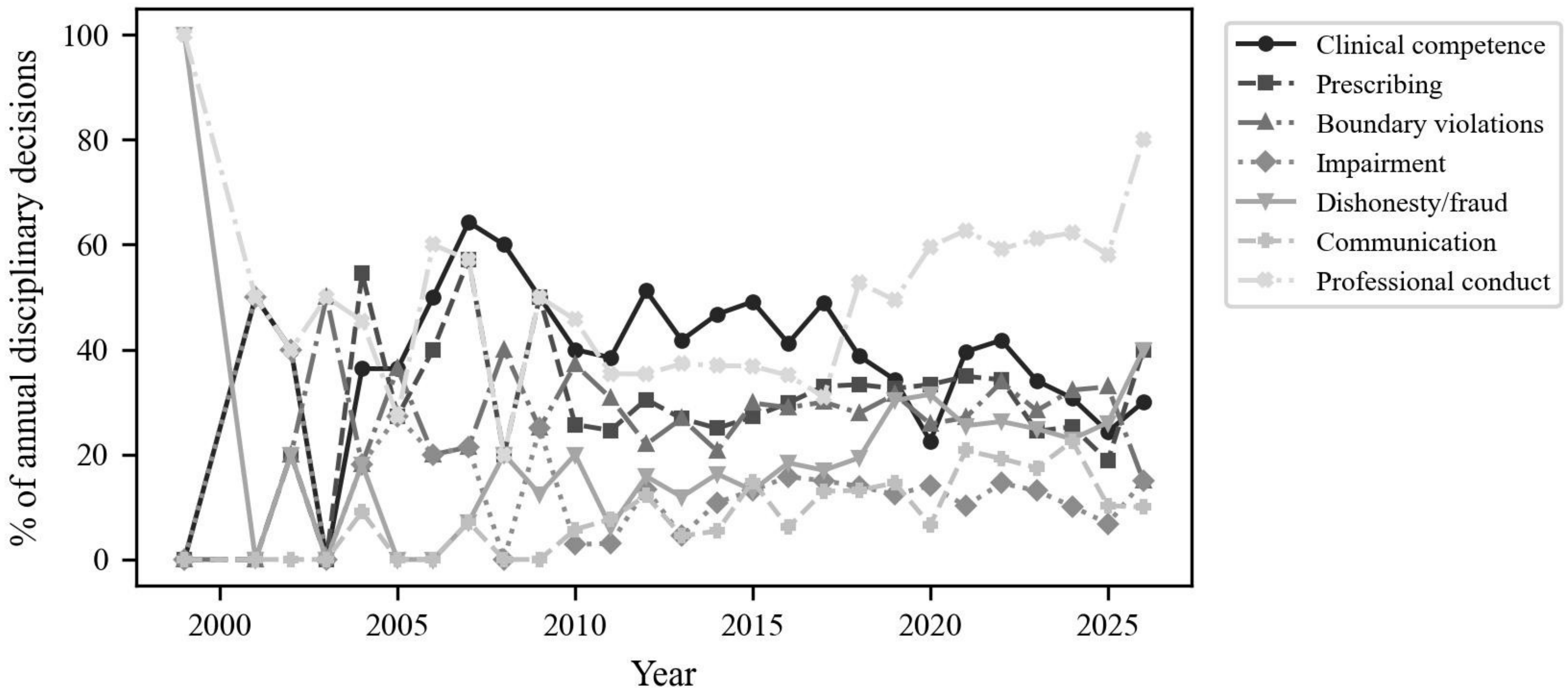


The dishonesty/fraud and communication trends in this figure do not pass the pre-specified check (Table S29).

### Appendix E: Classifier Technical Details

#### E.1 Model Architecture and Selection Protocol

All three classifiers use TF-IDF features (10,000 features, unigrams and bigrams, sublinear term frequency, minimum document frequency 2, maximum document frequency 0.95) and logistic regression with balanced class weights. The protocol below was written before any model was trained.

- **Training sets (fixed in advance):** (A) the 210 gold-standard training decisions; (B) set A plus 300 further corpus decisions, disjoint from the gold standard, labelled by the same logged language-model pass used as second coder (300 of 300 coded; 0 left out).
- **Selection:** for each training set and classifier, the regularisation strength C was chosen from [0.5, 1, 5, 10, 50, 100] by validation score (n = 45), ties going to the smaller C. The training set was then chosen by validation score, ties going to A. The test set was not used for any choice.
- **Misconduct classifier:** one-vs-rest over 7 misconduct types plus not applicable. Validation macro-F1: A 0.714 (C = 0.5), B 0.770 (C = 50). Chosen: B.

- **Finding classifier:** 5 classes (PM, UPC_SAT, UPC_GEN, NO_FINDING, NONE for non-disciplinary). Validation macro-F1: A 0.360 (C = 5), B 0.620 (C = 5). Chosen: B.
- **Is-disciplinary classifier:** binary, sharing the misconduct classifier's training set and vectoriser. Validation F1: A 0.954, B 0.985 (C = 5). Chosen: B.
- **Outcome extraction:** rule-based regular expressions targeting the Orders/Decision sections, with patterns informed by statutory language and common tribunal phrasing. It is not trained, and was evaluated on the 8 standard outcome categories (other orders excluded).

#### E.2 Test Set Performance (N = 45, scored once)

This is the first, gold-standard test set. Performance on the representative coded sample (n = 160) is in Tables S29 and S30 and main text Table 3.

*Is-Disciplinary Classification*

| Metric | Value |
|---|---|
| Accuracy | 97.8% |
| F1 (disciplinary) | 0.985 |
| F1 (non-disciplinary) | 0.960 |

**Confusion matrix:**

| | Predicted Non-Disc | Predicted Disc |
|---|---|---|
| Actual Non-Disc | 12 | 0 |
| Actual Disc | 1 | 32 |

*Multi-Label Misconduct Classification*

| Category | Precision | Recall | F1 | Support |
|---|---|---|---|---|
| CLIN | 1.000 | 0.706 | 0.828 | 17 |
| PRESC | 1.000 | 0.667 | 0.800 | 9 |
| BOUND | 1.000 | 0.750 | 0.857 | 8 |
| IMPR | 0.500 | 0.400 | 0.444 | 5 |
| FRAUD | 0.800 | 0.400 | 0.533 | 10 |
| COMM | 0.429 | 0.429 | 0.429 | 7 |
| PROF | 1.000 | 0.818 | 0.900 | 22 |

Overall: Macro-F1 = 0.719 (over the 7 types plus not applicable), Micro-F1 = 0.783, Hamming loss = 0.097

*Finding Classification*

| Category | Precision | Recall | F1 | Support |
|---|---|---|---|---|
| NONE | 0.846 | 0.917 | 0.880 | 12 |
| NO_FINDING | 1.000 | 1.000 | 1.000 | 1 |
| PM | 0.962 | 0.926 | 0.943 | 27 |

| Category | Precision | Recall | F1 | Support |
|---|---|---|---|---|
| UPC_GEN | 0.500 | 1.000 | 0.667 | 1 |
| UPC_SAT | 0.333 | 0.250 | 0.286 | 4 |

Overall: Accuracy = 86.7%, Macro-F1 = 0.755

*Outcome Extraction (Regex)*

| Category | Precision | Recall | F1 | Support |
|---|---|---|---|---|
| CANCEL | 0.529 | 1.000 | 0.692 | 9 |
| SUSPEND | 0.333 | 0.500 | 0.400 | 4 |
| CONDITIONS | 0.615 | 0.615 | 0.615 | 13 |
| REPRIMAND | 0.875 | 0.875 | 0.875 | 16 |
| CAUTION | 1.000 | 0.500 | 0.667 | 2 |

Overall: Micro-F1 = 0.694, Macro-F1 = 0.406

#### E.3 Error Patterns

- Total test decisions with at least one error: 32/45 (71.1%)
- Misconduct errors: 22/45 decisions affected
  - False negatives: FRAUD (6), CLIN (5), COMM (4), PROF (4), IMPR (3), PRESC (3), BOUND (2)
  - False positives: COMM (4), IMPR (2), FRAUD (1), N/A (1)
- Outcome errors: 21/45 decisions affected
  - False positives: CANCEL (8), CONDITIONS (5), SUSPEND (4), REPRIMAND (2), UNDERTAKING (1)
  - False negatives: CONDITIONS (5), SUSPEND (2), REPRIMAND (2), CAUTION (1)
- Finding errors: 6/45; is-disciplinary errors: 1/45
- Text length was not significantly associated with error (Mann-Whitney $p = 0.85$)

#### E.4 Sensitivity Analyses Summary

1. **Appeal exclusion:** Removing 516 appeal decisions (14.4% of the corpus; disciplinary decisions 2,477 to 2,374) changed misconduct prevalence by at most 0.6 percentage points (Table S13).
2. **Temporal split:** Pre-2015 (n = 410) vs post-2015 (n = 2,056): PROF, FRAUD, and COMM were more prevalent after 2015 and CLIN less prevalent; BOUND, PRESC, and IMPR did not differ significantly (Table S14).
3. **Classification threshold:** The default threshold (0) gave macro-F1 = 0.719 and micro-F1 = 0.783; the best threshold on the test set was 0 (macro-F1 = 0.719). Corpus-wide predictions use the default threshold (Table S16).
4. **Per-profession performance:** Macro-F1 ranged from 0.250 (physiotherapist, n = 3) to 0.638 (nurse, n = 9). Small test subsets limit interpretability (Table S17).
5. **Bootstrap CIs:** 95% confidence intervals for misconduct prevalence were within +/- 2.2 percentage points of the classifier’s corpus-wide point estimates. They reflect sampling variability only, not classification error.
6. **Author-only reference (post hoc):** Against the author’s blind codes alone, misconduct macro-F1 was 0.721 (gold standard 0.719) (Table S26).

### Appendix F: BERTopic Output

BERTopic was applied with MiniLM sentence embeddings, UMAP dimensionality reduction, and HDBSCAN clustering. The model identified 9 topics plus an outlier cluster (Topic -1).

#### F.1 Topic Summary

| Topic | Count | Label | Top Keywords |
|---|---|---|---|
| -1 | 444 | Outliers | tribunal, practitioner, health, decision, respondent, order, costs, commission, council, dr |
| 0 | 1,095 | General disciplinary (NSW-dominant) | tribunal, practitioner, decision, health, board, dr, commission, respondent, order, patient |
| 1 | 991 | Victorian proceedings | board, vcat, tribunal, australia, member, order, practitioner, medical, registration, dr |
| 2 | 847 | Queensland proceedings | qcat, tribunal, respondent, board, practitioner, dr, order, decision, health, conduct |
| 3 | 71 | Dental matters | dental, dr, board, tribunal, dentist, patient, practice, treatment, registration, health |
| 4 | 40 | ACT proceedings | acat, act, tribunal, respondent, practitioner, board, dr, health, order, registration |
| 5 | 39 | Pharmacy matters (QLD) | pharmacy, board, mr, qcat, pharmacist, tribunal, respondent, practice, registration, health |
| 6 | 22 | Temporal cluster (2022) | december 2022, ibid, ts, 2022, mr, paragraph, ms, respondent, order, practice |
| 7 | 22 | Transport/driving | transport, applicant, driver, mr, licence, authority, service, decision, vehicle, taxi |
| 8 | 15 | Appeal matters | appeal, appellant, evidence, decision, tribunal, ground, order, mr, error, law |

#### F.2 Interpretation

The topic model primarily captured jurisdictional and structural variation in the corpus rather than thematic misconduct types. Topics 0–2 correspond to the three largest jurisdictions (NSW, Victoria, Queensland), reflecting distinctive tribunal naming conventions, procedural language, and case formatting. Topics 3–5 captured profession-specific and smaller jurisdiction clusters. Topic 7 (transport/driving) likely represents a small number of non-health practitioner decisions that passed the initial filtering stage. The predominance of jurisdictional over thematic clustering suggests that structural and procedural language differences between tribunals are stronger signals than misconduct-specific content, a finding that may inform future work on jurisdiction-agnostic text preprocessing.

#### F.3 Visualisations

Topic model visualisations (a topic hierarchy dendrogram, a topic bar chart, an inter-topic distance map, and a topic–term heatmap) were generated with BERTopic's built-in plotting functions; they are exploratory and are not reproduced here.

### Appendix G: The Coded Sample and Its Re-Coding

**Design.** The sampling pool was every decision never labelled by anyone, with at least 500 characters of text and a known date (2,978 decisions). Pool sizes by period were 1999-2009: 77, 2010-2014: 416, 2015-2019: 829, 2020-2026: 1,656. Forty decisions were drawn at random from each period (seed 20261106), and item order was shuffled. The protocol (written before sampling) fixed the models, metrics, weights, bootstrap, and secondary analyses.

**First coding, set aside.** The first coding was frozen and scored under the protocol (classifier weighted misconduct macro-F1 0.43, disciplinary accuracy 0.81; language-model coder 0.47 and 0.81). Diagnostic checks then showed that many rows did not describe the decision in that row's text. Two indicators are shown below for each coding: agreement between the recorded profession and the logged language-model coder's profession, and whether each row's free-text note is more similar (TF-IDF cosine) to its own decision text than to any other text in the same set. The first coding was set aside unchanged, and its results are not used.

| Coding | Profession agrees with logged coder | Note best matches its own text |
|---|---|---|
| Gold standard (author, blind) | 288/300 | 251/300 |
| Coded sample, first coding (set aside) | 106/160 | 57/160 |
| Coded sample, re-code (used) | 156/160 | 110/160 |

**Re-code.** The author re-coded all 160 decisions from the packet texts, with the same fields and rules, in a fresh workbook that required for each row the practitioner's name (or anonymised label) taken from that row's text. The entry was found in its own text for 160 of 160 rows. The author declared that each row was coded only from its own text file, without assistance and without consulting the first coding. The re-code is a departure from the protocol and was recorded in dated amendments before the re-coded sample was scored. When re-coding, the author knew the aggregate results of the first scoring but had seen no prediction for any individual decision.

**Language-model coder.** The coder was run once, before either coding was scored, with the prompt, schema and settings used for the gold standard. Its outputs were scored once against each coding and never edited. On the re-code, it met the accuracy thresholds of its protocol but not the requirement that the 95% CI for its advantage over the classifier exclude zero (difference 0.069, 95% CI -0.009 to 0.152).

### Appendix H: Independent Second Coder

**Design.** Under a protocol fixed before recruitment, an independent coder recruited through Prolific coded 60 of the 160 coded-sample decisions (15 per period, drawn at random) in two sessions of 30, blind to the author's codes, to any machine output and to the manuscript. The coder's codes are used only to report reliability; the author's codes were not changed. Kappa is Cohen's kappa with a 95% percentile bootstrap interval (2,000 resamples, seed 20261109). A misconduct, subtype or outcome code on a decision a coder called non-disciplinary counts as absent. Kappa is not estimated for binary fields where either coder recorded fewer than 5 positives.

#### Table S33. Agreement Between the Author and the Independent Second Coder (n = 60)

| Field | Kappa (95% CI) | Raw agreement (%) | Positives, author / second coder |
|---|---|---|---|
| Disciplinary status | 0.79 (0.60 to 0.95) | 91.7 | – |
| Finding | 0.76 (0.63 to 0.88) | 83.3 | – |
| Profession | 0.91 (0.80 to 1.00) | 95.0 | – |
| Clinical competence | 0.74 (0.52 to 0.92) | 90.0 | 15 / 17 |
| Prescribing/dispensing | 0.89 (0.69 to 1.00) | 96.7 | 10 / 12 |
| Boundary violations | 0.95 (0.84 to 1.00) | 98.3 | 15 / 14 |
| Impairment | 0.78 (0.38 to 1.00) | 96.7 | 5 / 5 |
| Dishonesty/fraud | 0.59 (0.33 to 0.82) | 85.0 | 12 / 17 |

| Field | Kappa (95% CI) | Raw agreement (%) | Positives, author / second coder |
|---|---|---|---|
| Communication | not estimated | 91.7 | 2 / 7 |
| Professional conduct | 0.60 (0.39 to 0.80) | 81.7 | 24 / 17 |
| Cancellation | 0.88 (0.68 to 1.00) | 96.7 | 11 / 9 |
| Suspension | 0.84 (0.55 to 1.00) | 96.7 | 8 / 6 |
| Conditions | 0.93 (0.81 to 1.00) | 96.7 | 21 / 21 |
| Reprimand | 0.96 (0.88 to 1.00) | 98.3 | 23 / 22 |
| Fine | not estimated | 100.0 | 2 / 2 |
| Caution | not estimated | 98.3 | 0 / 1 |
| Undertaking | not estimated | 100.0 | 1 / 1 |
| No adverse finding | not estimated | 100.0 | 1 / 1 |
| Other orders | 0.37 (0.12 to 0.58) | 71.7 | 15 / 24 |
| Sexual boundary subtype (both coded boundary, n = 14) | 0.76 (0.00 to 1.00) | 92.9 | 11 / 12 |
| Emotional boundary subtype (both coded boundary, n = 14) | not estimated | 28.6 | 11 / 1 |
| Financial boundary subtype (both coded boundary, n = 14) | not estimated | 100.0 | 1 / 1 |

Positives are shown for the misconduct, subtype and order fields; disciplinary status, finding and profession are reported by kappa and raw agreement only.

**Table S34. Misconduct Type Prevalence on the Subset, by Coder (Unweighted; Among Each Coder’s Disciplinary Decisions: Author n = 41, Second Coder n = 46)**

| Misconduct type | Author % (95% CI) | Second coder % (95% CI) | Difference, percentage points (95% CI) |
|---|---|---|---|
| Clinical competence | 36.6 (22.2 to 51.2) | 37.0 (23.2 to 51.1) | +0.4 (-10.6 to +11.7) |
| Prescribing/dispensing | 24.4 (12.5 to 39.5) | 26.1 (14.9 to 40.0) | +1.7 (-4.1 to +8.2) |
| Boundary violations | 36.6 (22.0 to 51.2) | 30.4 (17.4 to 44.2) | -6.2 (-12.9 to -1.5) |
| Impairment | 12.2 (2.7 to 23.3) | 10.9 (2.3 to 20.5) | -1.3 (-8.2 to +5.8) |
| Dishonesty/fraud | 29.3 (15.9 to 43.5) | 37.0 (23.3 to 51.1) | +7.7 (-5.2 to +20.7) |
| Communication | 4.9 (0.0 to 12.5) | 15.2 (6.0 to 26.7) | +10.3 (+1.7 to +20.0) |
| Professional conduct | 58.5 (43.2 to 73.7) | 37.0 (23.9 to 51.2) | -21.6 (-36.6 to -6.6) |

The subset is unweighted, so these shares differ from the weighted coded-sample estimates in the main text.

**Table S35. Field-Level Disagreements by Type and Theme (n = 90)**

| Type | Theme | Fields |
|---|---|---|
| scope | procedural, review, interim or costs decision: disciplinary-status scope (field rule vs guideline 8.4) | 20 |
| scope | conduct discussed in the reasons but not separately determined | 5 |
| scope | orders proposed for submissions but not yet made | 4 |
| scope | suspension already in place: recognised vs ordered in this decision | 1 |
| definitional | dishonest conduct: dishonesty or fraud vs professional conduct, or which dishonest acts count | 11 |
| definitional | emotional subtype on sexual-boundary cases | 9 |
| definitional | non-publication / suppression order counted as a non-standard order | 5 |
| definitional | categorising a health-and-performance conditions review | 4 |
| definitional | non-review or preclusion period after cancellation counted as a non-standard order | 3 |
| definitional | disqualification of a person no longer registered: cancellation vs non-standard order | 3 |
| definitional | non-practitioner matter: ‘other’ vs ‘unknown’ | 3 |
| definitional | treating a spouse: boundary violation vs professional conduct (patient-relationship rule) | 2 |
| definitional | professional conduct alongside impairment in a penalty decision | 1 |
| definitional | underlying addiction coded as impairment (root-cause rule) | 1 |
| definitional | self-prescribing: prescribing alone vs prescribing plus clinical competence | 1 |
| definitional | failure to advise the patient: communication failure vs clinical competence | 1 |
| definitional | communication failure for confidentiality or communication conduct | 1 |
| definitional | drug diversion: clinical competence vs prescribing | 1 |
| definitional | setting aside of immediate action counted as a non-standard order | 1 |
| definitional | communication failure for misleading information | 1 |

| Type | Theme | Fields |
|---|---|---|
| definitional | mapping WA ‘improper conduct’ onto the finding scale | 1 |
| definitional | regulatory dispensing breach: prescribing alone vs prescribing plus professional conduct | 1 |
| definitional | education-course requirement counted as a non-standard order rather than conditions | 1 |
| reading | which finding the tribunal actually made | 4 |
| reading | whether sexual activity is described | 2 |
| reading | false statement to the Board not registered in one coder’s notes | 2 |
| reading | whether a consent particular was established | 1 |

Of the 90 field-level disagreements, 51 were definitional (both coders describe the same facts but apply a category or rule differently), 30 concerned scope (which proceeding or which part of the decision is coded) and 9 were differences in reading (the coders’ notes describe different facts). Of the scope disagreements, 15 follow from the five disagreements on disciplinary status, because the coding rules leave every other field blank for a non-disciplinary decision. The classification describes the cause of each disagreement and does not decide which code is correct; it was drafted with AI assistance from both coders’ notes and the decision texts and reviewed by the author (see AI Use Disclosure in the main text).

**Exploratory, not pre-specified.** Agreement on disciplinary status was kappa 0.59 in session 1 and 0.93 in session 2, and on the finding 0.65 and 0.86. The coder’s median time per decision was 116 seconds in session 1 and 84 seconds in session 2. Before session 2 the coder was reminded, in wording that applied to any coder, to read each decision in full rather than only the orders and catchwords; this instruction was not in the protocol.
